# Deep Learning Model and Multi Modal Late Fusion For Predicting Adverse Events Following Cardiothoracic Surgery in the ICU Using STS Data and Time Series Intraoperative Data

**DOI:** 10.1101/2024.09.04.24312980

**Authors:** Rajashekar Korutla, Anne Hicks, Marko Milosevic, Dipti Kulkarni, Felistas Mazhude, Mehdi Mortazawy, Yashar Seyed Vahedein, Tyler Kelting, Jaime B Rabib, Qingchu Jin, Robert Kramer, Douglas Sawyer, Raimond L Winslow, Saeed Amal

**Affiliations:** The Roux Institute, Northeastern University, Portland, ME, USA; Maine Medical Center, Portland, ME, USA; Spectrum Healthcare Partners, Portland, ME, USA; The Roux Institute, Department of Bioengineering, College of Engineering at Northeastern University, United States; Coredio Corp., Mountain View, CA

**Keywords:** Artificial Intelligence, Deep Learning, Time-Series Intraoperative Data, Predictive Modeling, Cardiothoracic Surgery in ICU, Adverse Events, Ablation Study

## Abstract

Accurate prediction of post-operative adverse events following cardiothoracic surgery is crucial for timely interventions, potentially improving patient outcomes and reducing healthcare costs. By leveraging advanced deep learning techniques, this study highlights the transformative potential of incorporating intraoperative variables into predictive analytics models to enhance postoperative care for cardiothoracic surgery patients in the ICU. We developed deep learning predictive models for anticipating adverse events in patients following cardiothoracic surgery using a dataset from the Society of Thoracic Surgeons’ database (^4^) and intraoperative data. Our models perform late fusion by integrating static patient data and intra-operative time-series data, utilizing Fully Connected Neural Networks (FCNN) and long short-term memory (LSTM) networks, respectively. The hybrid model was validated through five-fold cross-validation, demonstrating robust performance with a mean AUC of 0.93, Sensitivity of 0.83 and Specificity of 0.89. This work represents a significant step forward in the proactive management of cardio thoracic surgery patients in the ICU by effectively predicting potential adverse events associated with mortality in the post operative period.

## 1 Introduction

Cardiothoracic surgery, despite advances in medical technology and practice, is fraught with risks of untoward events. To prevent these untoward complications, rapid and effective detection and interventions are crucial. The Society of Thoracic Surgeons (STS) developed a standardized national clinical data registry for cardiothoracic surgery in 1989 which facilitated the development of risk models to help guide real-time patient counseling and shared decision making for patients contemplating high risk surgery (Shahian 2018). Using the STS database, Reddy et al identified 17 post operative complications following open heart surgery associated with mortality in the post operative phase; they went on to demonstrate a disparity between the variation in mortality and the variation in incidence of the 17 complications between hospitals with low-, medium- and high-mortality rates (8.3% vs 10.0% vs 12.7%, P < 0.001, respectively). Subsequent studies support the assertion that early prediction of these “failure to rescue (FTR)” complications could facilitate early intervention and treatment to prevent mortality in the post operative period. A study by Gabriela O. Escalante et al. [2023] (^23^) used data provided by the Society of Thoracic Surgeons (STS) (^4^) from 20,950 consecutive patients to find that 1 or more of 4 complications viz. prolonged ventilation, stroke, renal failure, and unplanned reoperation, lead to mortality via Failure to Rescue. These findings asserted the association between FTR and hospital processes of care.

The STS database was also used by another Michigan study group to identify “failure to rescue” -like complications following esophagectomy, a procedure that definitively treats esophageal cancer and other esophageal diseases. Anastomotic leak is a dreaded complication of this procedure highly associated with morbidity and mortality and has received significant attention in the literature. Stanley Kalata et al. [2023] (^29^), analyzed STS data from 621 esophagectomies performed at the University of Michigan to find early post-operative complications associated with anastomotic leak. They found that early post operative atrial fibrillation and pneumonia were associated with the later finding of anastomotic leak. Cox proportional hazards analysis and got a sensitivity and specificity of 31.3% and 76.2% respectively for atrial fibrillation and 7.0% and 90.6% respectively for pneumonia. This finding is notable because atrial fibrillation and pneumonia are typically presented by POD#2-3, vs anastomotic leak, which is not typically identified until POD#8. Ferrando-Vivas P et al. [2022] (^28^) implemented risk models for predicting the outcomes viz. acute hospital mortality and 1-year mortality after cardiothoracic critical care admissions. They added pre-operative as well as intra-operative data from NHS cardiothoracic critical care units participating in CMP (Case Mix Program) to better the performance of their refitted multivariable model and got a c-index value of 0.892.

Jahan C. Penny-Dimri et al. [2022] (^20^) worked on reviewing 2,792 peer-reviewed studies of traditional machine learning (ML) techniques. They predicted adverse postoperative outcomes such as 30-day mortality and in-hospital mortality in cardiac surgery and got C-index scores which were 0.81 (0.78−0.84) and 0.79 (0.73−0.84) for pooled ML models and independent LR respectively. Dimitris Bertsimas et al. [2021] (^21^) explored various ML models to predict the outcomes of cardiac surgery and better resource optimization using data of more than 235,000 patients and 295,000 operations provided by the European Congenital Heart Surgeons Association (ECHSA) Congenital Database (^33^). One of their findings was that optimal classification trees (OCTs) got the highest AUC training score of 85.5% and AUC testing score of 86.2% for the prediction of mortality. OCTs aim to find the most accurate tree model by optimizing for the best possible splits in data. This motivated the use of XGBoost model which like OCTs is another decision tree-based ML algorithm and works on finding non-linear relationships while providing insights into feature importance. The study also suggested that age and weight were important factors to be considered while building prediction models.

Previous efforts to predict FTR have faced limitations, particularly in the transparency and robustness of risk estimators. Studies by Shahian et al. [2018] (^6^), O’Brien et al. [2018] (^5^), and Kurlansky et al. [2021] (^9^) have pointed out the insufficiency of using the c-statistic as a sole performance measure. Shahian et al. used entries from July 2011 to June 2014 in the STS Adult Cardiac Surgery Database (ACSD) to build parsimonious models that used backward selection from a full model. New versions of the ACSD risk models were built to predict renal failure in Coronary Artery Bypass Grafting, prolonged ventilation in Coronary Artery Bypass Grafting, and stroke in Valve procedures among others. Even though our study considered some of these targets, our final model combined all adverse effects to form a unified target column mostly due to limited data. Similarly, O’Brien et al. built separate statistical models for different surgeries and endpoints like stroke, renal failure, prolonged ventilation, mediastinitis/deep sternal wound infection to name a few. They also used data from ACSD such as 439,092 entries for coronary artery bypass grafting surgery, 81,588 entries for combined valve plus coronary artery bypass grafting surgery to name a few. The c-index score ranged from 0.588 for reoperation and 0.826 for renal failure. Kurlansky et al. used ACSD for modeling a predictive model. They included pre-operative and intra-operative features in their model and got a c-index score of 0.806. These gaps highlight the necessity for a more nuanced approach that integrates a broader spectrum of data and analytical techniques.

Already in 1993, a study by Mark H. Ebel (^27^) established the efficacy of Artificial Neural Networks to predict Failure to Survive with a positive predictive value of 97%. This reinforces the integration of neural networks into our model, especially considering that positive results were obtained in such early studies.

Arman Kilic et al. [2021] (^26^) employed an XGBoost model to predict the outcomes of Aortic Valve Replacement using entries from the STS National Database between 2007 and 2017. Some of the outcomes considered by the model were operative mortality, acute renal failure and deep sternal wound infection among others. The average observed-to-expected ratio of the model was 0.985. A study by Aixia Guo et al. [2021] (^16^) that employs an LSTM model to predict cardiovascular health trajectories using the Guideline Advantage (TGA) dataset (^35^) containing Electronic Health Record data from 70 outpatient clinics across the United States (US). They achieved an AUROC score as high as 0.97 for Body Mass Index (BMI). The study also identified deep sternal wound infection and renal failure as two of the major complications. Sundos Alabbadi et al. [2022] (^30^) used join point regression software on the National Inpatient Sample database (^36^) to conclude that there has been a significant decrease in FTR in 6,185,032 elderly patients undergoing cardiac surgeries from 2000 to 2018. The decrease was due to targeted quality metrics and care bundles for complications such as pneumonia and gastrointestinal bleeding. Incidence and trends were adjusted in FTR for groups based on patient information.

Our study is encouraged by studies like the ones mentioned above to introduce advanced ML techniques utilizing both static and intra-operative time-series data to develop a more accurate predictive model for FTR. The inclusion of intra-operative data makes sure that the model is more comprehensive than using data only from the STS database and reflective of critical intra-operative events that affect post operative outcomes. Since the dataset is limited, a technique had to be considered that captured complex, non-linear interactions in the data while preventing overfitting. Also, since we are working with intra-operative factors, we needed a technique that could work with time series data and chose Long Short-Term Memory (LSTM) networks. By employing a combination of fully connected neural networks (FCNN) and Long Short-Term Memory (LSTM) networks, our approach addresses the complexity of the data structures and captures the temporal dynamics essential for timely predictions. Deep learning provides a more sophisticated understanding of different variables and modalities. Our approach combines all target columns to predict the possibility of any adverse event among them. To make sure we got the best results, we decided to use the most useful features by computing the feature importances using XGBoost for the static data and performed an extensive ablation study for the time series data. We achieved an average c-index score of 0.92 by following this approach.

We are focusing in identifying failure to rescue complications, which if intervened upon in the early post operative course may reduce the mortality rate after cardiothoracic surgery. Similarly, by providing a transparent methodology and rigorous performance evaluation, our research contributes to the ongoing efforts to enhance patient safety and quality of care in cardiac surgery settings. This work not only advances the field of surgical risk prediction but also sets a foundation for developing actionable insights that can be operationalized across various healthcare contexts.

## 2 Methods

### 2.1 Data Source

The Society of Thoracic Surgeons (STS) Adult Cardiac Surgery Database (ACSD) (^5^) was utilized for this study, focusing on cases from Maine Medical Center over a period from September 2022 to April 2024. This database includes detailed time series intraoperative information for each patient. 1,470 patients were included in the dataset. All personal identifiers and private health information (PHI) were removed to protect patient confidentiality. Data were managed according to the ethical standards for medical research involving human subjects, ensuring the reproducibility and portability of our research across other institutions using similar datasets.

### 2.2 Cohort

The cohort for this study was defined to identify patients likely to develop one or more of eleven specific adverse events post-cardiac surgery, as endorsed by STS National Quality Forum (NQF) measures. These adverse events are mentioned in Table 1 below:

**Table 1:** Counts of target variables in the dataset.

| Target Name | Positive Counts |
| --- | --- |
| Post-Op-Aortic Intervention [6774: CAortReint] | 2 |
| Post-Op-ReOp for Valvular Dysfunction [6765: COpReVlv] | 4 |
| Post-Op-Deep Sternal [6700: DeepSternInf] | 5 |
| Post-Op-Unplanned Coronary Artery Intervention [6771: CReintMI] | 7 |
| Post-Op-ReOp Other Cardiac Reasons [6778: COpReOth] | 21 |
| Post-Op-Neuro-Stroke Perm [6810: CNStrokP] | 26 |
| Post-Op-Renal-Renal Failure [6870: CRenFail] | 40 |
| Post-Op-ReOp Bleeding/Tamponade [6755: COpReBld] | 47 |
| Status at Hospital Discharge [7007: DischMortStat] | 48 |
| Mort-Status at 30 Days After Surgery (either discharged or in-hospital) [7001: Mt30Stat] | 52 |
| Post-Op-Pulm-Vent Prolonged [6835: CPVntLng] | 108 |
| **Total Events** | **171** |

From the 1,470-patient dataset from the MMC STS registry, we identified 171 patients who developed at least one of the adverse events. This group formed our case set, as shown in Table 1. The control set comprised 171 randomly selected patients from the dataset who did not develop any of the adverse events.

### 2.3 Traditional Risk Score Model

In previous studies, traditional risk models such as those developed by Kurlansky et al. [2021] (^9^) utilized a select set of variables—prolonged ventilation, stroke, reoperation, and renal failure - to differentiate between failure to rescue (FTR) cases. Despite achieving a c-statistic of 0.81, the lack of additional performance metrics (sensitivity, specificity, PPV, NPV, and 95% CI) limits the comprehensive evaluation of these models’ effectiveness. Further limitations are noted in the works of Shahian et al. [2018] (^6^) and O’Brien et al. [2018] (^5^), who used the STS Adult Cardiac Surgery Database to develop risk scores for various outcomes but did not fully disclose the functional forms of the risk estimators, nor did they include intraoperative information, which hampers replicability and thorough assessment.

### 2.4 Current Deep Learning Model

We explored the integration of static and time-series data using a FCNN + LSTM network to capture dynamic changes in patient conditions. This model emphasizes the importance of dynamic data in predicting patient outcomes more accurately than traditional static models.

The incorporation of multi-modal late fusion is crucial for enhancing the performance of our predictive model (^2^). By integrating static data from the Society of Thoracic Surgeons (STS) database with time-series data from the operating room, our model leverages the strengths of both data types. This fusion approach allows for a more comprehensive understanding of patient conditions, leading to better predictive accuracy.

Similarly, our FCNN + LSTM model benefits from the fusion of static and dynamic patient data, providing a more complete and accurate representation of patient conditions. This multi-modal approach is essential for optimizing the predictive capabilities of our model, ultimately leading to better clinical decision-making and outcomes. We also captured the results by using FCNN over the static data alone and the time series alone using LSTM. Additionally, from the mixed model of FCNN + LSTM (Figure 1) where we used both static and time series data, we performed an ablation study by removing each time series feature one at a time and recording the results, as seen in the <u>sub-</u> section 4.3 and detailed description in <u>supplemental 8</u>. This ablation study helped identify the most critical time-series features that contribute to the model’s performance, highlighting the significance of each feature in predicting patient outcomes.

**Figure 1:**
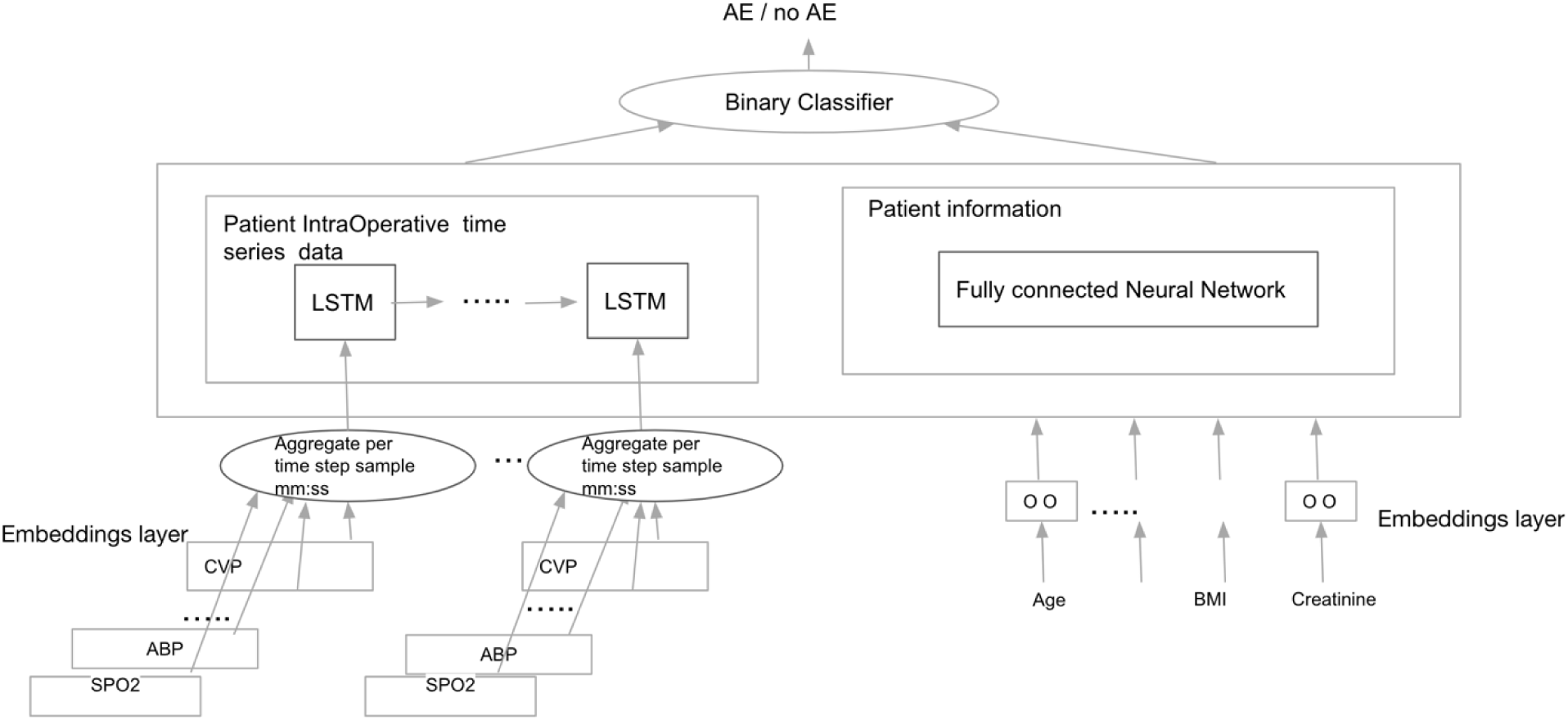
Architecture of Fully Connected and LSTM Networks

## 3 Statistical Analysis

We performed statistical analysis to compare demographic and clinical factors between the control cohort and cases cohort. The analysis was conducted using independent t-tests for continuous variables (Patient Age, Calculated BMI) to compare the means between the control and cases cohorts. Chi-square tests were used for categorical variables (Sex, Race/Ethnicity, ASCVD, Cardiovascular Disease, CAD, Heart Failure, Hypertension, Diabetes, Current Smokers) when expected frequencies were sufficiently large.

Fisher’s Exact Test was used for categorical variables when expected frequencies were small (less than 5). Table 2 presents the calculated p-values for each characteristic, indicating the significance of differences observed between the two cohorts.

**Table 2:**
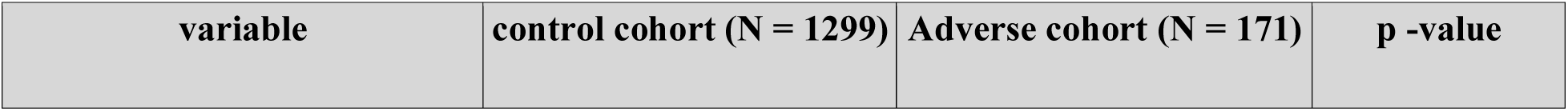
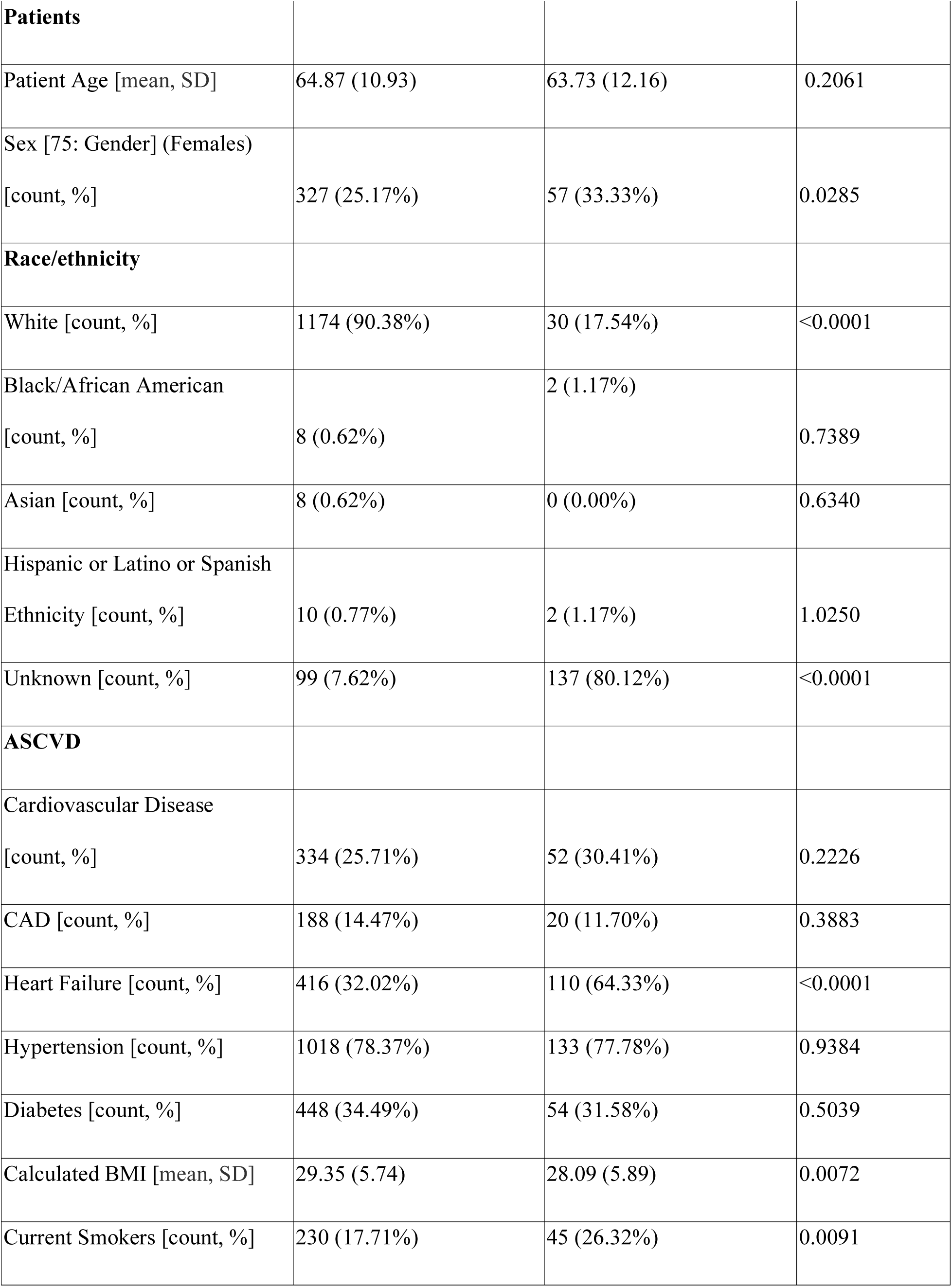
Distributions of characteristics of patients.

## 4 Results

After creating the cohort of 171 patients with at least one of the adverse events and the control group of 171 randomly selected patients that did not have any adverse events. The FCNN + LSTM model is validated with stratified five - fold cross validation and outperformed the best reported model for similar task by achieving a Mean AUC of 0.93, Mean Sensitivity of 0.84, Mean Specificity of 0.9, Mean PPV of 0.89, Mean NPV of 0.85 for balanced classes as shown in the last row of Table 3. The graphical representation of the performance of each fold can be seen in Figure 2.

**Figure 2:**
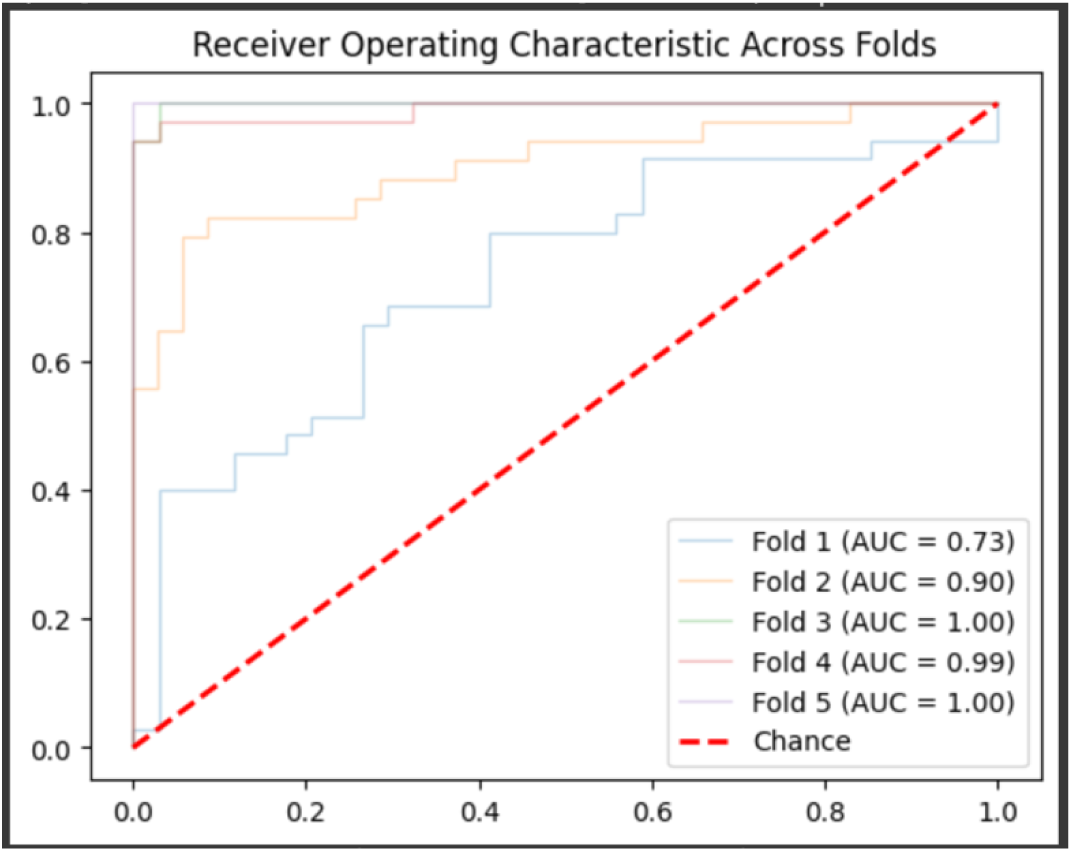
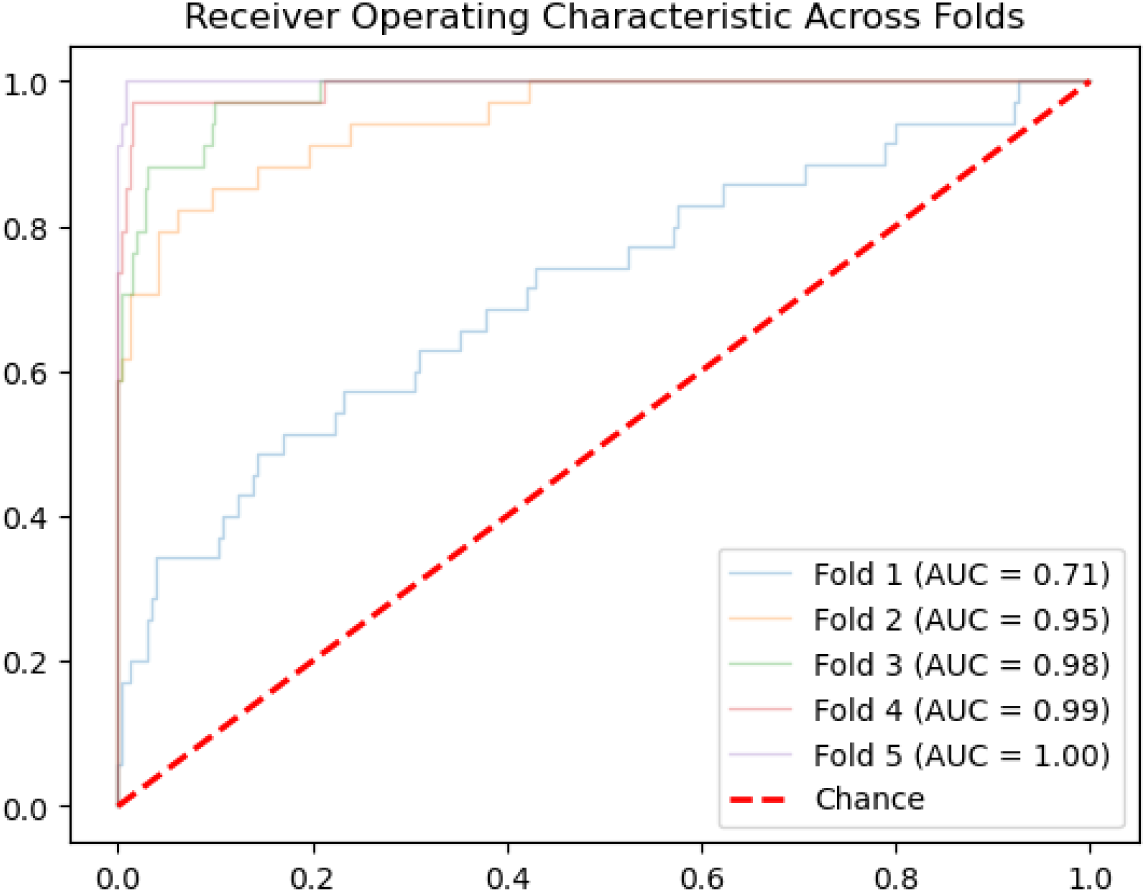
Area under the curve for the FC + LSTM model with balanced classes

**Table 3:** Performance of the model across all 5 folds in balanced dataset.

| Fold | AUC | 95% CI | Sensitivity | Specificity | PPV | NPV |
| --- | --- | --- | --- | --- | --- | --- |
| 1 | 0.7303 | 0.6030 -<br>0.8504 | 0.5714 | 0.7353 | 0.6897 | 0.6250 |
| 2 | 0.9025 | 0.8201 -<br>0.9687 | 0.8235 | 0.7714 | 0.7778 | 0.8182 |
| 3 | 0.9983 | 0.9913 -<br>1.0000 | 0.9118 | 1.0000 | 1.0000 | 0.9189 |
| 4 | 0.9896 | 0.9614 -<br>1.0000 | 0.8824 | 1.0000 | 1.0000 | 0.8947 |
| 5 | 1.0000 | 1.0000 -<br>1.0000 | 1.0000 | 0.9706 | 0.9714 | 1.0000 |
| average | $0.9210 \pm 0.1165$ | 0.8751 -<br>0.9638 | 0.8378, 95%<br>CI:<br>(0.69,0.98) | 0.8955, 95%<br>CI:<br>(0.78,1.01) | 0.8878, 95%<br>CI:<br>(0.76,1.01) | 0.8514, 95%<br>CI:<br>(0.73,0.98) |

### 4.1 Results after using only static features with fc model on balanced data

The scores of individual folds while working with static features in balanced data are in Table 4 and the graphical representation is in Figure 3.

**Figure 3:**
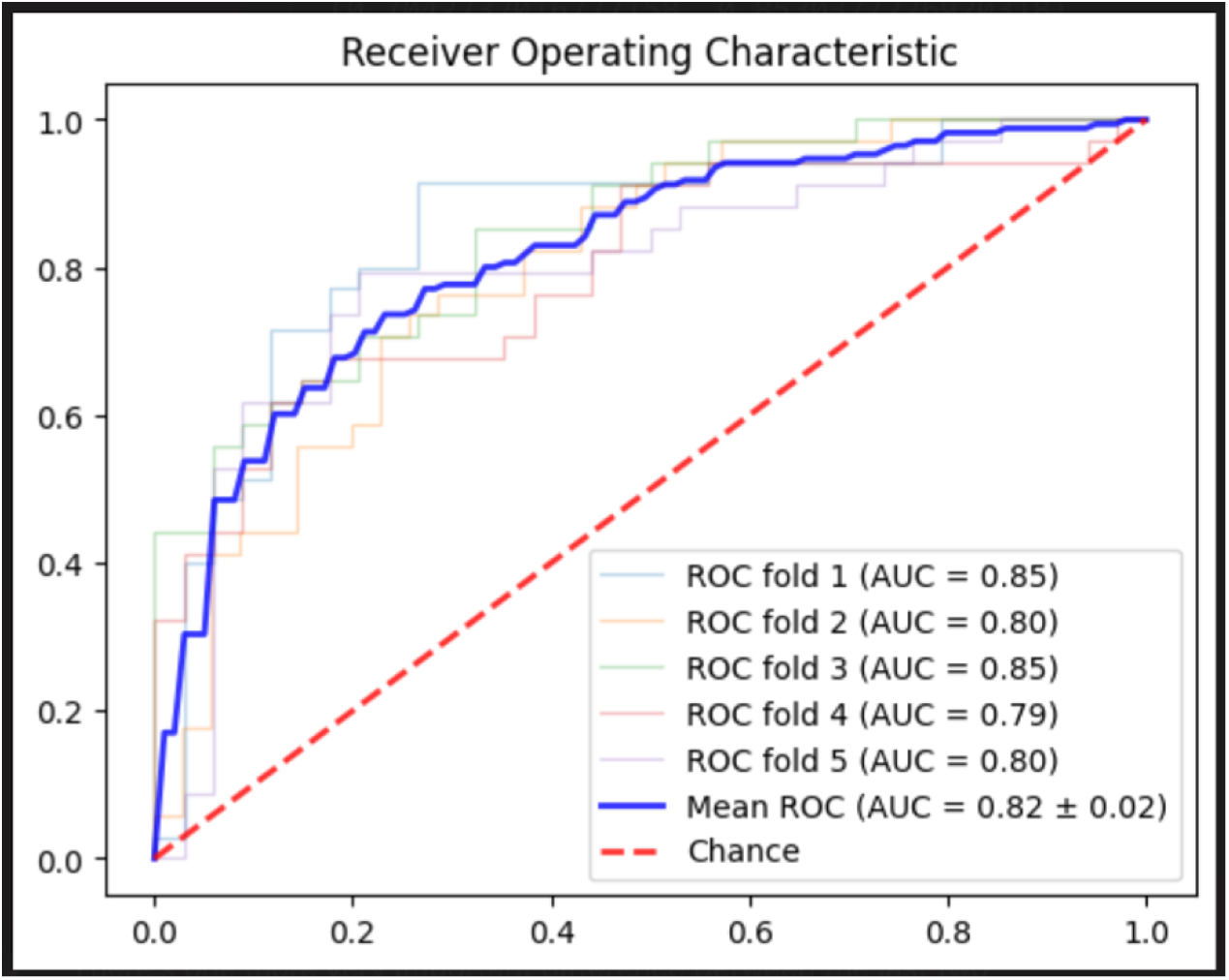
Area under the curve for the FC model with balanced classes

**Table 4:** Performance of the FC model across all 5 folds in balanced dataset.

| Fold | AUC | 95% CI | Sensitivity | Specificity | PPV | NPV |
| --- | --- | --- | --- | --- | --- | --- |
| 1 | 0.8630 | 0.7711 -<br>0.9436 | 0.8286 | 0.7353 | 0.7632 | 0.8065 |
| 2 | 0.7471 | 0.6204 -<br>0.8530 | 0.6471 | 0.7429 | 0.7097 | 0.6842 |
| 3 | 0.7889 | 0.6831 -<br>0.8874 | 0.6471 | 0.8235 | 0.7857 | 0.7000 |
| 4 | 0.8391 | 0.7368 -<br>0.9264 | 0.6765 | 0.9118 | 0.8846 | 0.7381 |
| 5 | 0.6920 | 0.5597 -<br>0.8187 | 0.5588 | 0.7059 | 0.6552 | 0.6154 |
| average | 0.8183 ±<br>0.0249 | 0.67422 -<br>0.88582 | 0.7192, 95%<br>CI:<br>(0.65,0.78) | 0.7787, 95%<br>CI:<br>(0.71,0.85) | 0.7696, 95%<br>CI:<br>(0.73,0.82) | 0.7385, 95%<br>CI:<br>(0.71,0.77) |

### 4.2 Results after using only time series features with the lstm model on balanced data

The scores of individual folds while working with time series features in balanced data are in Table 5 and the graphical representation is in Figure 4.

**Figure 4:**
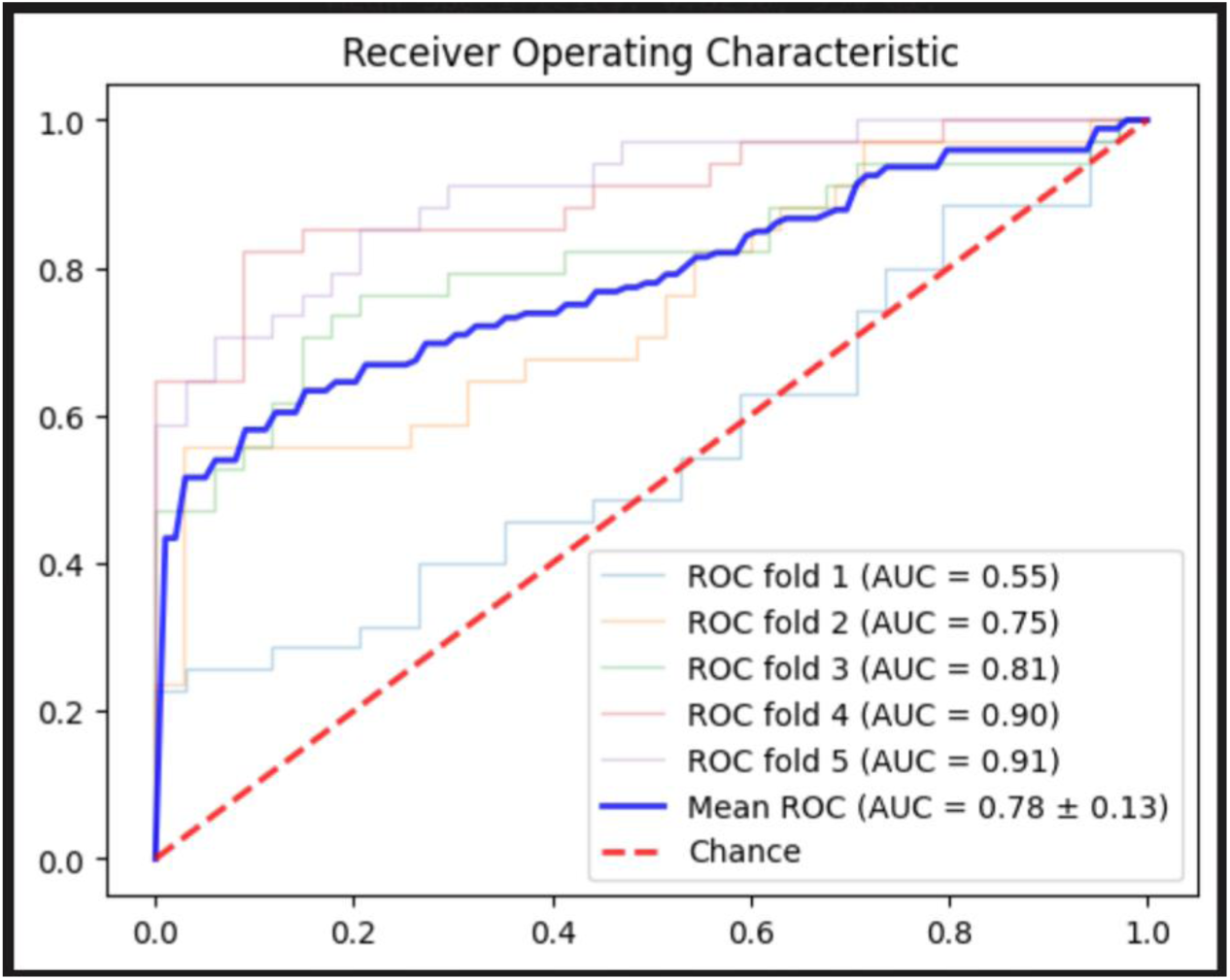
Area under the curve for the LSTM model with balanced classes

**Table 5:** Performance of the LSTM model across all 5 folds in balanced dataset.

| Fold | AUC | 95% CI | Sensitivity | Specificity | PPV | NPV |
| --- | --- | --- | --- | --- | --- | --- |
| 1 | 0.7874 | 0.6700 -<br>0.8795 | 0.5143 | 0.9118 | 0.8571 | 0.6458 |
| 2 | 0.8798 | 0.7895 -<br>0.9561 | 0.7647 | 0.8857 | 0.8667 | 0.7949 |
| 3 | 0.9464 | 0.8787 -<br>0.9922 | 0.7059 | 0.9706 | 0.9600 | 0.7674 |
| 4 | 0.9066 | 0.8300 -<br>0.9729 | 0.6471 | 0.9706 | 0.9565 | 0.7333 |
| 5 | 0.9619 | 0.9134 -<br>0.9947 | 0.9118 | 0.8529 | 0.8611 | 0.9062 |
| average | 0.7837 ±<br>0.1281 | 0.8163 –<br>0.9591 | 0.6160, 95%<br>CI:<br>(0.41,0.82) | 0.8250, 95%<br>CI:<br>(0.77,0.88) | 0.7695, 95%<br>CI:<br>(0.71,0.83) | 0.7026, 95%<br>CI:<br>(0.59,0.82) |

### 4.3 Ablation study

In our ablation study analysis, we have removed one time series feature at a time to check the performance fluctuations in the model, different features exhibited varying impacts on key metrics such as AUC, sensitivity, specificity, positive predictive value (PPV), and negative predictive value (NPV). Starting with the AUC, the exclusion of the Pulse rate as a feature contributed to the most substantial increase, improving the AUC by 0.0301 or 3%. Conversely, the removal of all Pulmonary Artery (PA) values resulted in a notable decrease in AUC by 0.1991, a 22% reduction. The only other feature that decreased the AUC marginally by 0.0066 (1%), was the Electrocardiogram Heart Rate (ECG HR).

Regarding sensitivity, the maximum increase was observed when ETCO2 (end-tidal carbon dioxide) was excluded, enhancing sensitivity by 0.0467 or 3%. On the other hand, removing all PA values led to the largest decrease in sensitivity by 0.1912 or 23%. Another notable decline was when the bypass feature was removed, which decreased sensitivity by 0.0287 or 3%.

For specificity, the exclusion of Pulse rate again showed the most significant positive impact, increasing specificity by 0.0526 or 6%. In contrast, the removal of all Arterial (ART) values led to the largest decrease in specificity, a substantial reduction of 0.2637 or 29%. Only the removal of the feature ST generic V slightly decreased specificity by 0.0002 or less than 1%.

In terms of PPV, like specificity, exclusion of Pulse rate provided the most significant boost, increasing PPV by 0.0539 or 6%. The removal of all ART values again caused the most significant decrease, lowering PPV by 0.2118 or 24%. The removal of ST generic V also slightly decreased PPV by 0.0021. Lastly, the NPV saw its greatest increase with the exclusion of ETCO2, which improved NPV by 0.0438 or 5%. The largest decline in NPV occurred with the removal of all PA values, which decreased it by 0.1909 or 22%. Additionally, removing the bypass feature led to a minor decrease in NPV by 0.0047 or 1%.

As observed from the above comparisons, we can say that there is always a decrease in scores when the entirety of the ABP, AO, PA or ART values are removed. Using just the time series or static variables with the LSTM and FC models respectively also always results in a reduction of scores. The model combing both the models gives us better results.

During the ablation study, some features when removed, resulted in better performance. The ablation study, as mentioned, only deals with time series features. Based on the results, the scores improve when the features Pulse Rate, ETC02, PA Sys are removed from the time series features.

Results of ablation study can be found in Table 6 below and detailed description in supplemental 8.

**Table 6:**
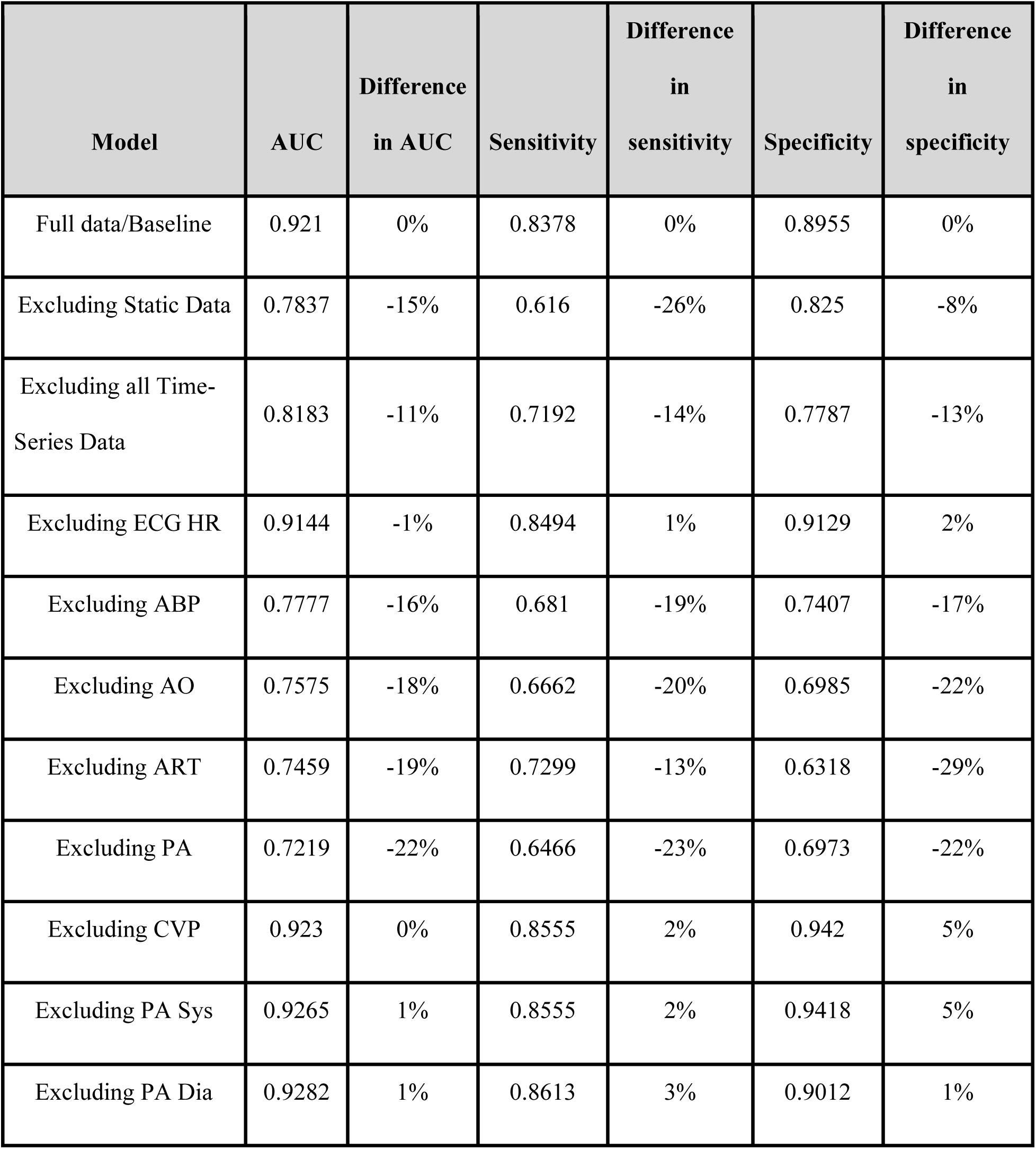
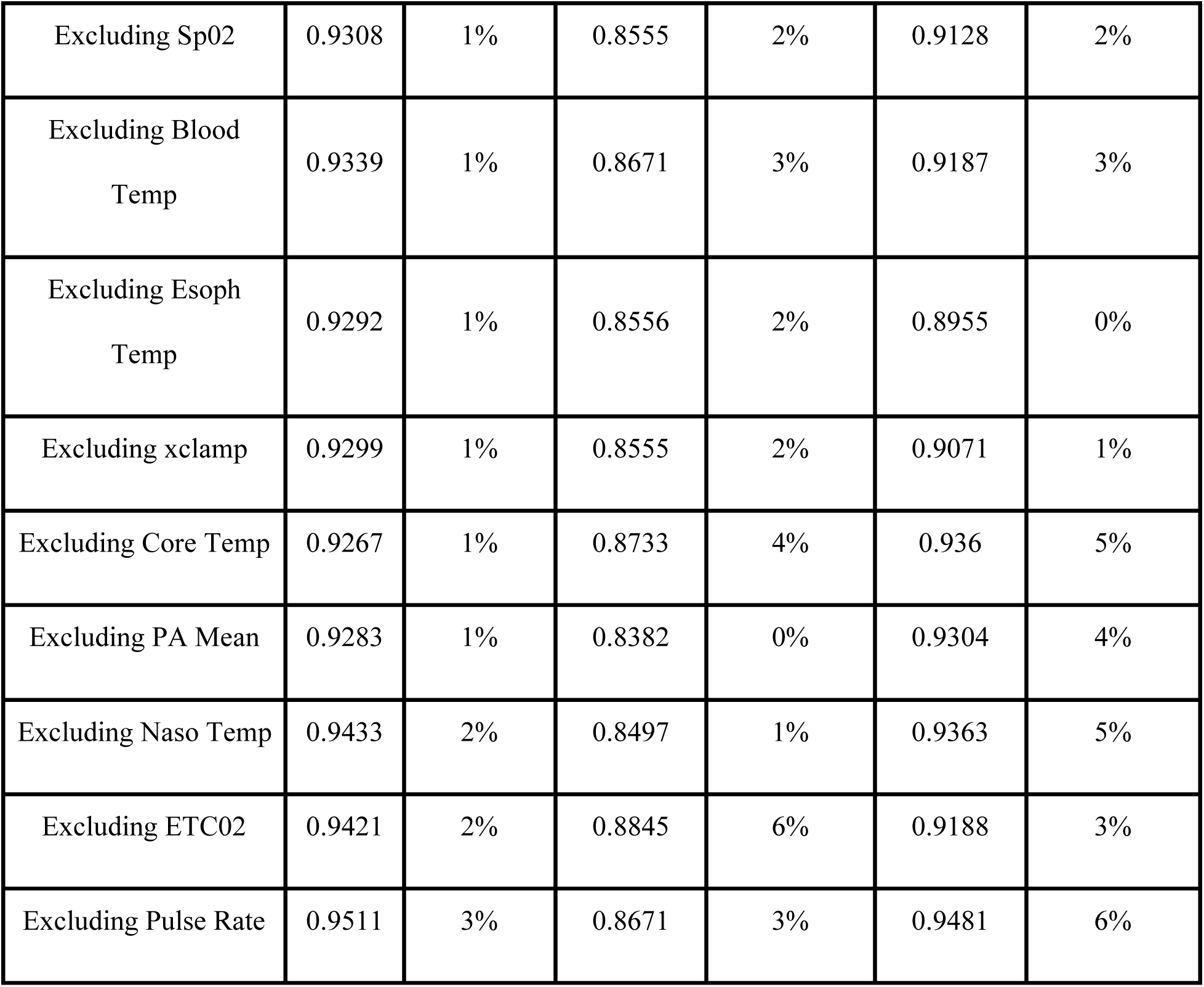
Results of ablation study.

### 4.4 Results with only the most impactful variables from the ablation study

As noticed in the ablation study above, certain variables, namely AO, ABP, PA, ART, and ECG HR, were found to be important as their removal reduced the model’s overall performance. We tested a model incorporating only these few variables along with all the other static variables using the same FC + LSTM architecture. The results were quite impressive; the average AUC across all the Folds obtained from this model was 0.9392 ± 0.0848, while the average AUC across all the folds from the model with all the variables was 0.9210 ± 0.1165, the detailed results can be seen in the Figure 5 and Table 7 below.

**Figure 5:**
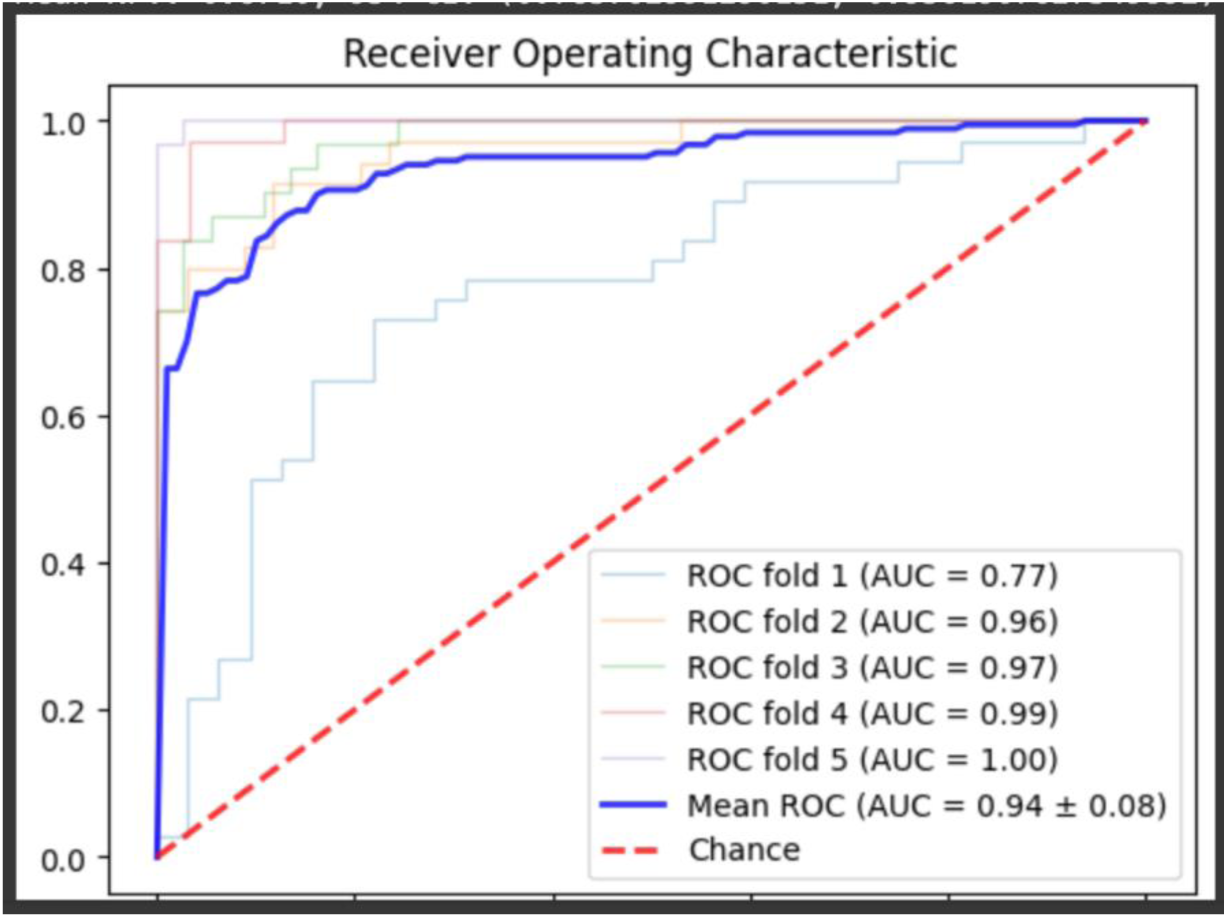
Area under the curve for the FC + LSTM model with only impactful time series features

**Table 7:**
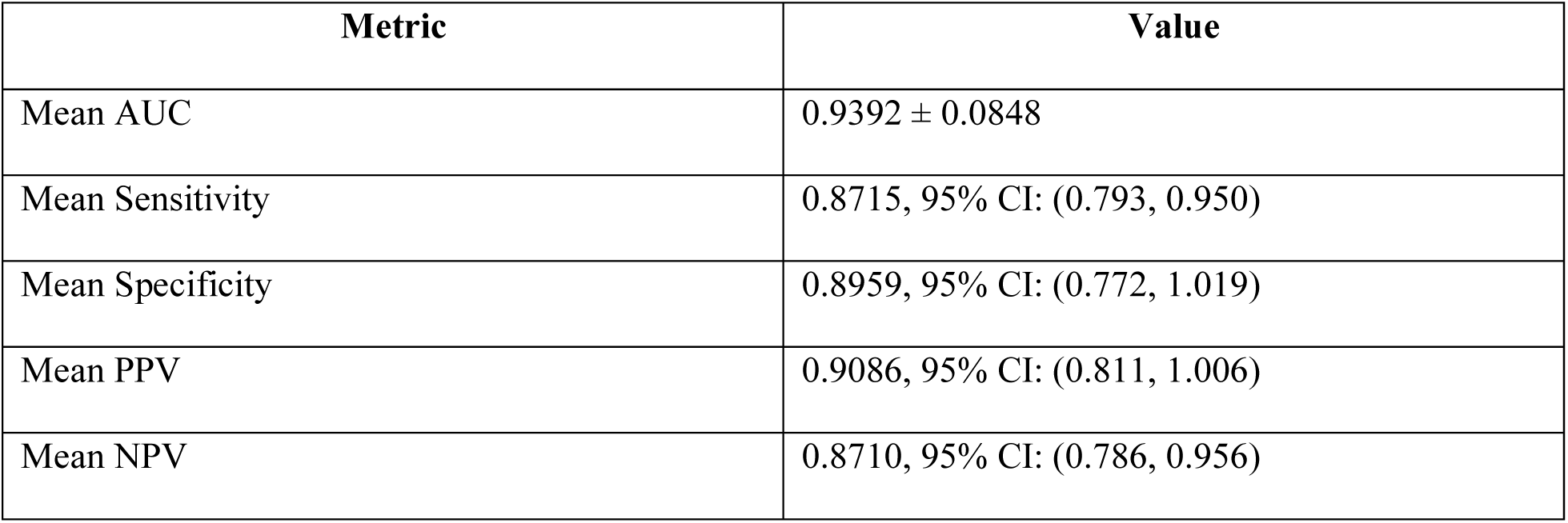
Results for FC + LSTM model with only impactful time series features.

| Metric | Value |
| --- | --- |
| Mean AUC | 0.9392 ± 0.0848 |
| Mean Sensitivity | 0.8715, 95% CI: (0.793, 0.950) |
| Mean Specificity | 0.8959, 95% CI: (0.772, 1.019) |
| Mean PPV | 0.9086, 95% CI: (0.811, 1.006) |
| Mean NPV | 0.8710, 95% CI: (0.786, 0.956) |

We tried to pull the feature importances using the static variables through XGBoost model. The top 20 static variables that most contributed to outcomes are stated in the Figure 6 while their absolute SHAP values are represented in the Figure 7. We had an initial assumption that ‘sex’ of the patient would also be among the most important variables contributing for the prediction, it is found that ‘sex’ is the 37^th^ among the feature importances.

**Figure 6:**
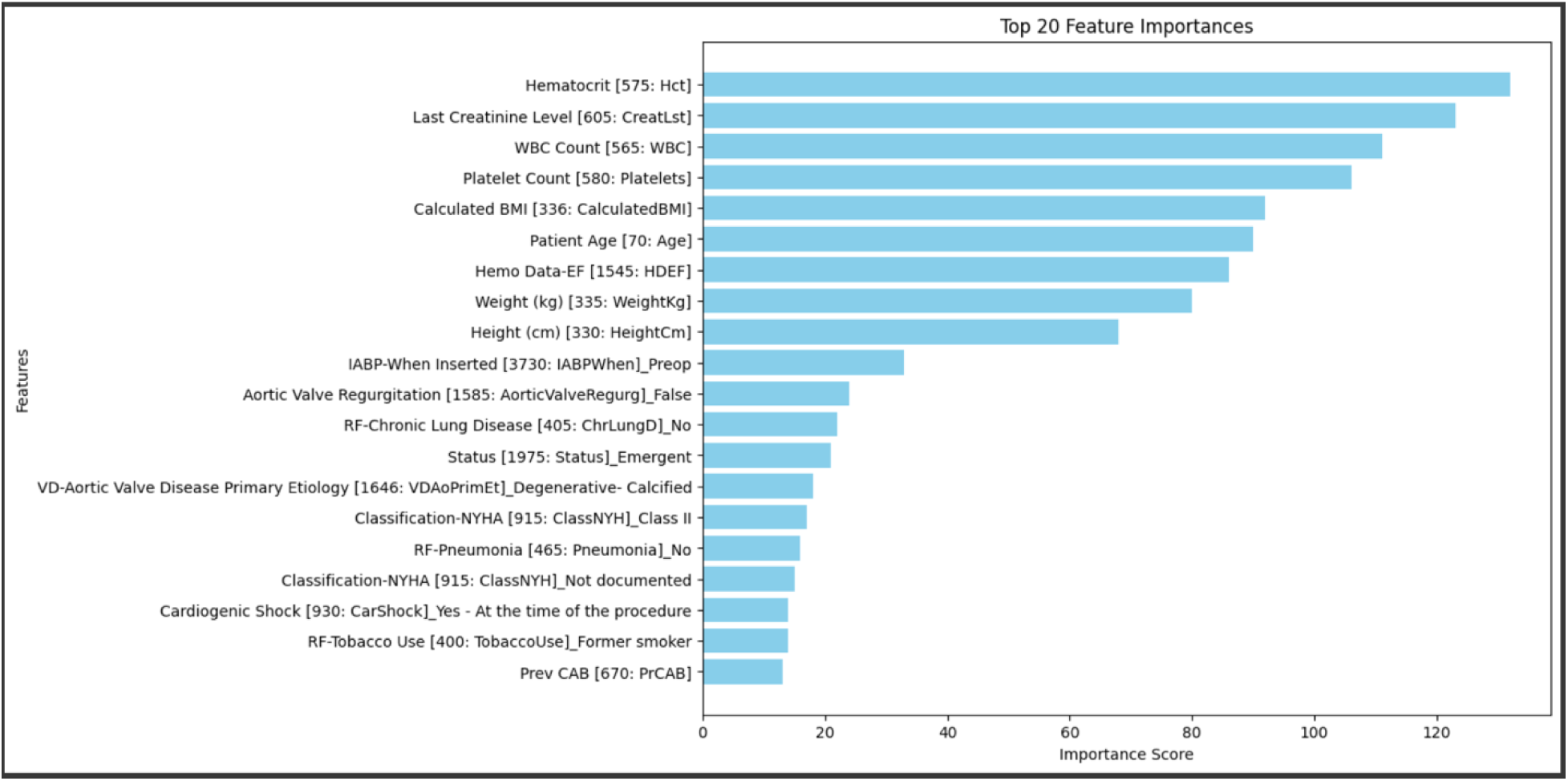
Top 20 most predictive features in static variables according to XGBoost

**Figure 7:**
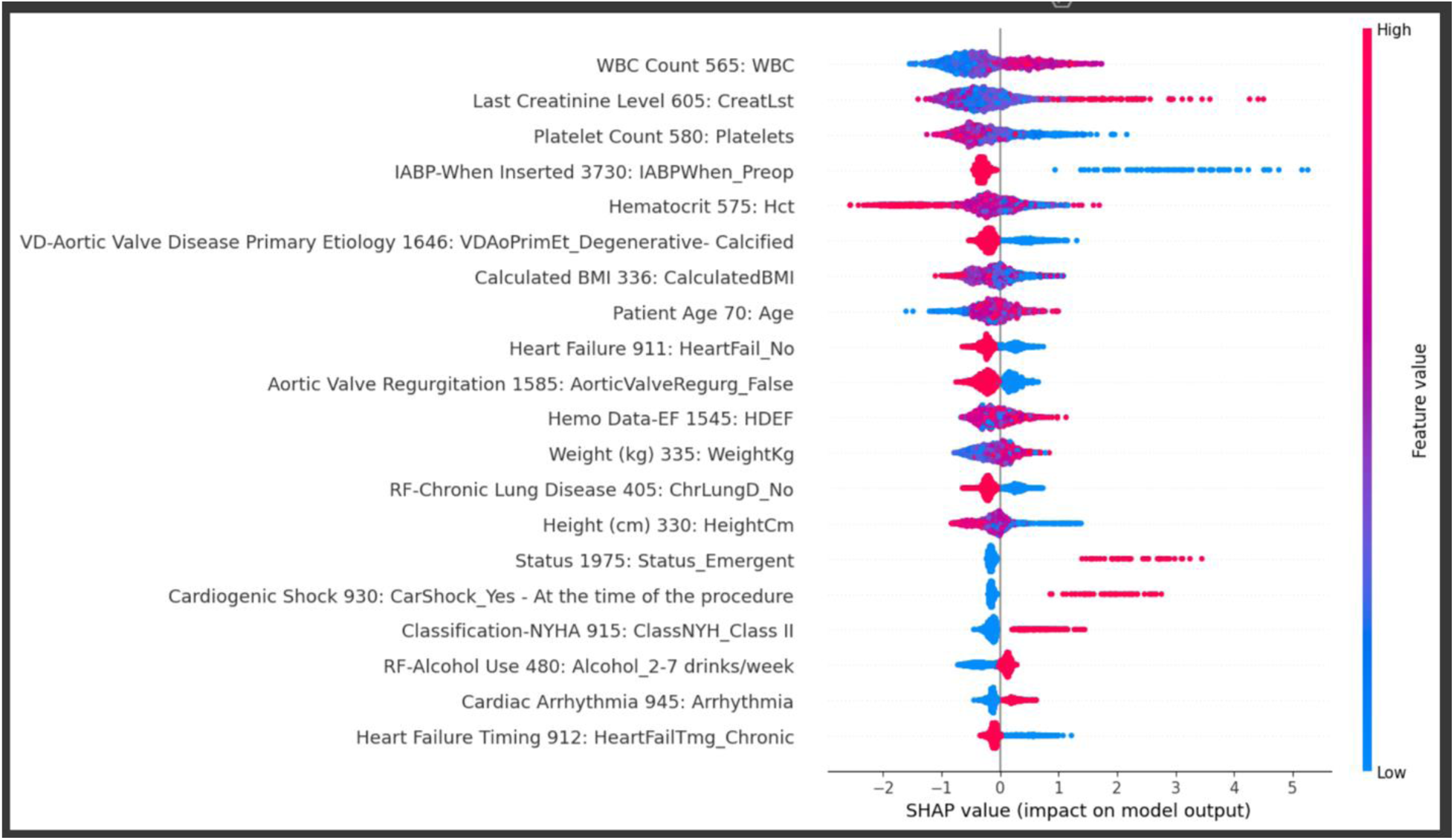
Absolute SHAP Values for Top 20 Predictive Features in the XGBoost Model

SHAP values are used to understand how each feature affects a model. An importance is attached to each feature and the magnitude of this importance value indicates the intensity of its effect on the model. Each dot in the above plot represents a row in the dataset.

The colors indicate the high and low values of the features. Categorical features are converted into dummy variables through one-hot encoding. Therefore, ones in these binary dummy features represent high values in the corresponding categorical variables. The magnitude of the positive or negative impact of each feature is determined by the position of the dots relative to zero, with dots on one side indicating a positive impact and dots on the other side indicating a negative impact.

To understand this better we can consider the example of the “IABP-When Inserted 3730” feature. It can very clearly be seen that the low values of this feature have a tangible positive impact on the model while the high values have a comparatively smaller negative impact. The same understanding is followed by all other features.

## 5 Discussions

In this investigation, we employed a hybrid FC + LSTM model to predict adverse events in patients undergoing cardiac surgery, utilizing a dataset of 1,470 patients from the Society of Thoracic Surgeons (STS) Adult Cardiac Surgery Database (ACSD) at Maine Medical Center between September 2022 and April 2024 (^5^). This model uniquely integrates static patient data, such as age and gender, processed through Fully Connected (FC) layers, with dynamic, time-series intraoperative data such as ABP Dia, ABP Mean, ABP Sys, CVP, and Pulse Rate processed via Long Short-Term Memory (LSTM) layers. The outputs from both data streams are then concatenated to perform binary classification. Validated using Fivefold cross-validation, our model demonstrated exceptional performance, notably achieving a Mean AUC of 0.93.

The advantage of our approach is further underscored by its robust performance across both balanced and imbalanced datasets, achieving high specificity and NPV in scenarios typical in clinical settings where class distribution can often be skewed. These results not only highlight the model’s precision but also its adaptability, making it highly suitable for real-world clinical applications. Our FC + LSTM model thus marks a significant advancement over traditional models, providing a deeper, more accurate assessment of patient risk profiles which could facilitate more timely and targeted interventions in cardiothoracic surgery. Future studies will focus on expanding the application of this model to other types of surgical data and further validating its effectiveness and generalizability across different healthcare environments. We achieved an average AUC of around 0.92 using our FC + LSTM model. Though Kurlansky et al. [2021] (^9^) used a different dataset, their purpose was similar, and by using the STS and time-series data, we obtained better results. In our previous random forest model, we used only static variables and achieved reasonable results. However, with the inclusion of time-series variables, we achieved even better results.

Before incorporating the time-series data, we created models using XGBoost, random forest, and decision trees with static data alone. The results of the four models are represented in Table 8, these results prove that the approach of using deep learning is better than traditional ML algorithms.

**Table 8:** Results of ML models with static data.

| **Model** | **AUC** | **Sensitivity** | **Specificity** | **PPV**<br>**(Precision)** | **NPV** | **95% CI for**<br>**AUC** |
| --- | --- | --- | --- | --- | --- | --- |
| Random<br>Forest | 0.79 | 0.72 | 0.8 | 0.79 | 0.73 | [0.70, 0.88] |
| Gradient<br>Boosting | 0.73 | 0.74 | 0.64 | 0.68 | 0.7 | [0.63, 0.83] |
| XGBoost | 0.69 | 0.68 | 0.68 | 0.69 | 0.67 | [0.58, 0.79] |
| Decision<br>Tree | 0.69 | 0.6 | 0.72 | 0.7 | 0.63 | [0.59, 0.79] |

## 6 Limitations

This model is based on a significantly small sample size of only 171 patients in the cases cohort. The previous models have been built with larger datasets. The model’s performance might be further enhanced with the addition of more patients as well as more variables such as intraoperative medication administration, intravenous fluid administration, blood product administration and urine output.

## 7 Conclusions and future work

We built an FC + LSTM model using the static and time series data. The static data has been processed with the FC neural network while the Time Series data has been processed with the LSTM neural network. We then concatenated both the layers and made a binary classifier with sigmoid activation function. We created a prediction model that out-performed our previous work that leveraged random forest with static variables, and existing logistic regression prediction models in cardiac surgery as our prediction model had a mean AUC of 0.93 for multiple adverse events in all the folds of data. We also observed that the model is robust to both classes balanced data and class imbalance data. Furthermore, the dynamic design of this FC + LSTM model is designed to improve with time and more data. This work combining the static data with time-evolving data in dynamic settings such as the operating room or intensive care units will drive real-time feedback to the care team with potential improvement in failure to rescue rates. In the future, as done by Ghanzouri et al. [2022] (^11^) we will add visualization to reflect the risk score for the care team to interpret in real time. We can then perform a user study similar to Ho et al. [2022] (^12^) to collect feedback from the care team on the efficacy of the risk score to help guide timely patient care.

This current approach combines 11 failures to rescue outcomes into a single target column which predicts if a cardiovascular event takes place post operatively or not. With complete datasets available for more patients, there is scope for creating a model which could predict individual failure to rescue outcomes.

Future work could explore the integration of digital twin models to enhance our predictive capabilities. By incorporating detailed fluid-structure interaction models(^39^), we could account for patient-specific arterial compliance and pulse transit time(^37^). This approach could provide more accurate hemodynamic parameters, potentially improving our ability to predict adverse events. Additionally, coupling 3D and 1D models(^38^) could offer a more comprehensive view of the cardiovascular system, allowing for better representation of complex structures like aneurysms or the venous system. These advancements could lead to more personalized risk assessments and treatment strategies for cardiothoracic surgery patients.

## Conflict of Interest

### Author Contributions

Conceptualization, S.A. and A.H.; methodology, S.A. R.K.; software, S.A. and K.R.; validation, S.A., K.R., A.H.; formal analysis, S.A. and K.R.; investigation, S.A., R.K. and D.K.; resources, S.A., K.R. A.H., R.K., J.A., D.S., R.W., F.M. and D.K.; data curation, K.R.; writing—original draft preparation, S.A. and K.R.; writing—review and editing, S.A., R.K., A.H., F.M., T.K, J.A., R.K., Q.J., D.S. R.W.; visualization, R.K; supervision, S.A.; project administration, S.A.; All authors have read and agreed to the published version of the manuscript.

## Funding

**AC COBRE**

## Data Availability

All data produced in the present study are available upon reasonable request to the authors

---

**Supplement 1.**
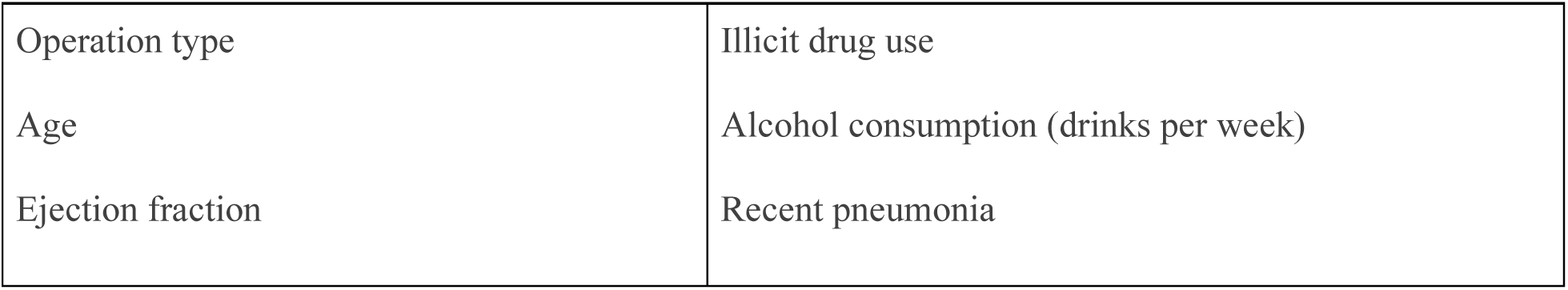
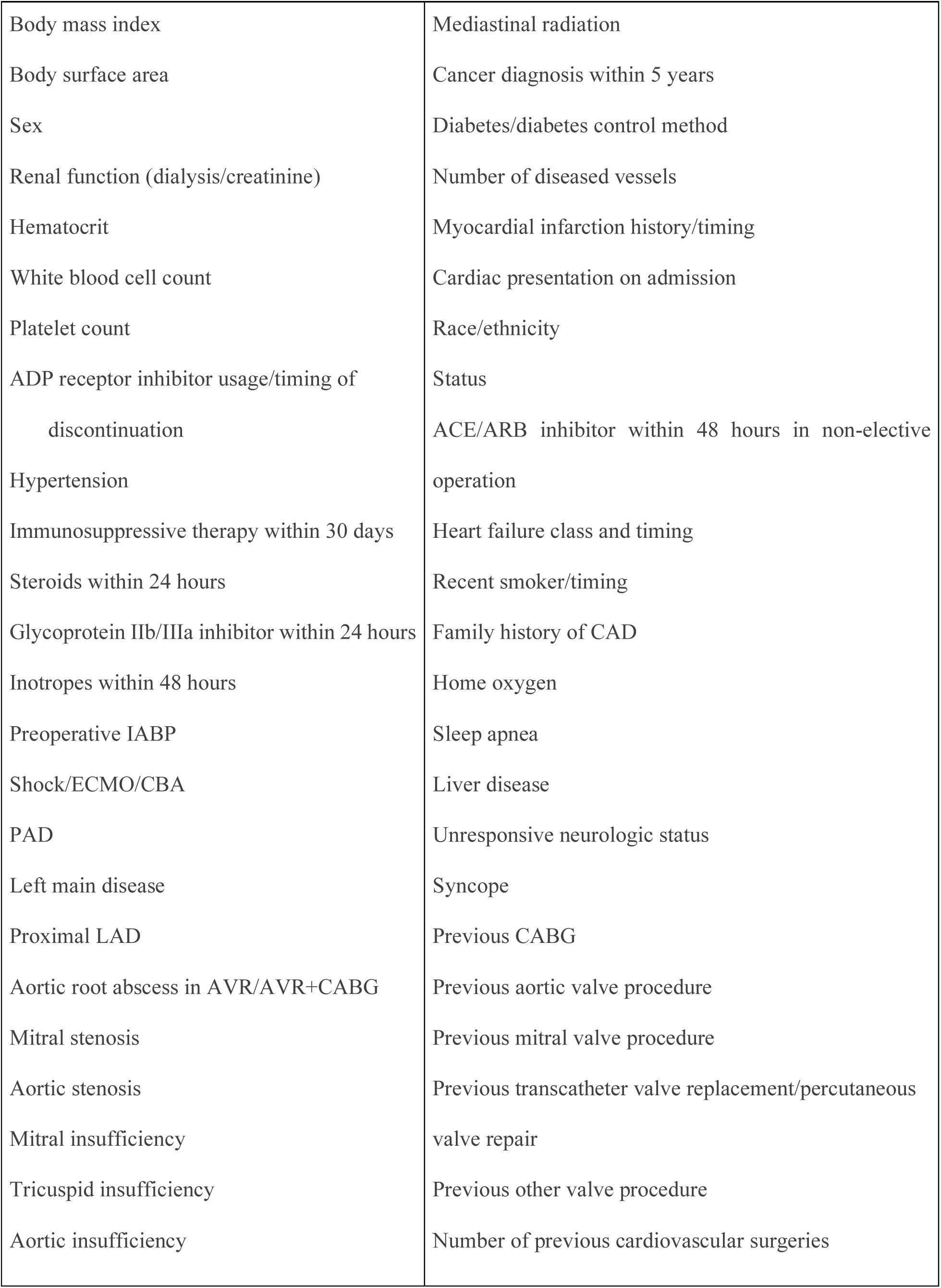
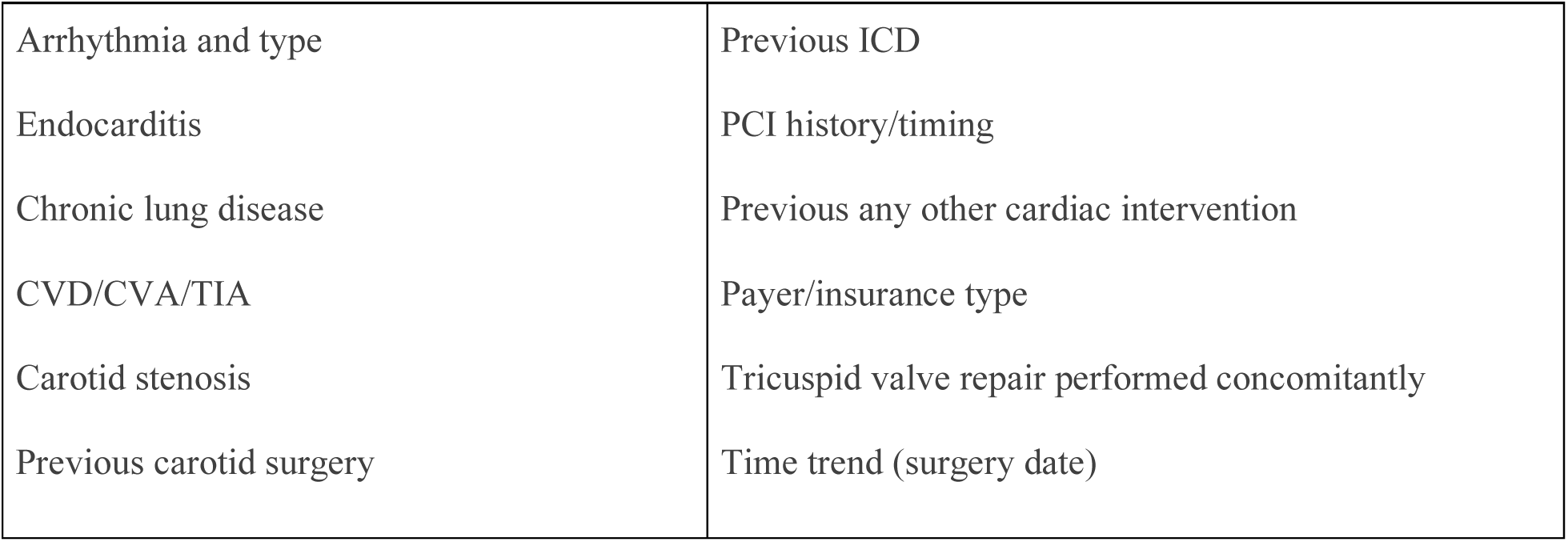
Static variables used to construct risk scores for these outcomes identified by O’Brien et al [2018]. (**^6^**). ACE = angiotensin-converting enzyme; ADP = adenosine diphosphate; ARB = angiotensin-receptor blocker; AVR = aortic valve replacement; CABG = coronary artery bypass grafting surgery; CAD = coronary artery disease; CBA = catheterization-based assist device; CVA = cerebrovascular accident; CVD = cardiovascular disease; ECMO = extracorporeal membrane oxygenation; IABP = intra-aortic balloon pump; ICD = implantable cardioverter-defibrillator; LAD = left anterior descending artery; PAD = peripheral arterial disease; PCI = percutaneous coronary intervention; TIA = transient ischemic attack.

**Supplement 2.** Static variables used in random forest algorithm.

|  |  |
| --- | --- |
| Patient Age | Pre Op Transient ischemic attack |
| Pre Op Ejection Fraction | Carotid stenosis |
| Body mass index | Illicit drug use |
| Height (cm) | Alcohol Use (drinks per week) |
| Weight (kg) | Recent pneumonia |
| Sex | Pre Op mediastinal radiation |
| Pre Op Dialysis | Cancer diagnosis within 5 years |
| Last Creatinine level | Pre Op Diabetes |
| Pre Op Hematocrit | diabetes control method |
| Pre Op White blood cell count | Number of diseased vessels |
| Pre Op Platelet count | Prior myocardial infarction |
| Pre Op Hypertension | Prior myocardial infarction timing |
| Immunosuppressive therapy w/in 30 days of procedure | Cardiac presentation on admission |
| Steroids within 24 hours of procedure | Race Documented |
| Glycoprotein IIb/IIIa inhibitor w/in 24 hours of procedure | Patient Race |
| Inotropes within 48 hours of procedure | Hispanic, Latino or Spanish ethnicity |
| IABP Insertion Timing | Operative Status |
| Pre Op Cardiogenic shock | Heart failure history |
| ECMO Insertion Timing | Heart failure timing |
| Temp assist Device Type (Open or Catheter) | Heart failure type |
| Catheter based assist device insertion timing | NYHA classification |
| Peripheral Arterial Disease | Pre Op Tobacco use |
| Left main coronary artery disease | Family history of Premature CAD |
| Proximal LAD percent stenosis | Home oxygen use |
| Aortic valve disease etiology | Pre Op Sleep apnea |
| Mitral valve stenosis | Pre Op Liver disease |
| Aortic valve stenosis | Pre Op Unresponsive neurologic status |
| Pre Op Mitral insufficiency grade | Pre Op Syncope |
| Pre Op Tricuspid insufficiency grade | Previous CABG procedure |
| Pre Op Aortic insufficiency grade | Previous valve procedure |
| Arrhythmia type | Previous valve procedure type |
| Pre Op Endocarditis | Previous other cardiac procedure type |
| Pre Op Chronic lung disease | Primary payer/insurance type |
| Pre Op Cerebrovascular disease | Tricuspid valve repair performed |
| CVA | Surgery date |

**Supplement 4.**
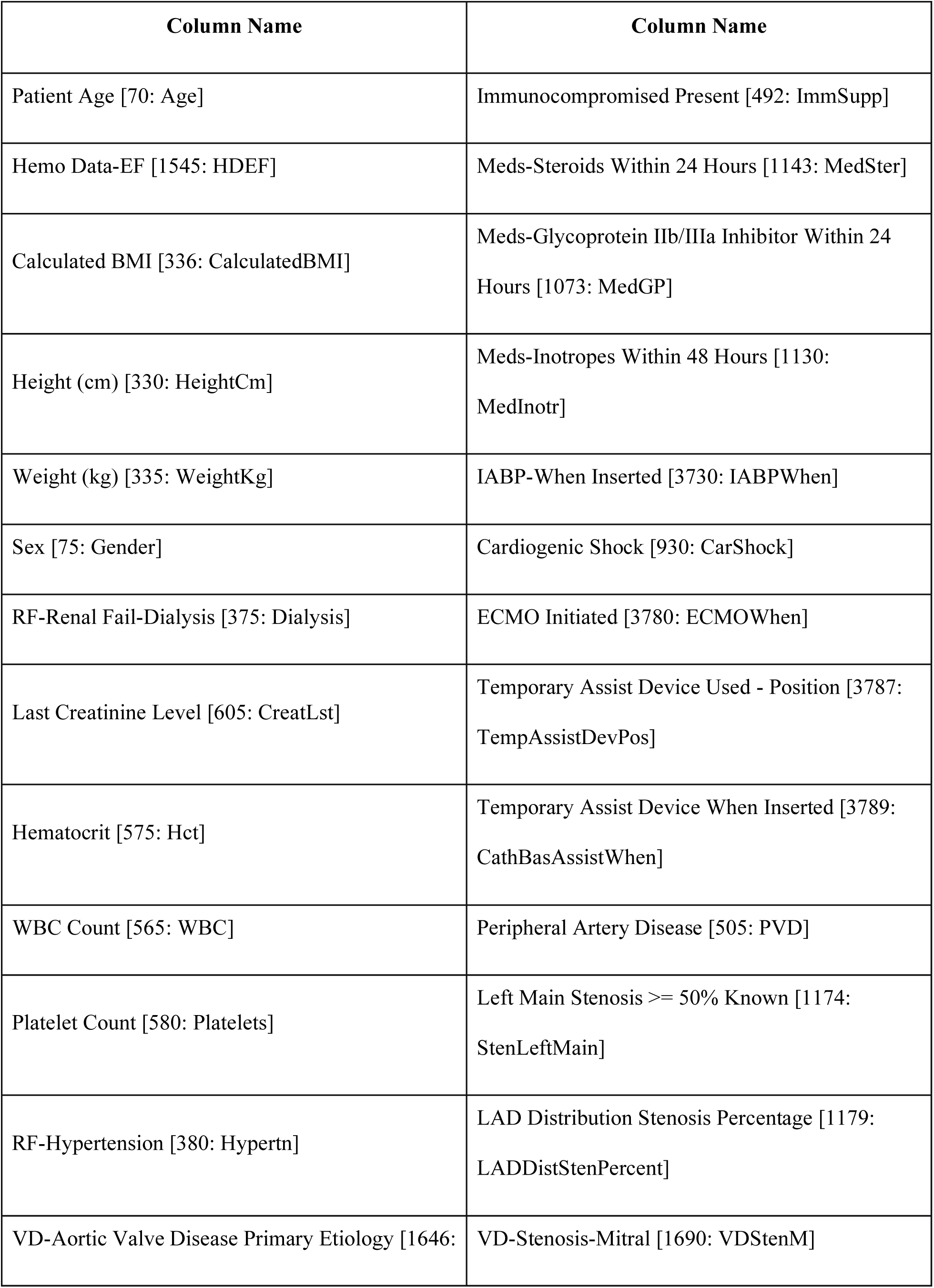
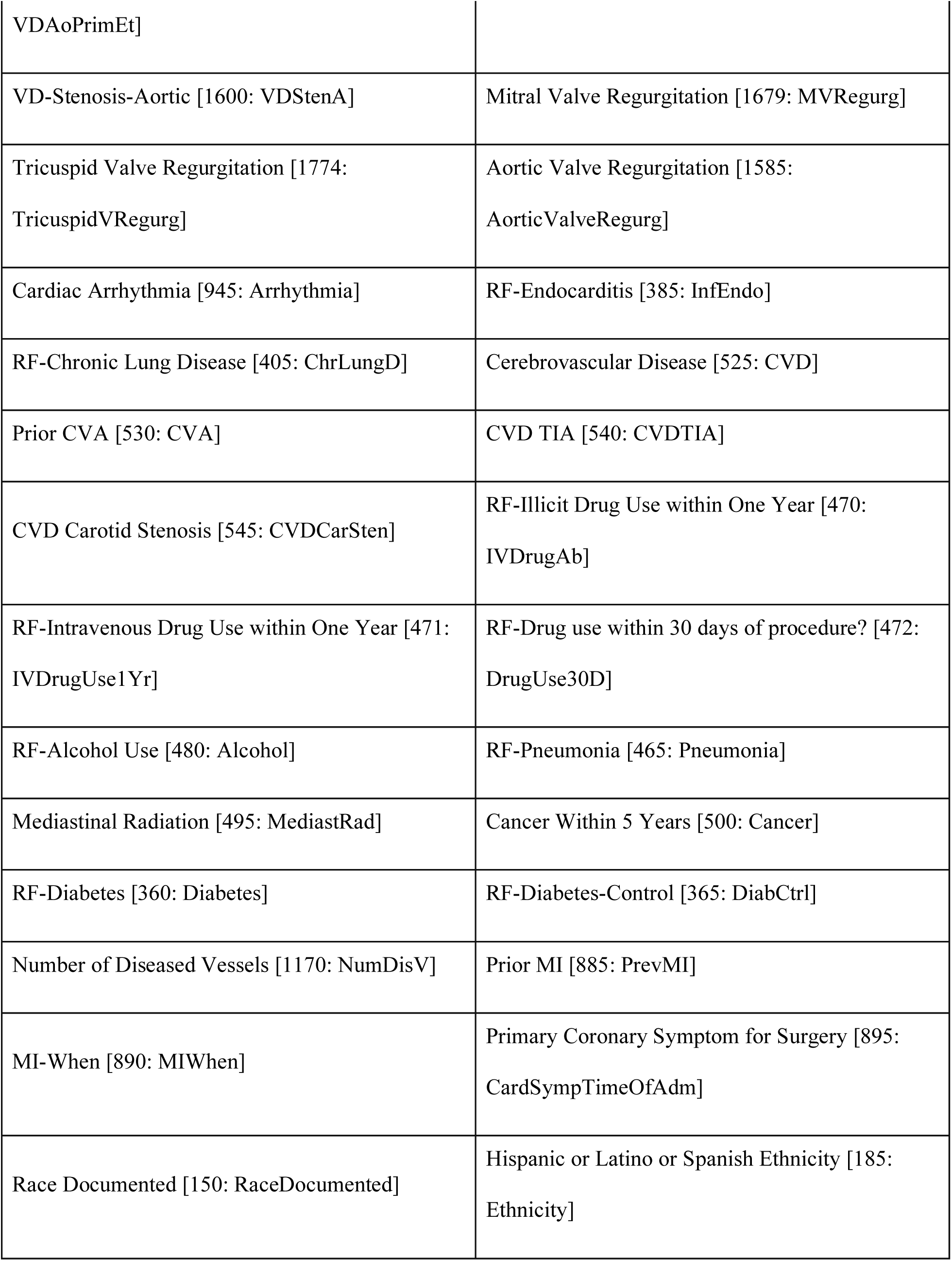
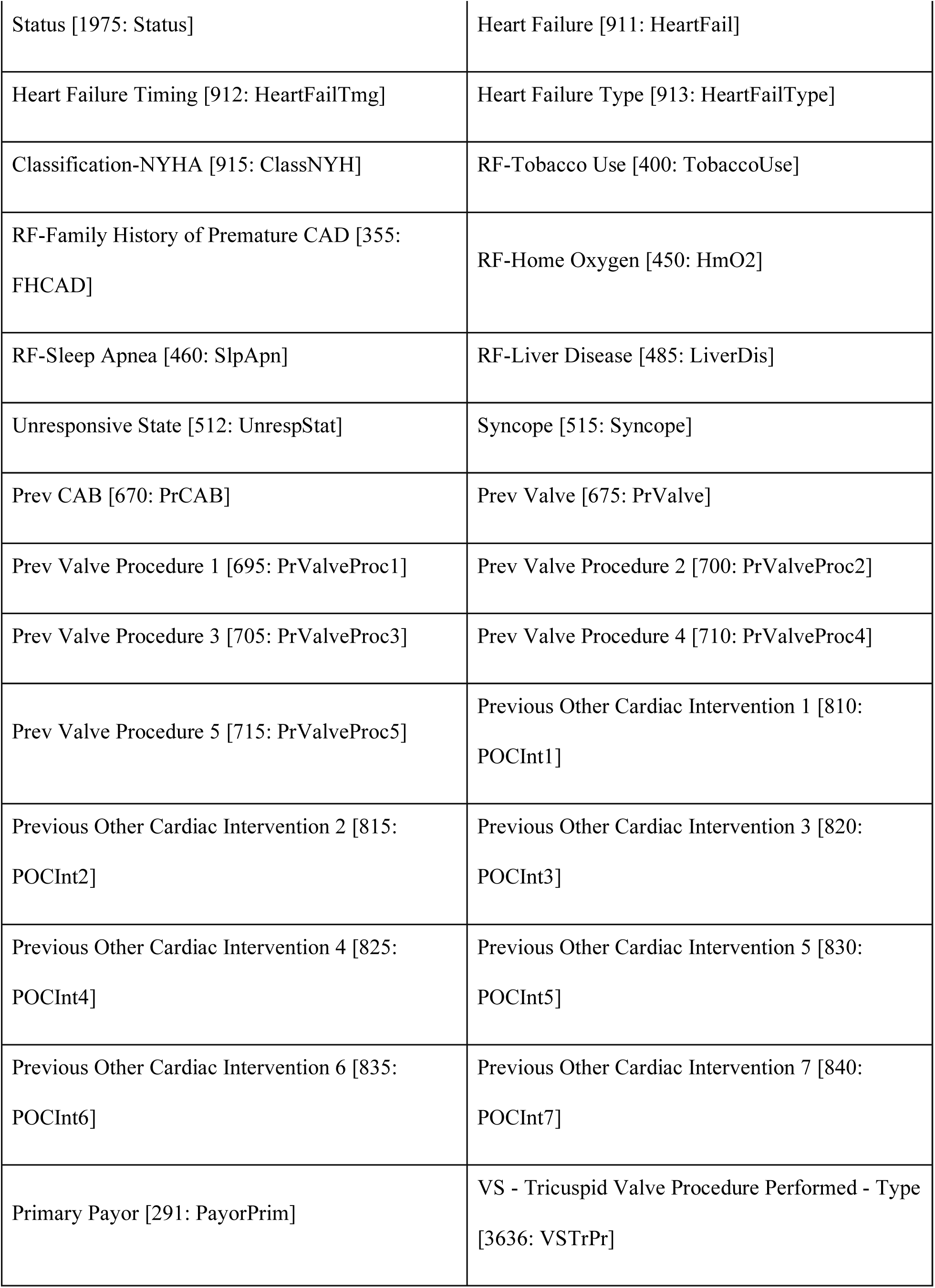
Static variables used in the FC + LSTM Model.

**Supplement 5.** Time Series variables used in the FC + LSTM Model.

| Column Name | Column Name |
| --- | --- |
| ABP Dia | ART Dia |
| ABP Mean | ART Mean |
| ABP Sys | ART Sys |
| AO Dia | CVP |
| AO Mean | ETCO2 |
| AO Sys | Core Temp |
| ECG HR | Esoph Temp |
| Naso Temp | PA Dia |
| PA Mean | PA Sys |
| Pulse Rate | SpO2 |
| ST generic II | ST generic V |
| Blood Temp | bypass |
| xclamp |  |

**Supplement 6.** Top 20 static variables with rank of importance for XGBoost.

| **Feature Rank** | **Column Name** |
| --- | --- |
| 1 | Hematocrit [575: Hct] |
| 2 | Last Creatinine Level [605: CreatLst] |
| 3 | WBC Count [565: WBC] |
| 4 | Platelet Count [580: Platelets] |
| 5 | Calculated BMI [336: CalculatedBMI] |
| 6 | Patient Age [70: Age] |
| 7 | Hemo Data-EF [1545: HDEF] |
| 8 | Weight (kg) [335: WeightKg] |
| 9 | Height (cm) [330: HeightCm] |
| 10 | IABP-When Inserted [3730: IABPWhen]_Preop |
| 11 | Aortic Valve Regurgitation [1585: AorticValveRegurg] |
| 12 | RF-Chronic Lung Disease [405: ChrLungD]_No |
| 13 | Status [1975: Status]_Emergent |
| 14 | VD-Aortic Valve Disease Primary Etiology [1646: VDAoPrimEt]_Degenerative- Calcified |
| 15 | Classification-NYHA [915: ClassNYH]_Class II |
| 16 | RF-Pneumonia [465: Pneumonia]_No |
| 17 | Classification-NYHA [915: ClassNYH]_Not documented |
| 18 | Cardiogenic Shock [930: CarShock]_Yes - At the time of the procedure |
| 19 | RF-Tobacco Use [400: TobaccoUse]_Former smoker |
| 20 | Prev CAB [670: PrCAB] |

**Supplement 7.**
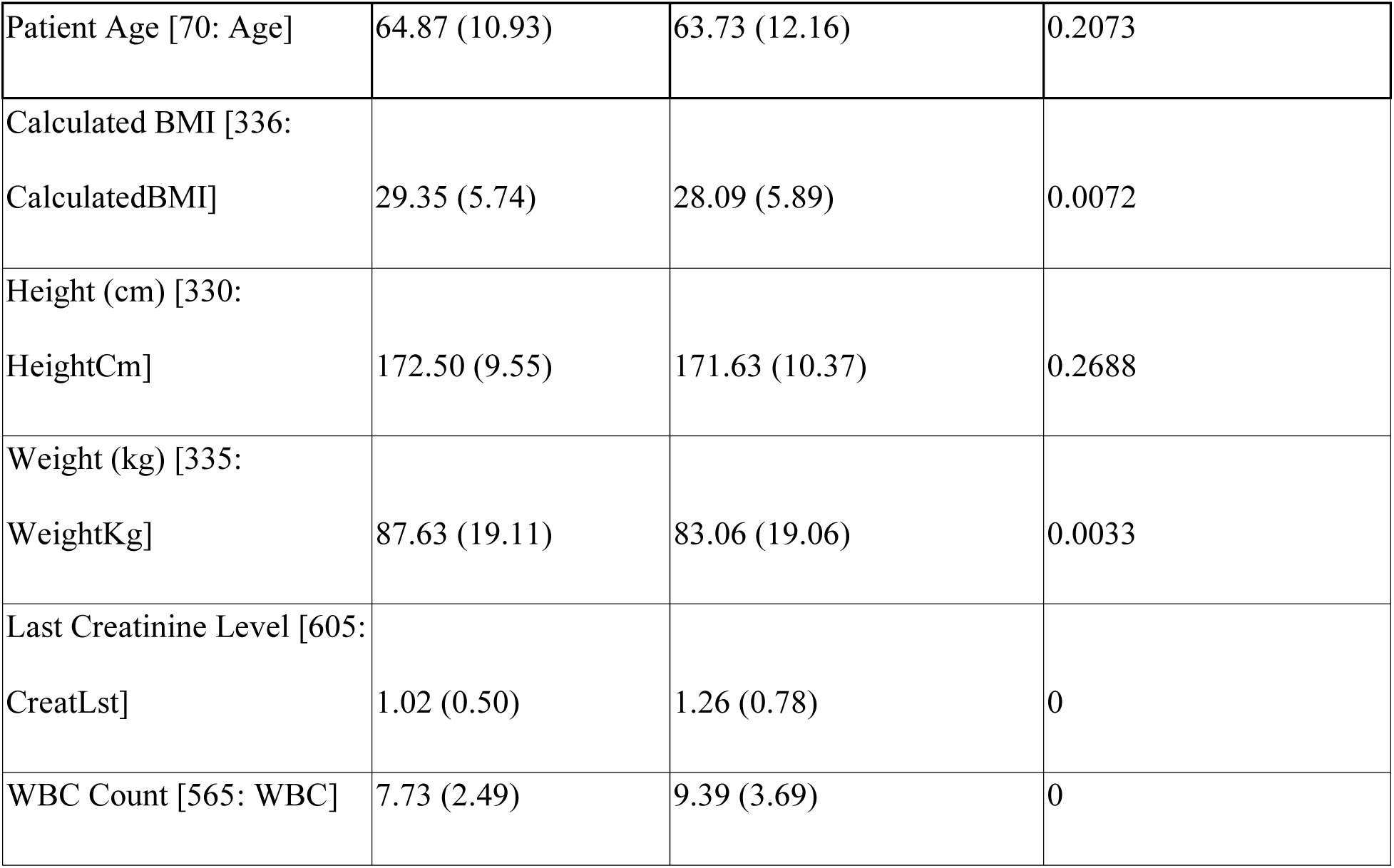
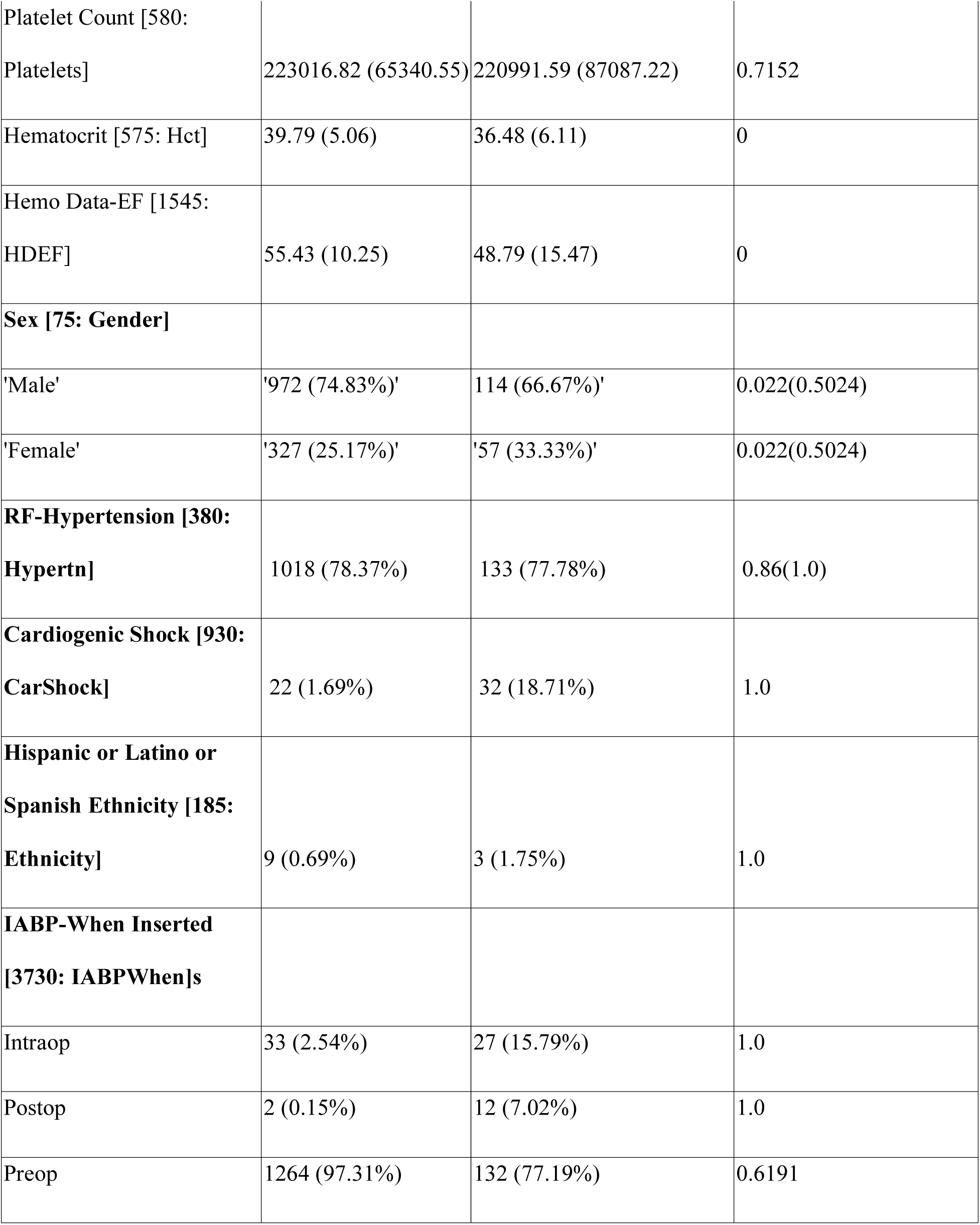
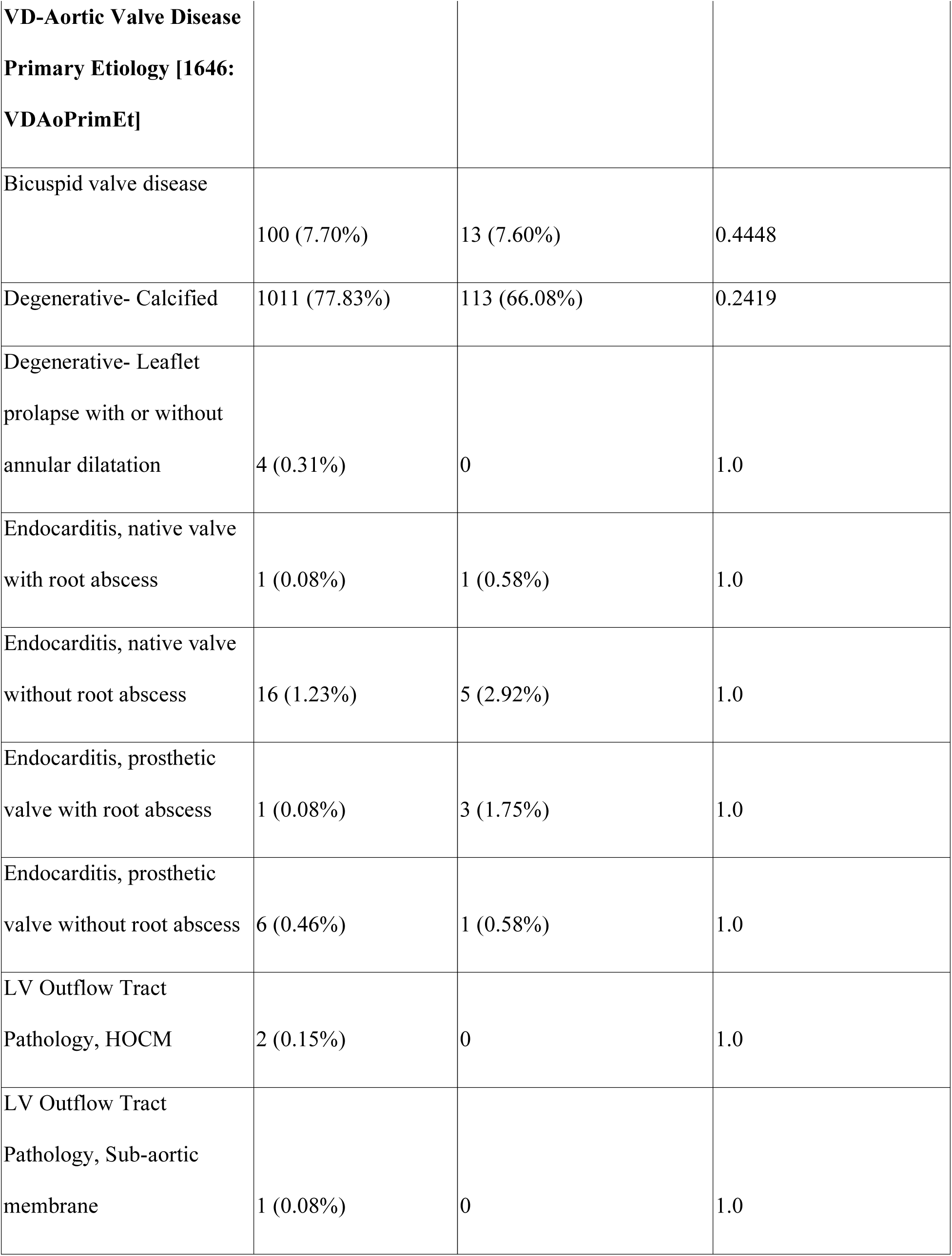
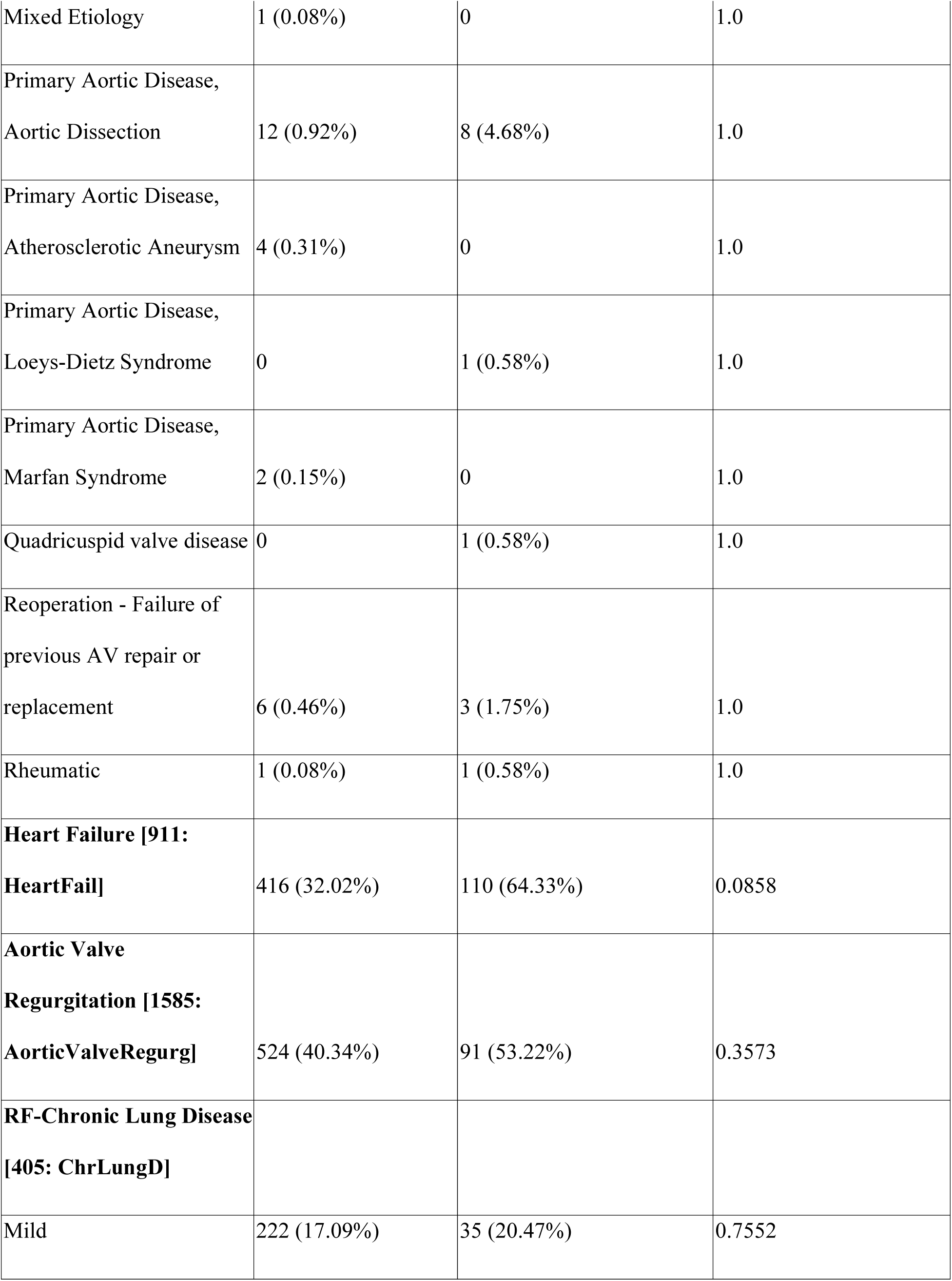
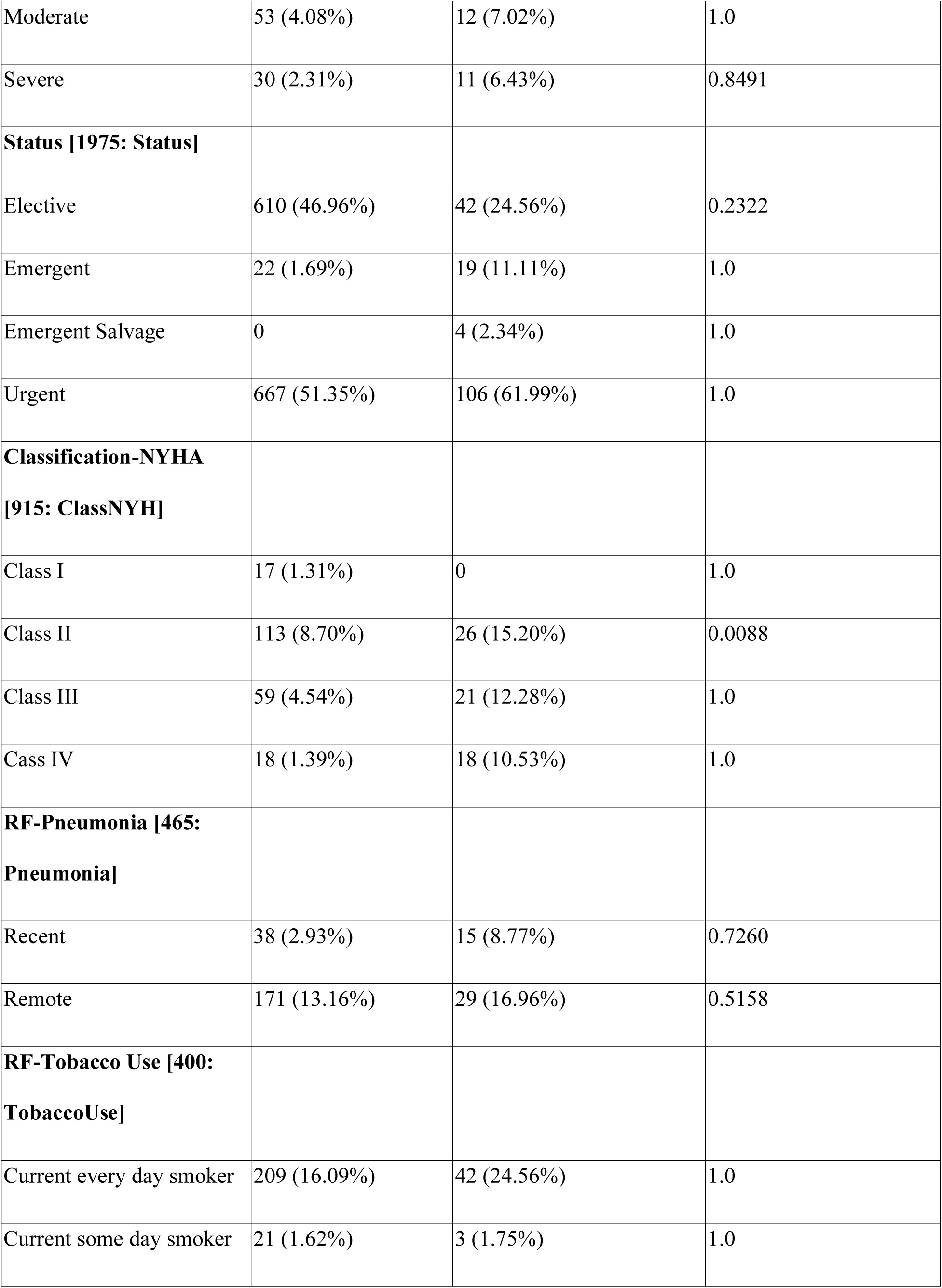
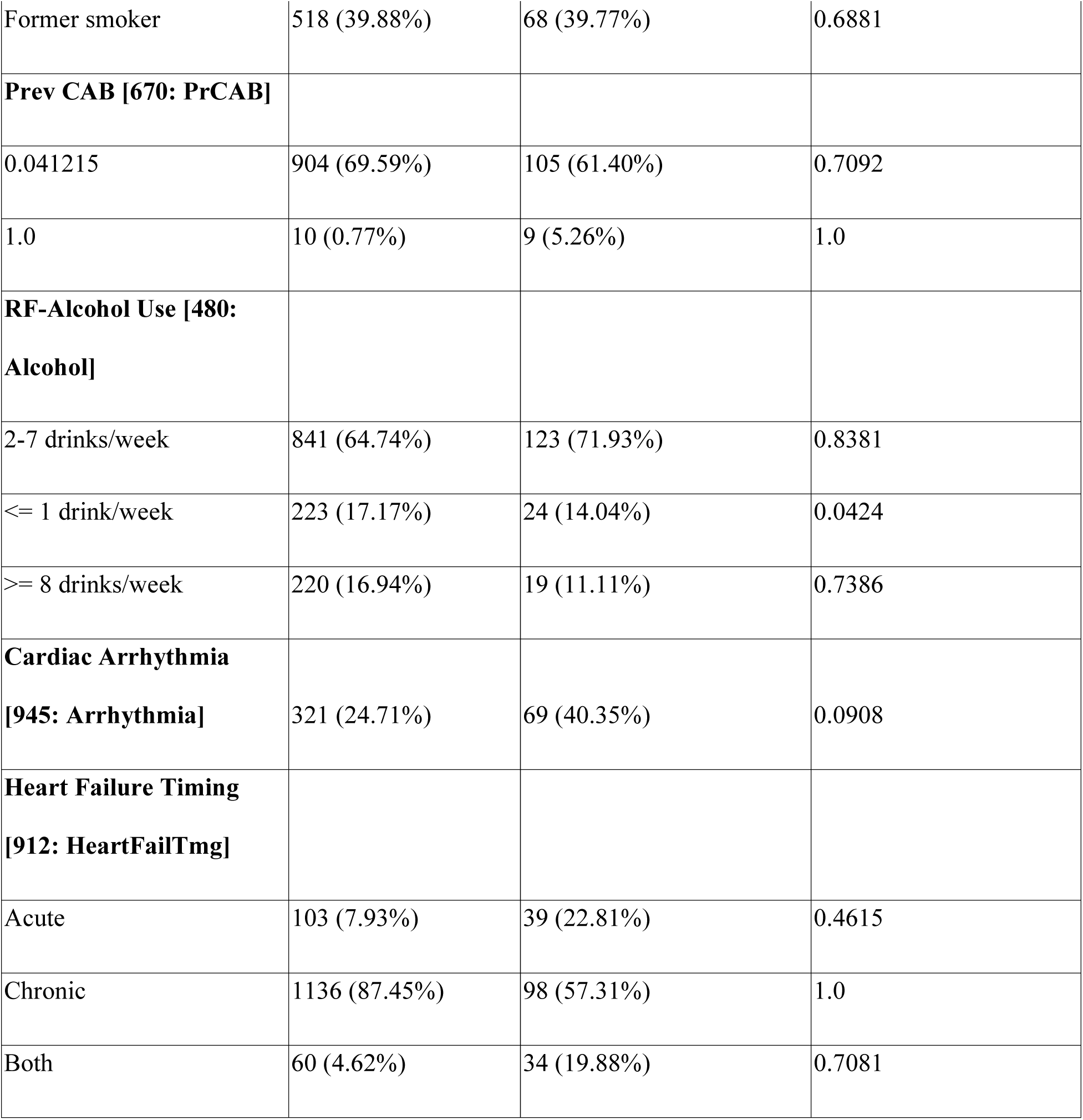
Additional static variables with scores.

**Supplement 8.**
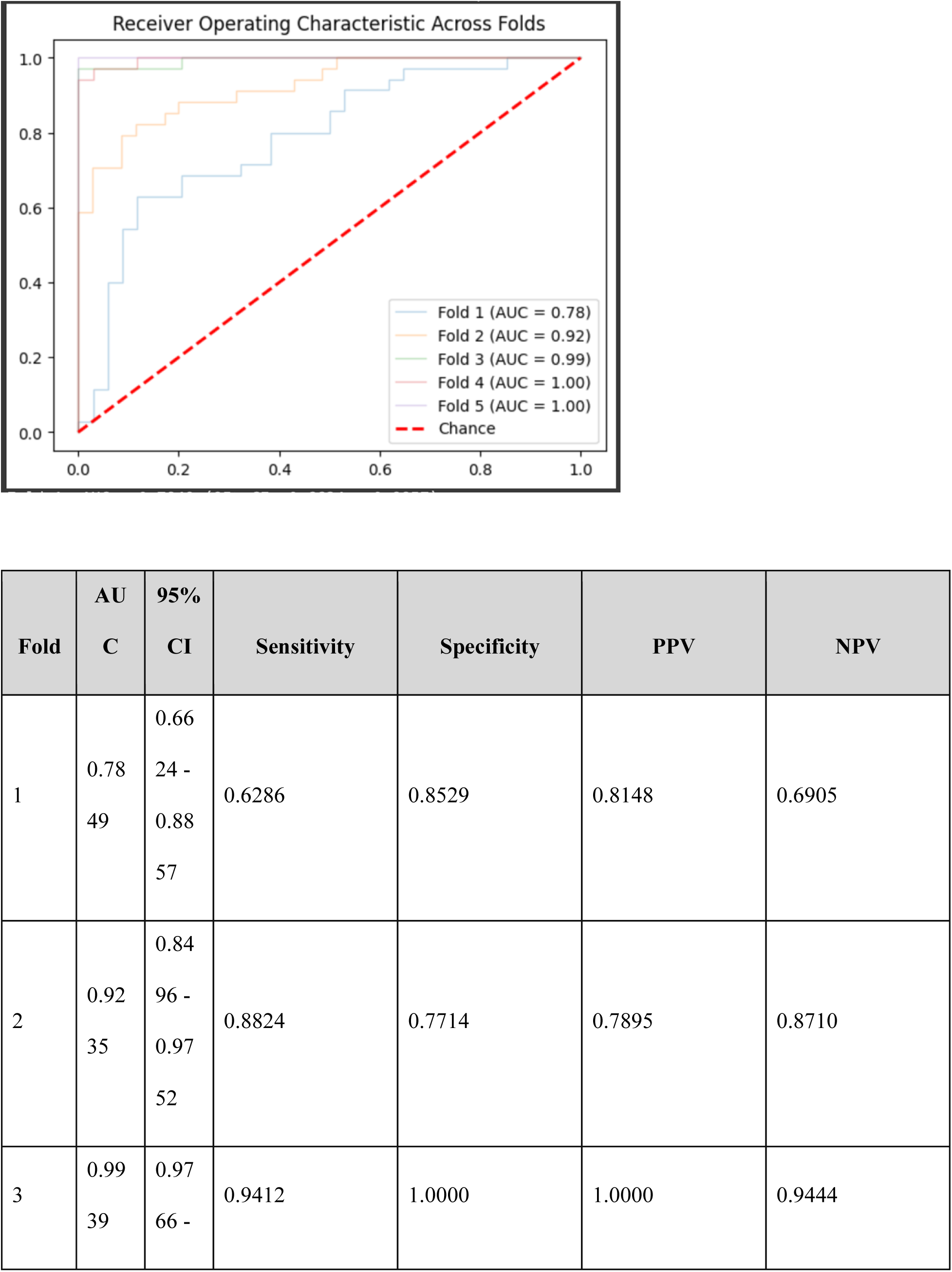
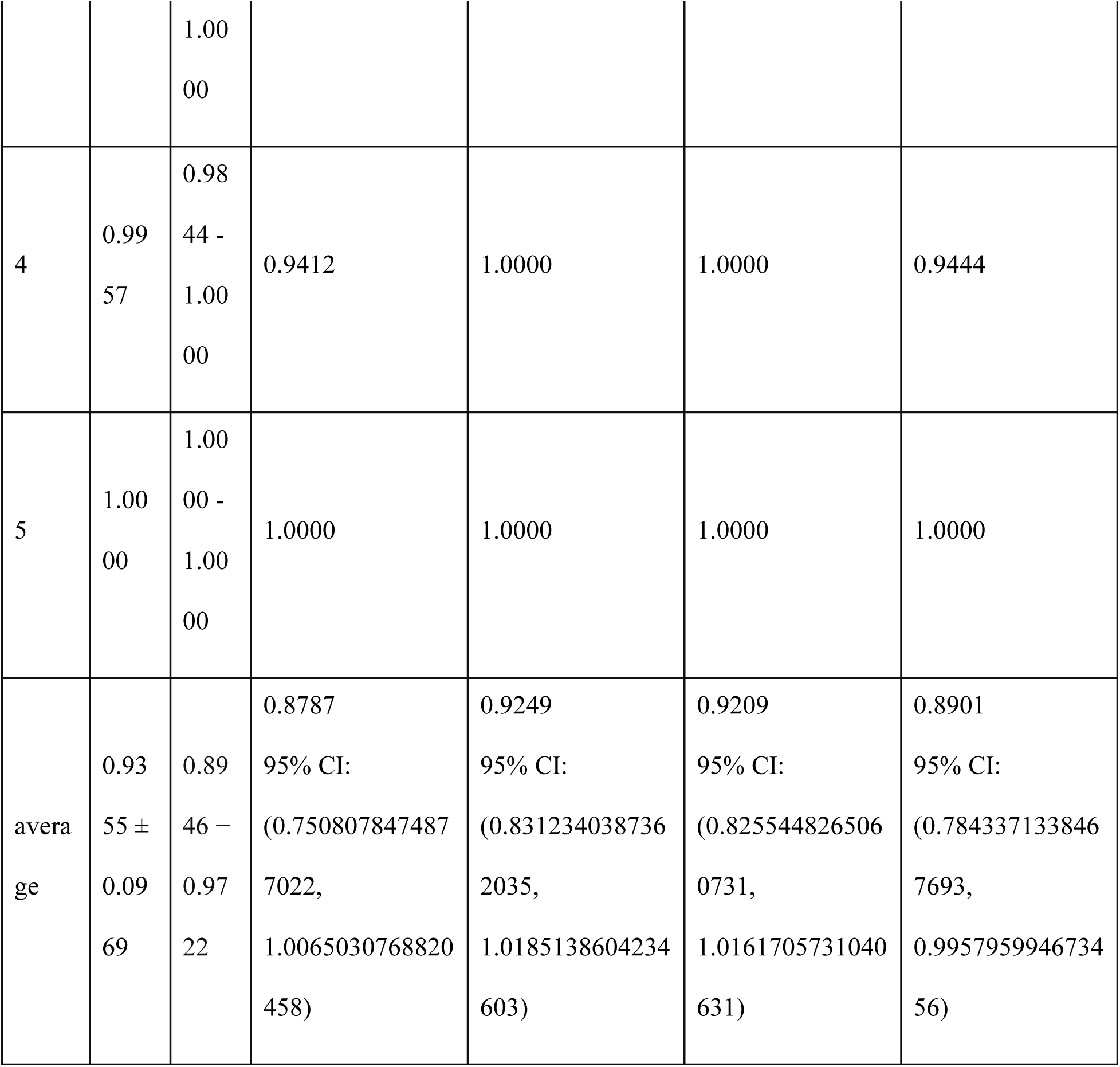
Ablation study results. Results after removing column “ABP Dia”:

Results after removing column “ABP Mean”:

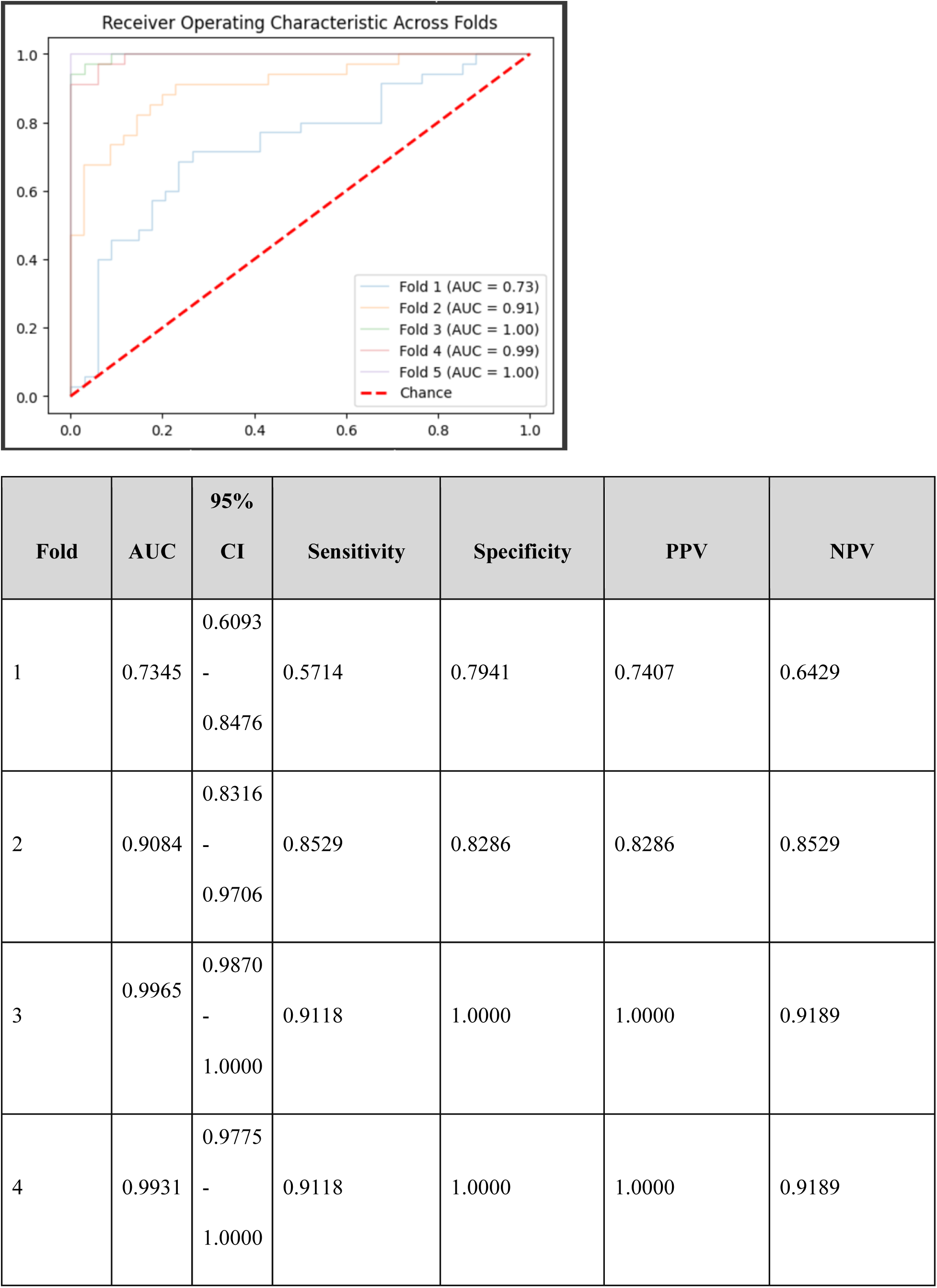
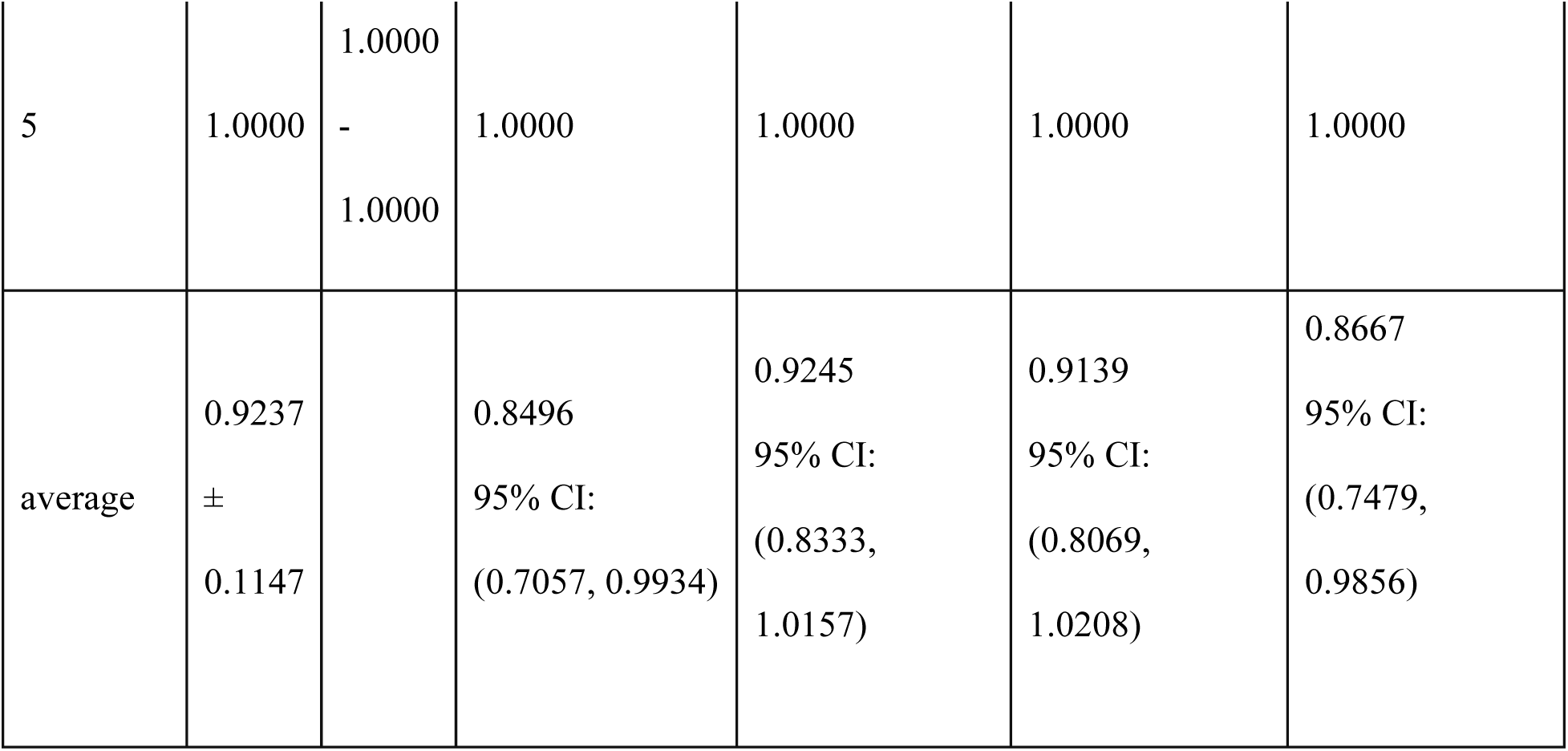

Results after removing column “ABP Sys”:

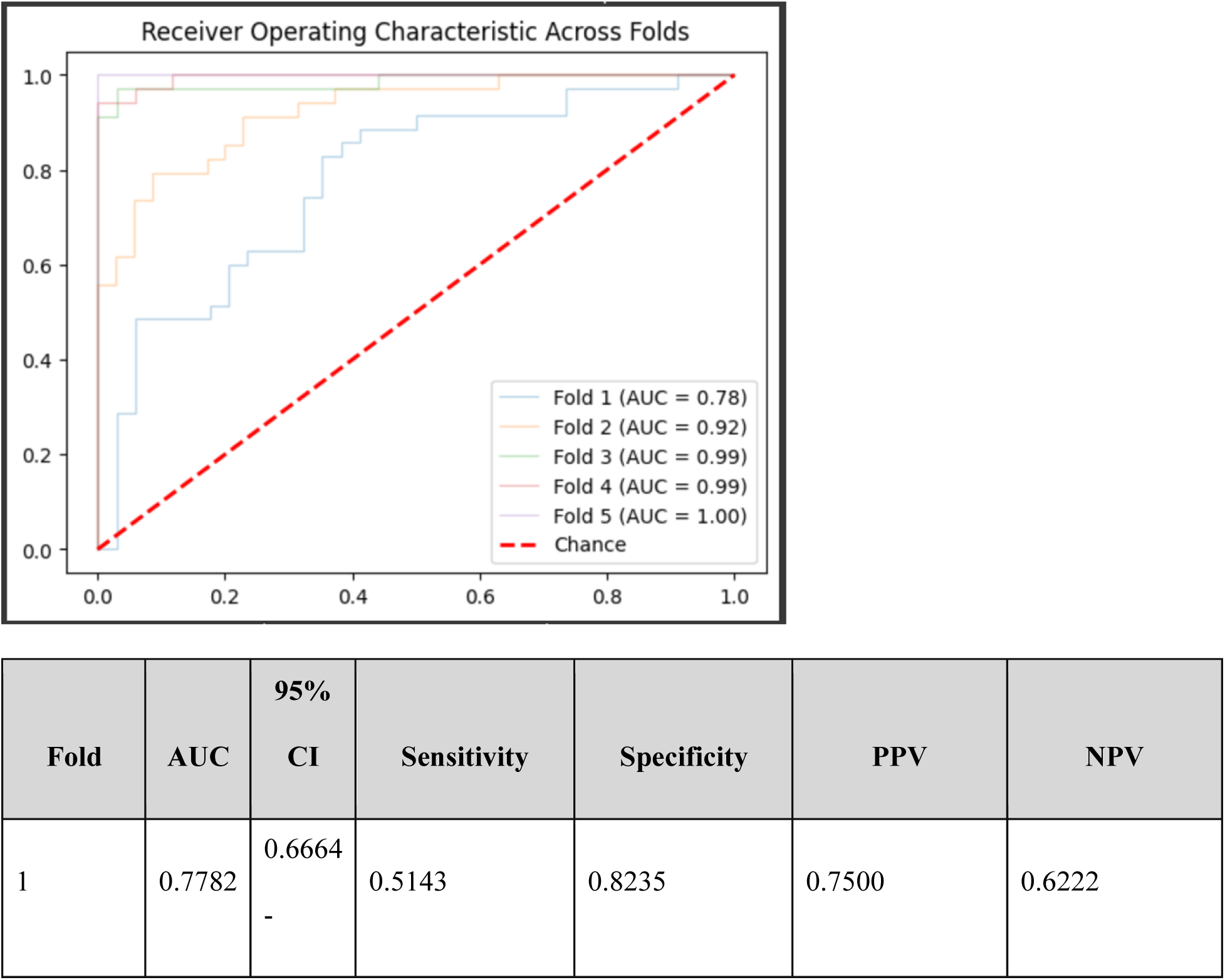
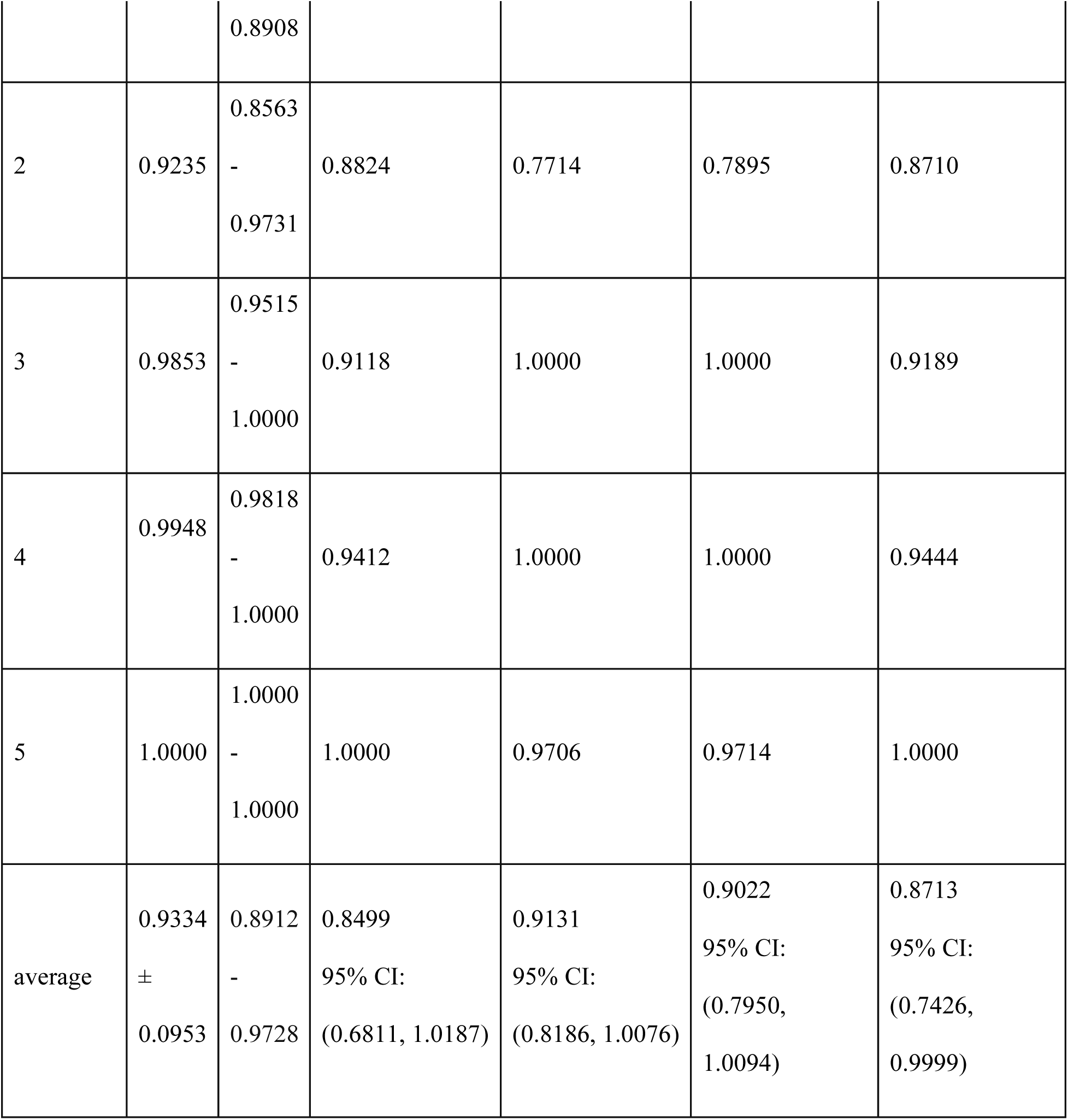

Results after removing column “AO Dia”:

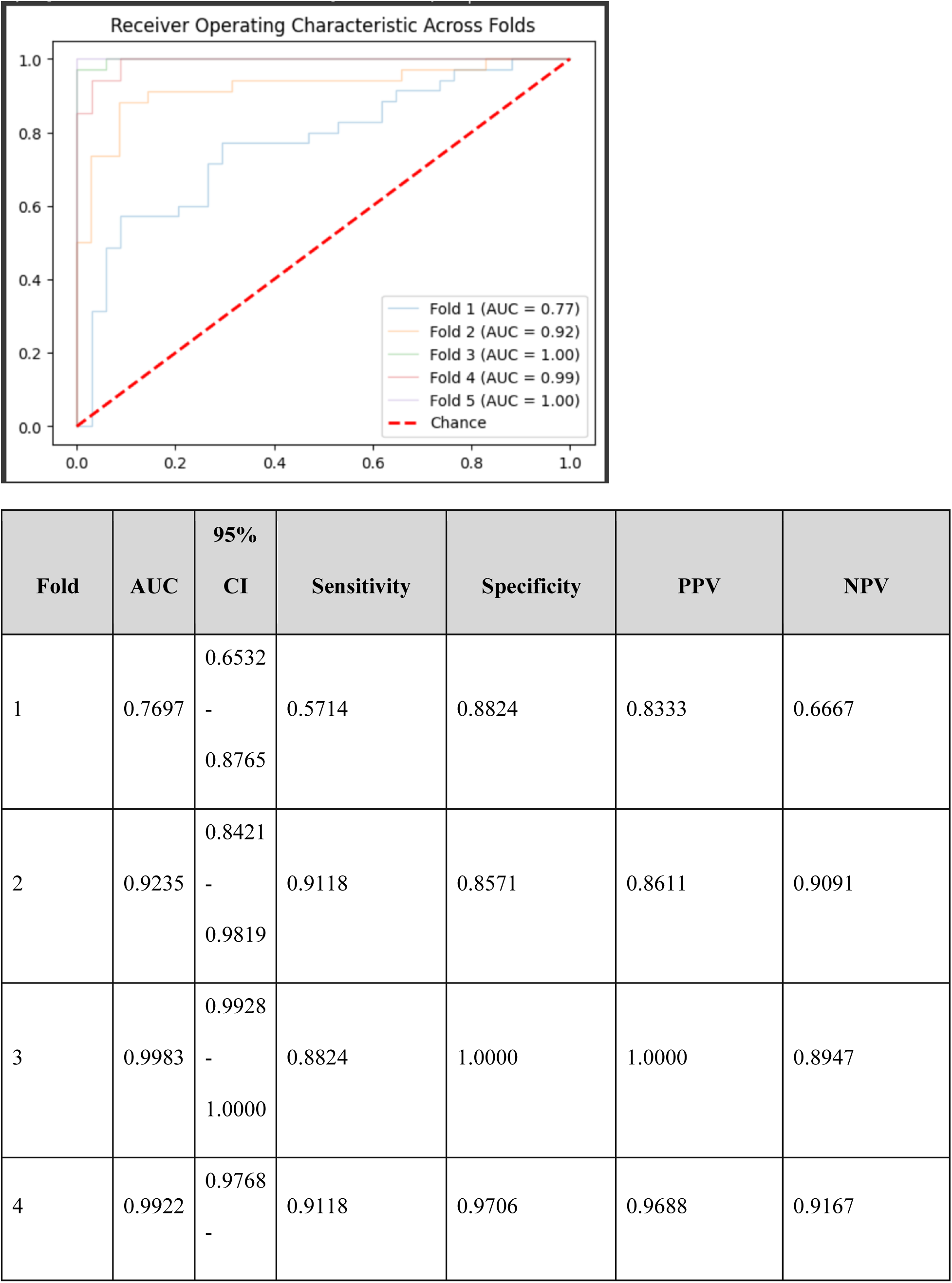
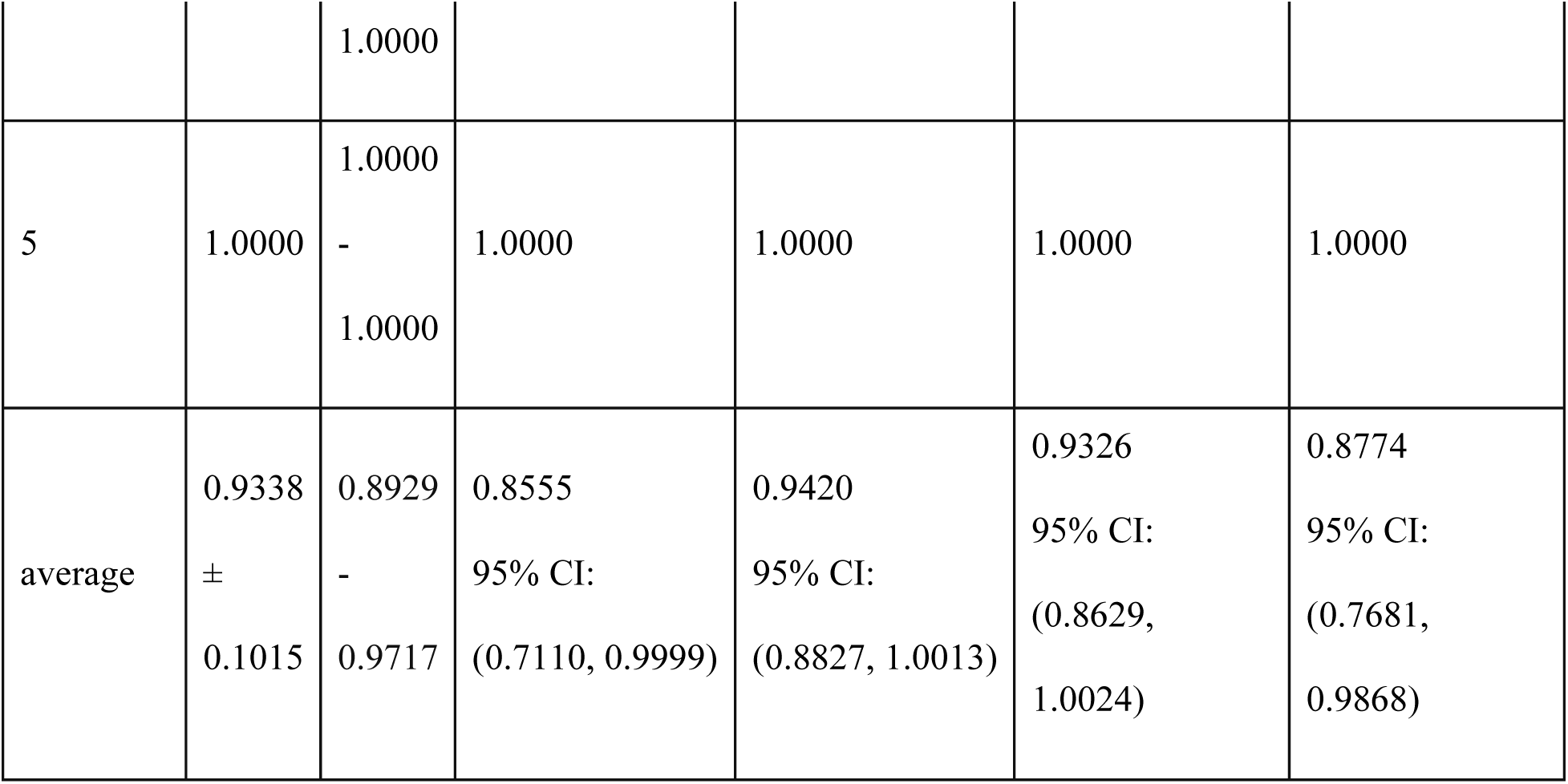

Results after removing column “AO Mean”:

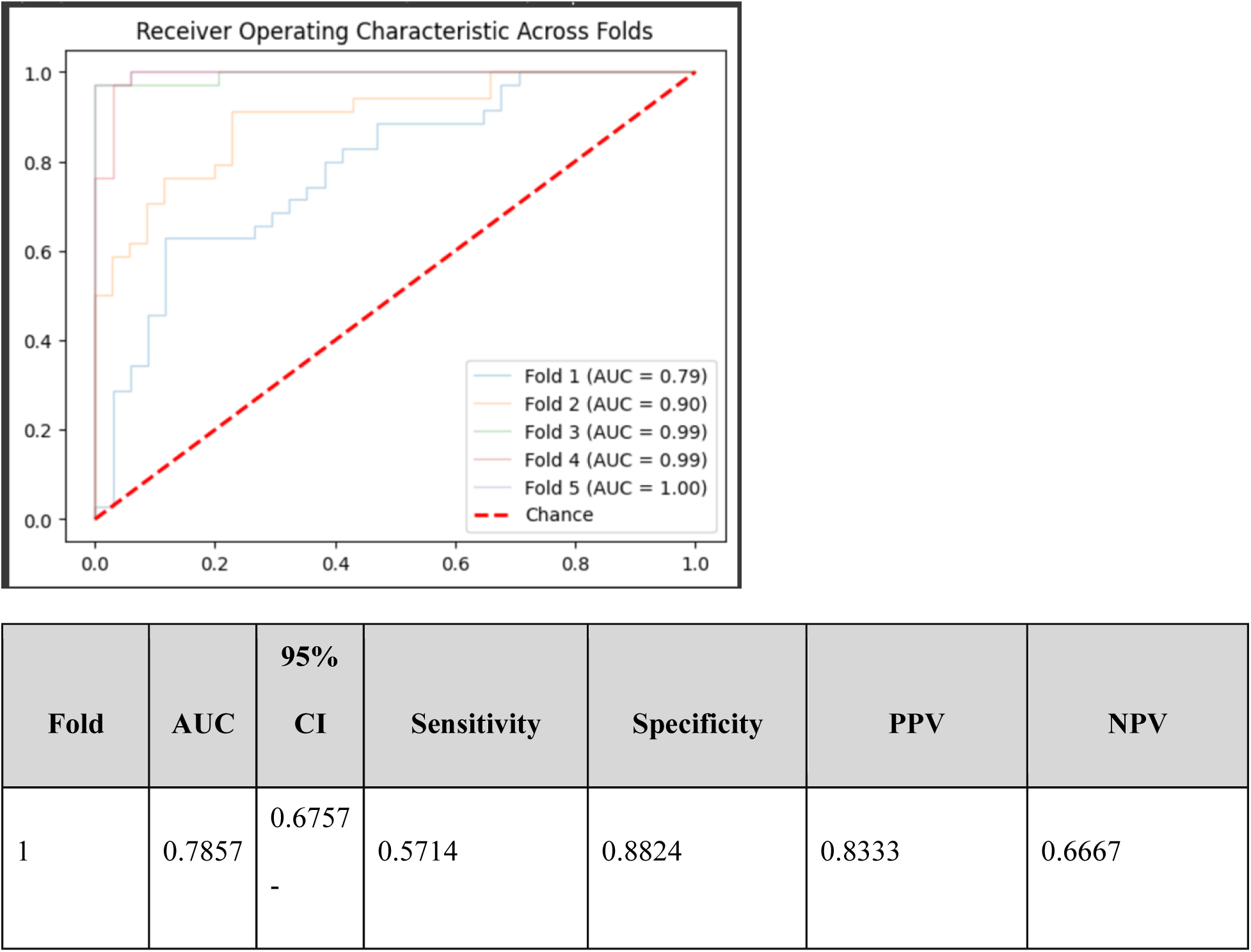
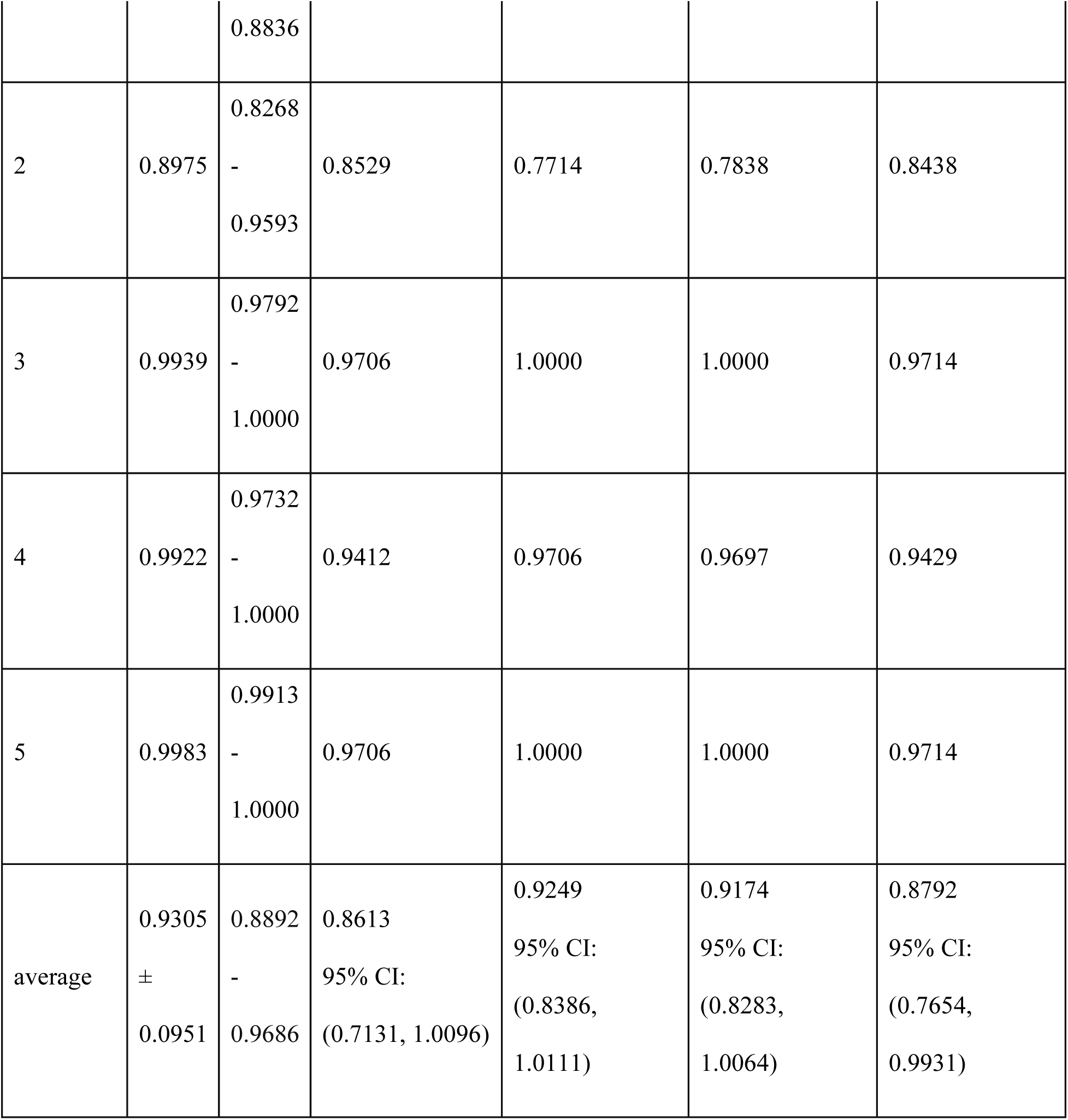

Results after removing column “AO Sys”:

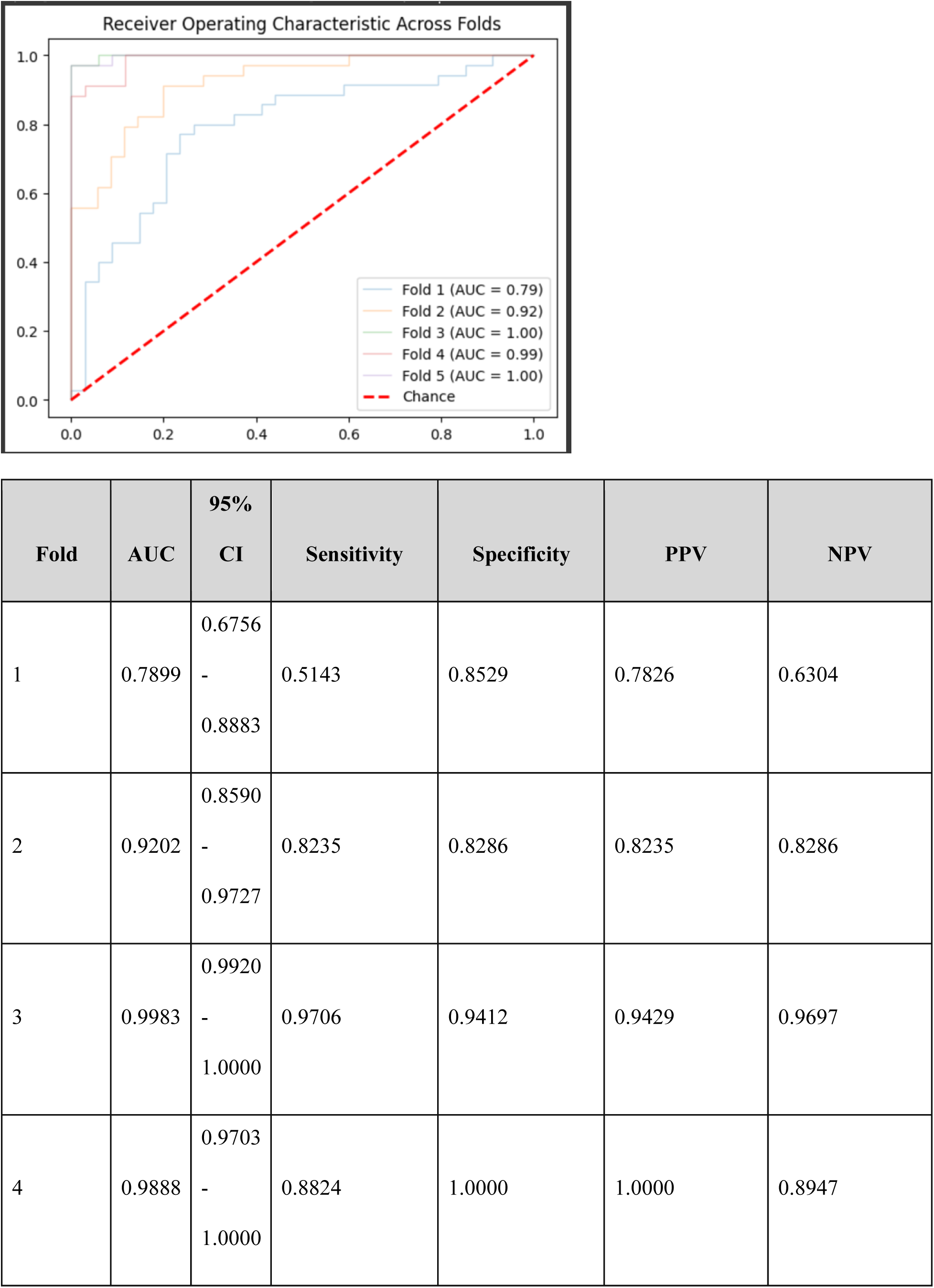
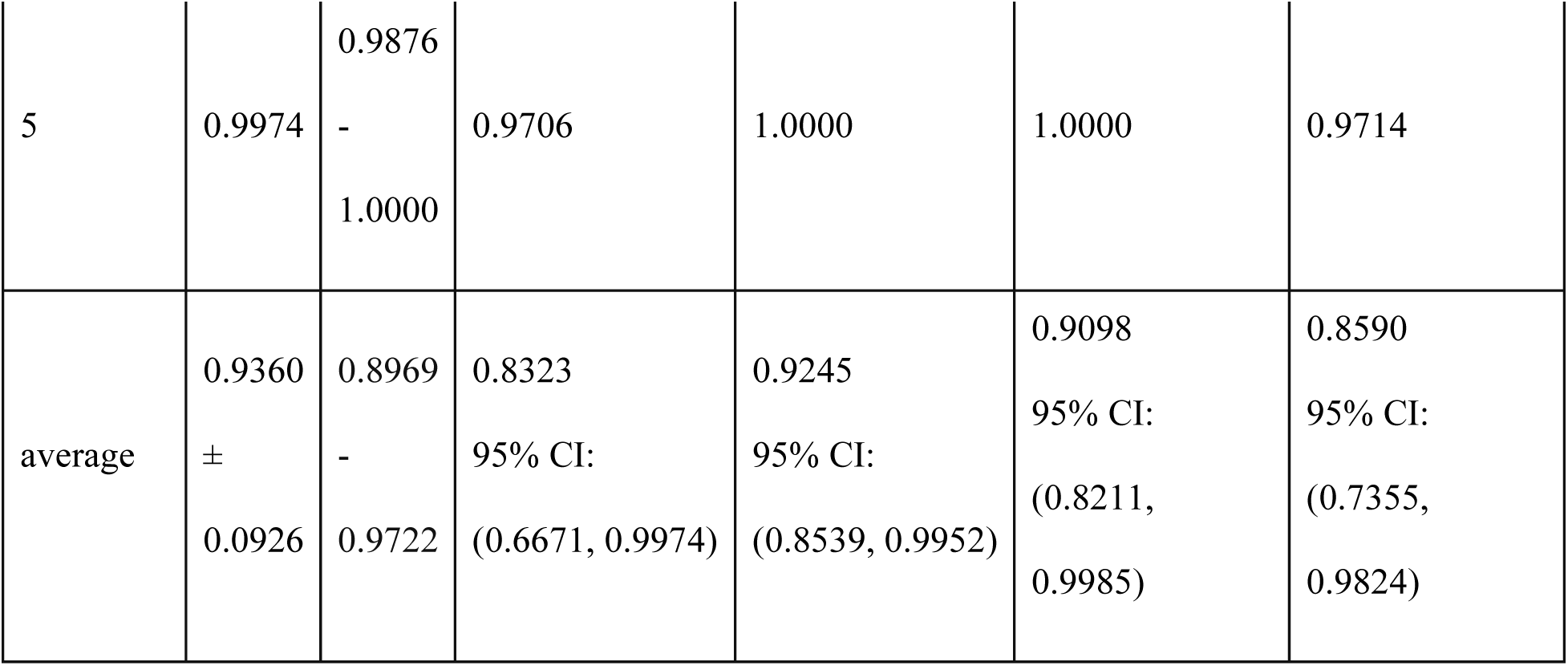

Results after removing column “ART Dia”:

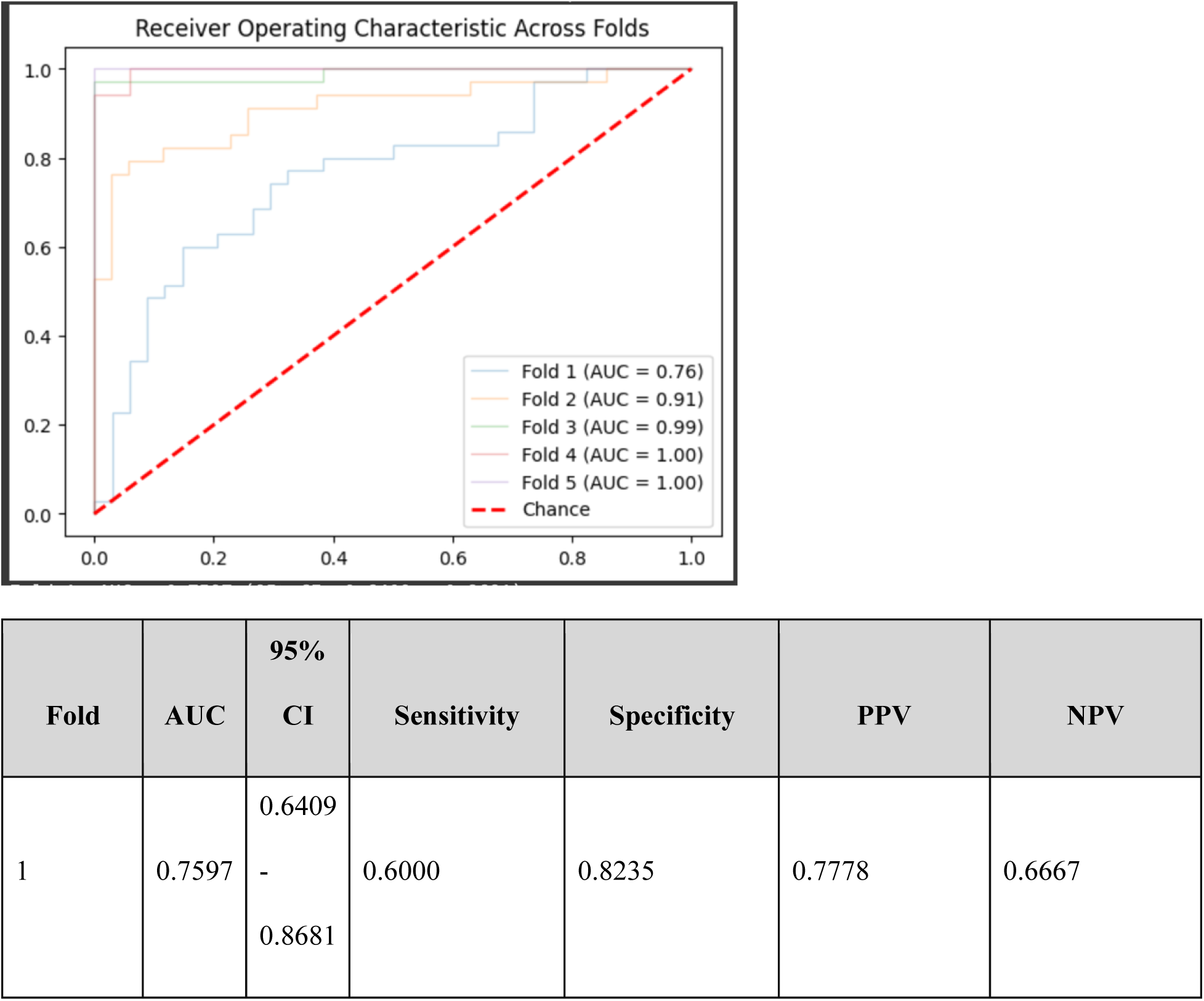
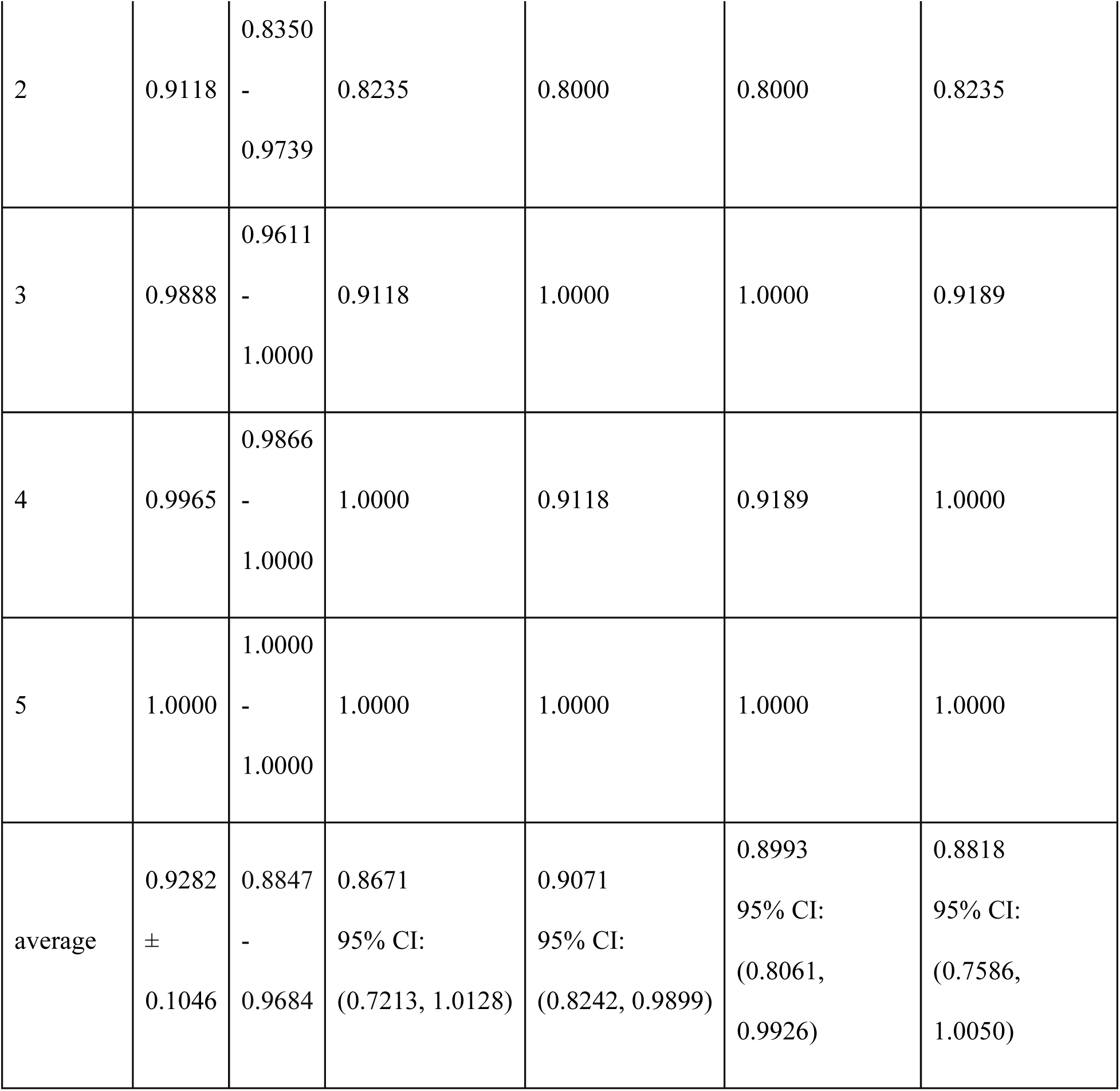

Results after removing column “ART Mean”:

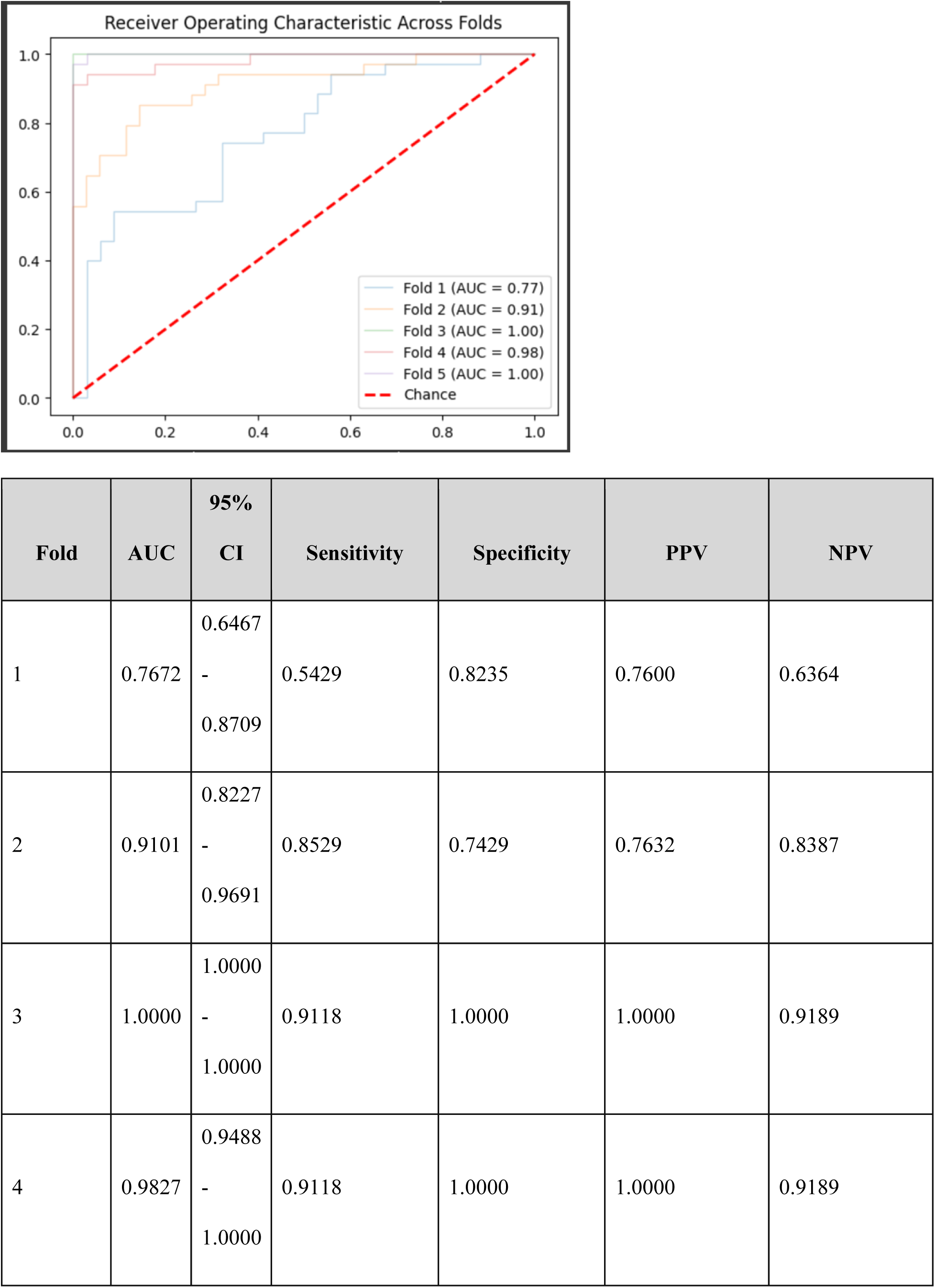
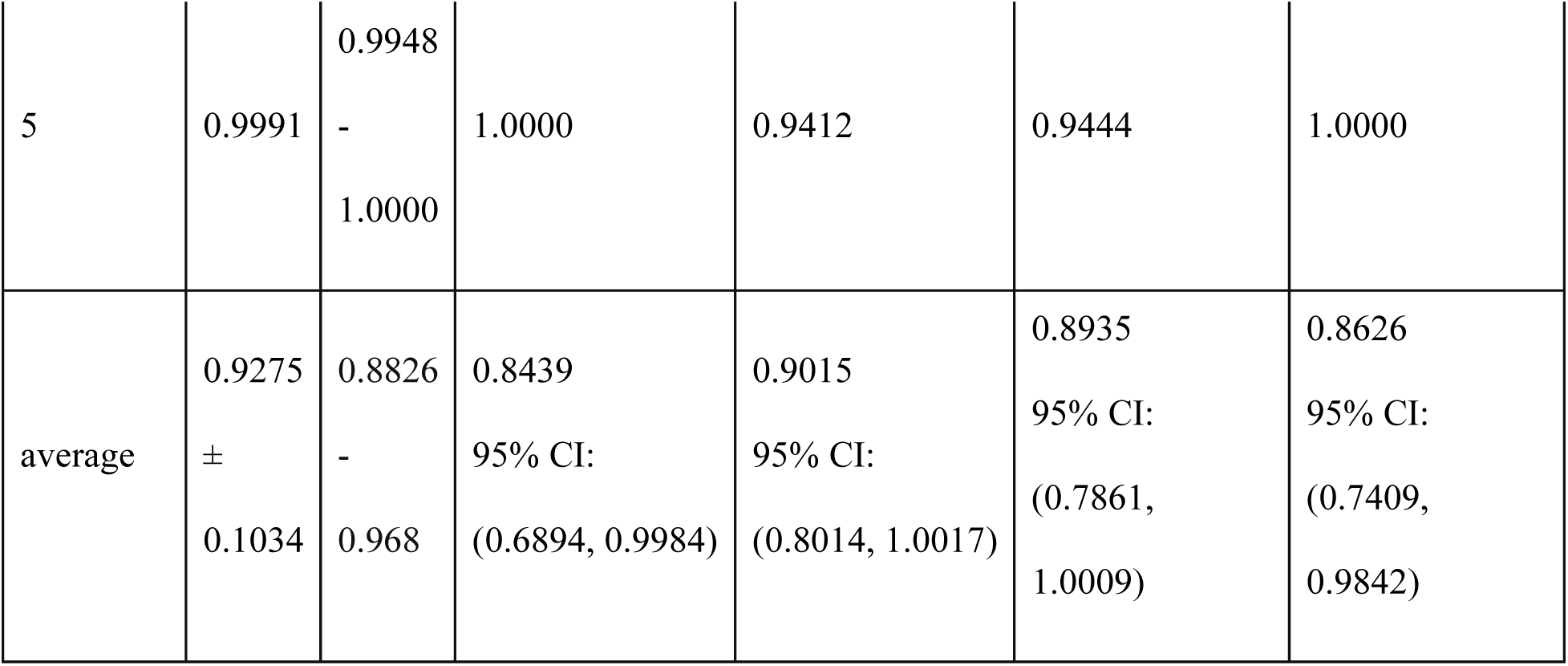

Results after removing column “ART Sys”:

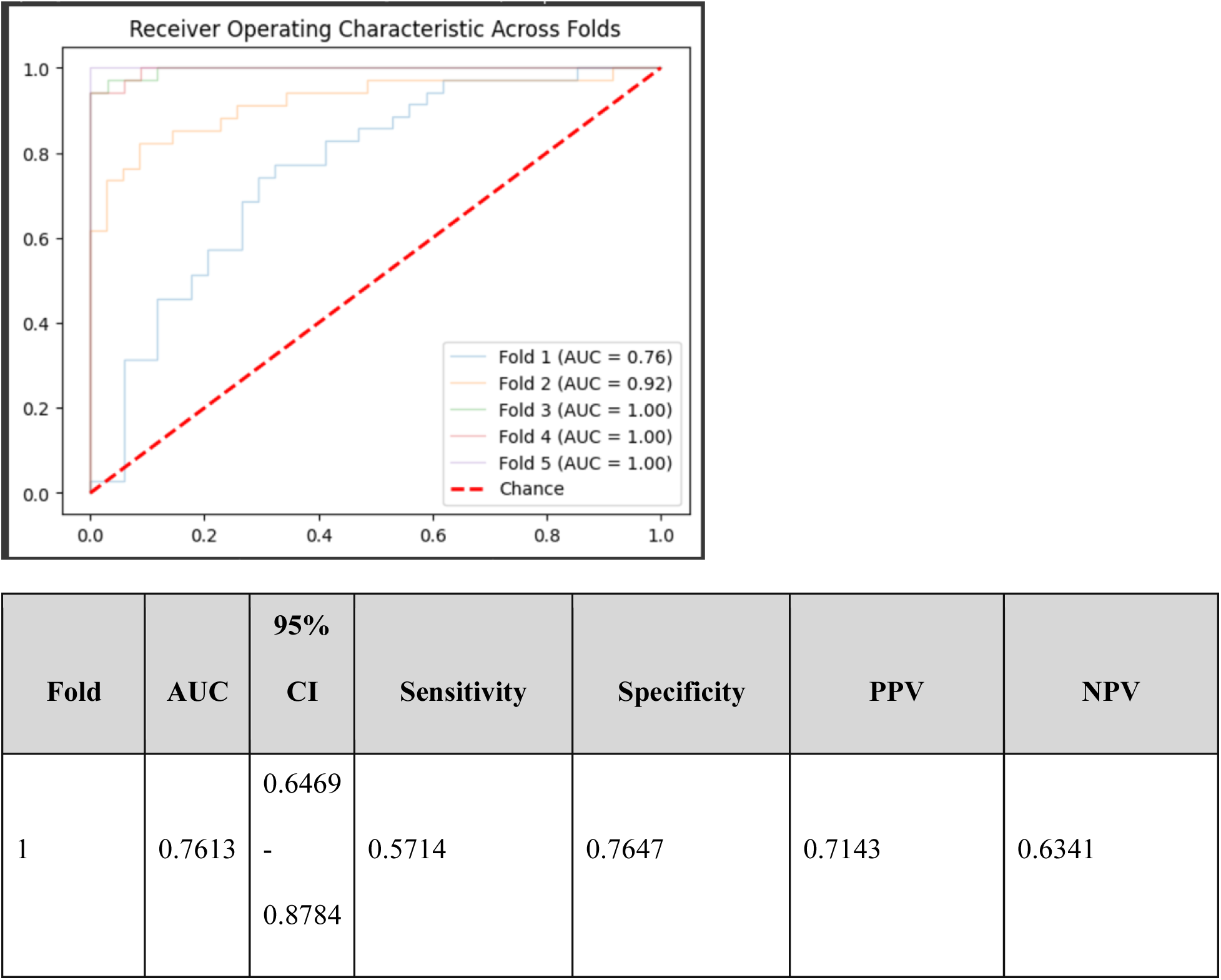
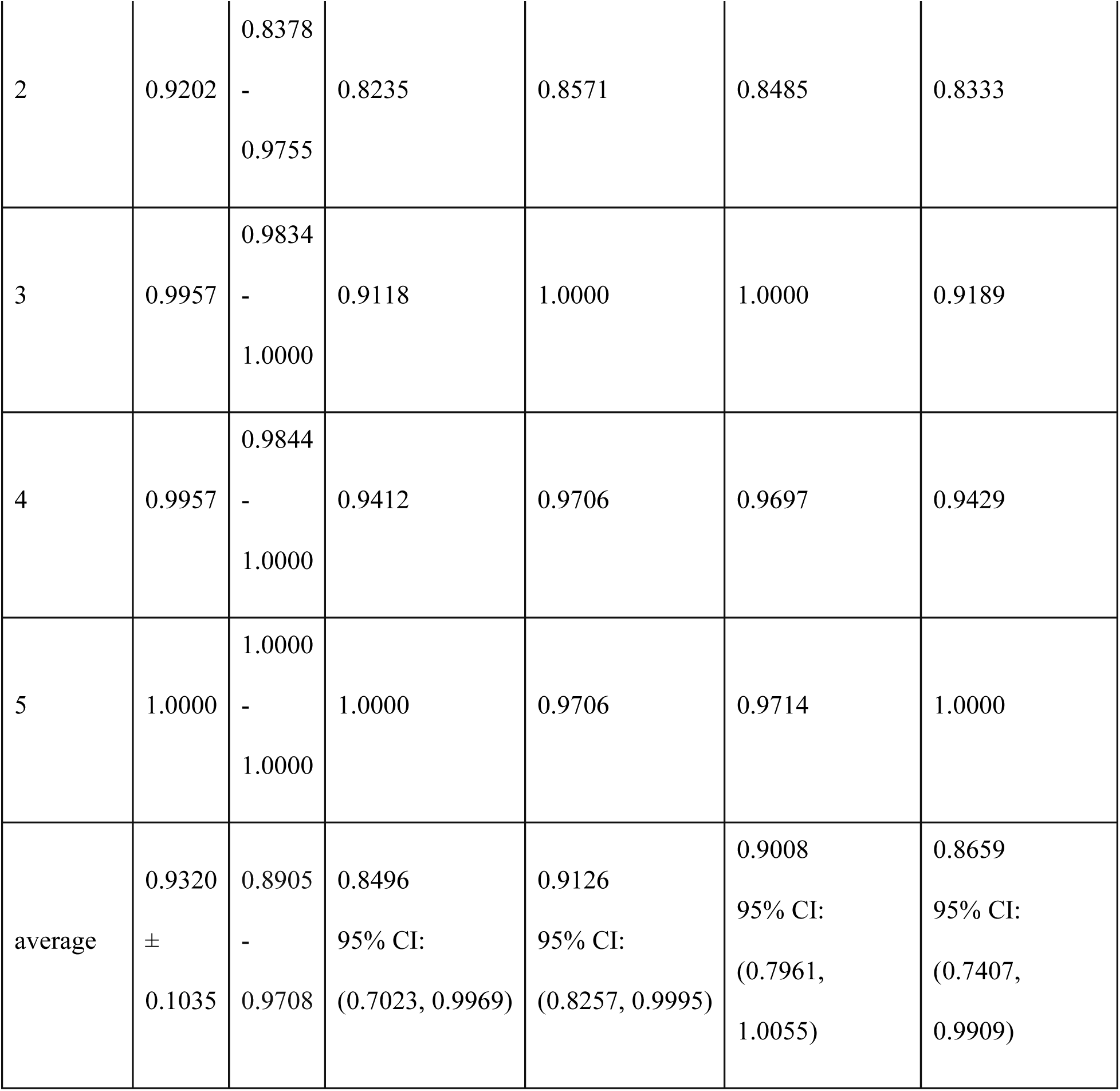

Results after removing column “CVP”:

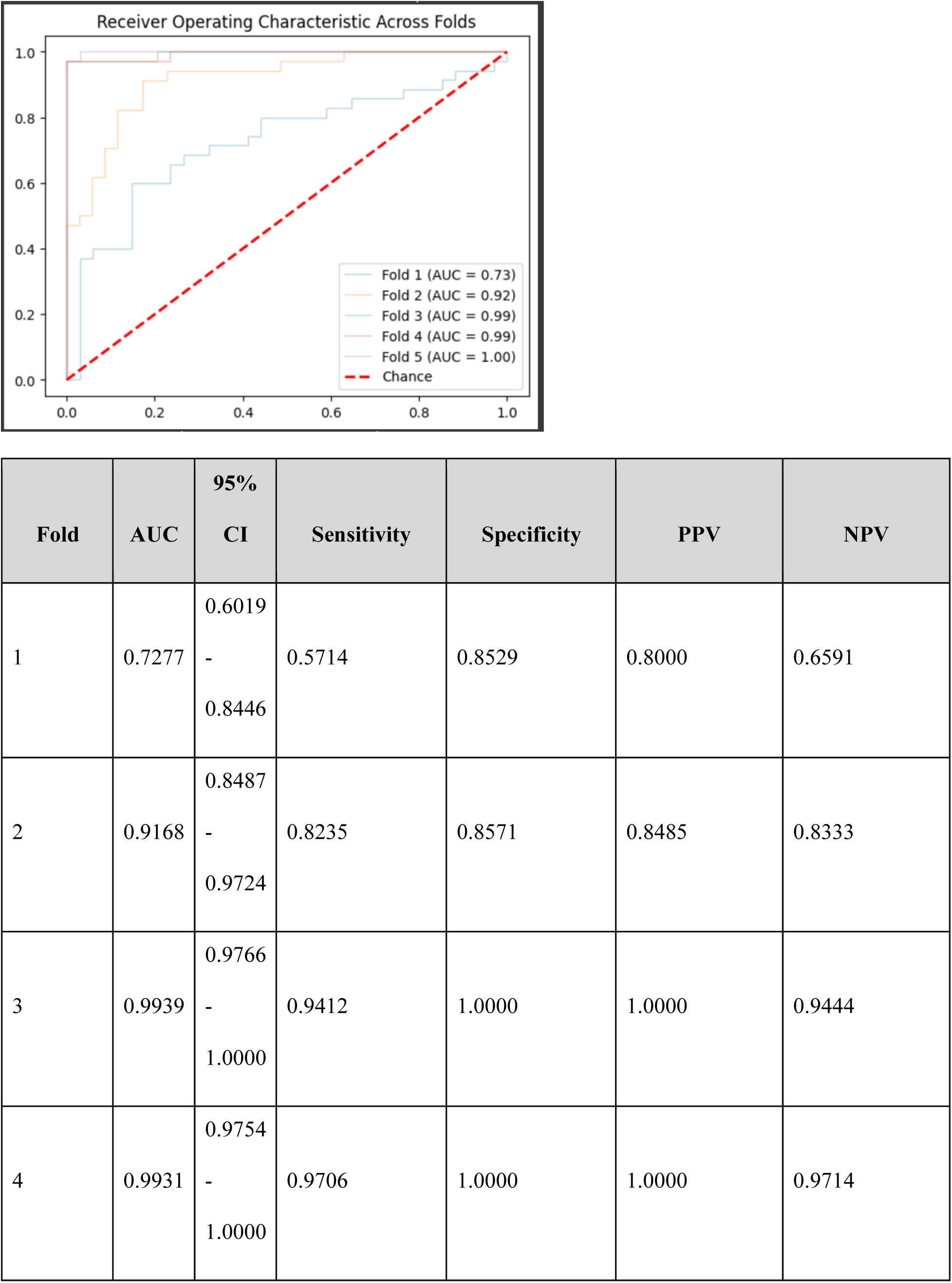
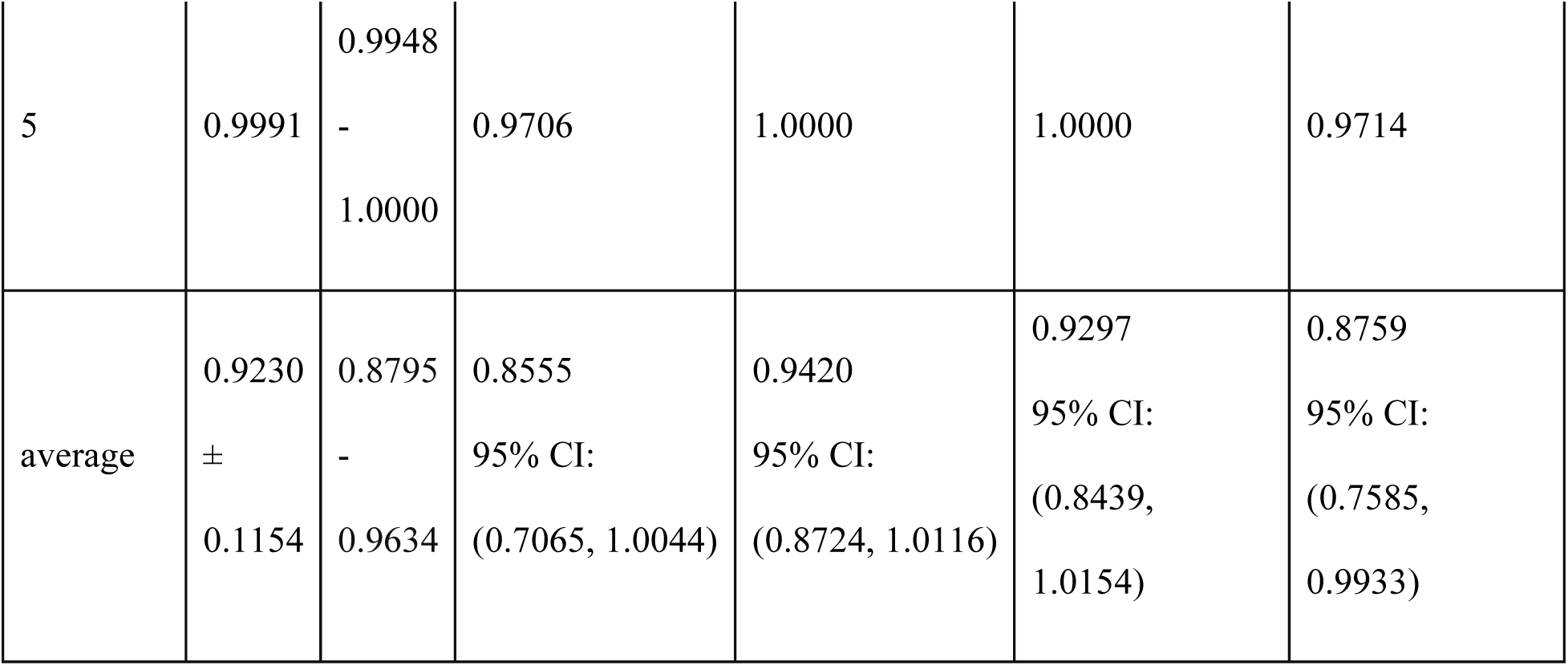

Results after removing column “ETC02”:

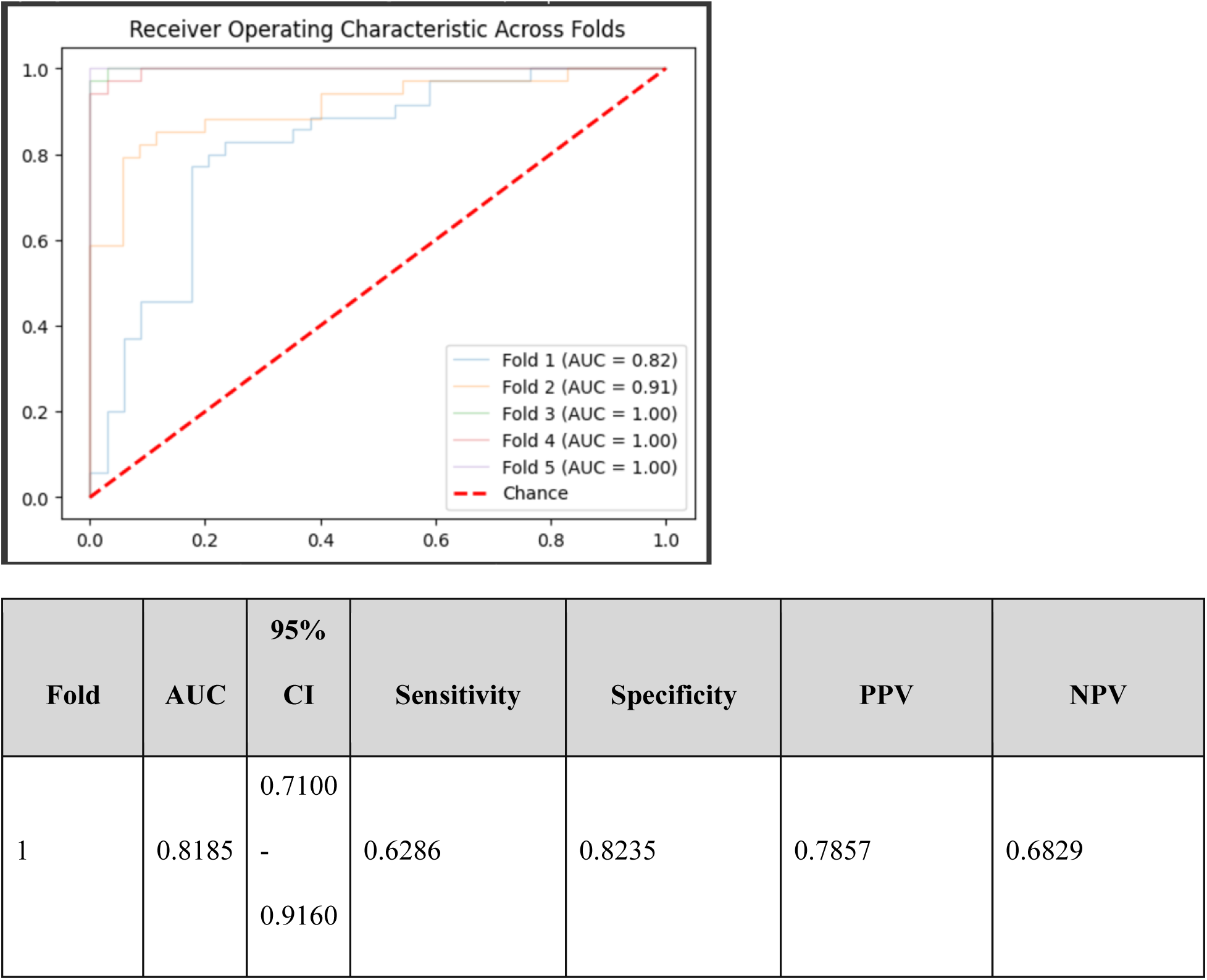
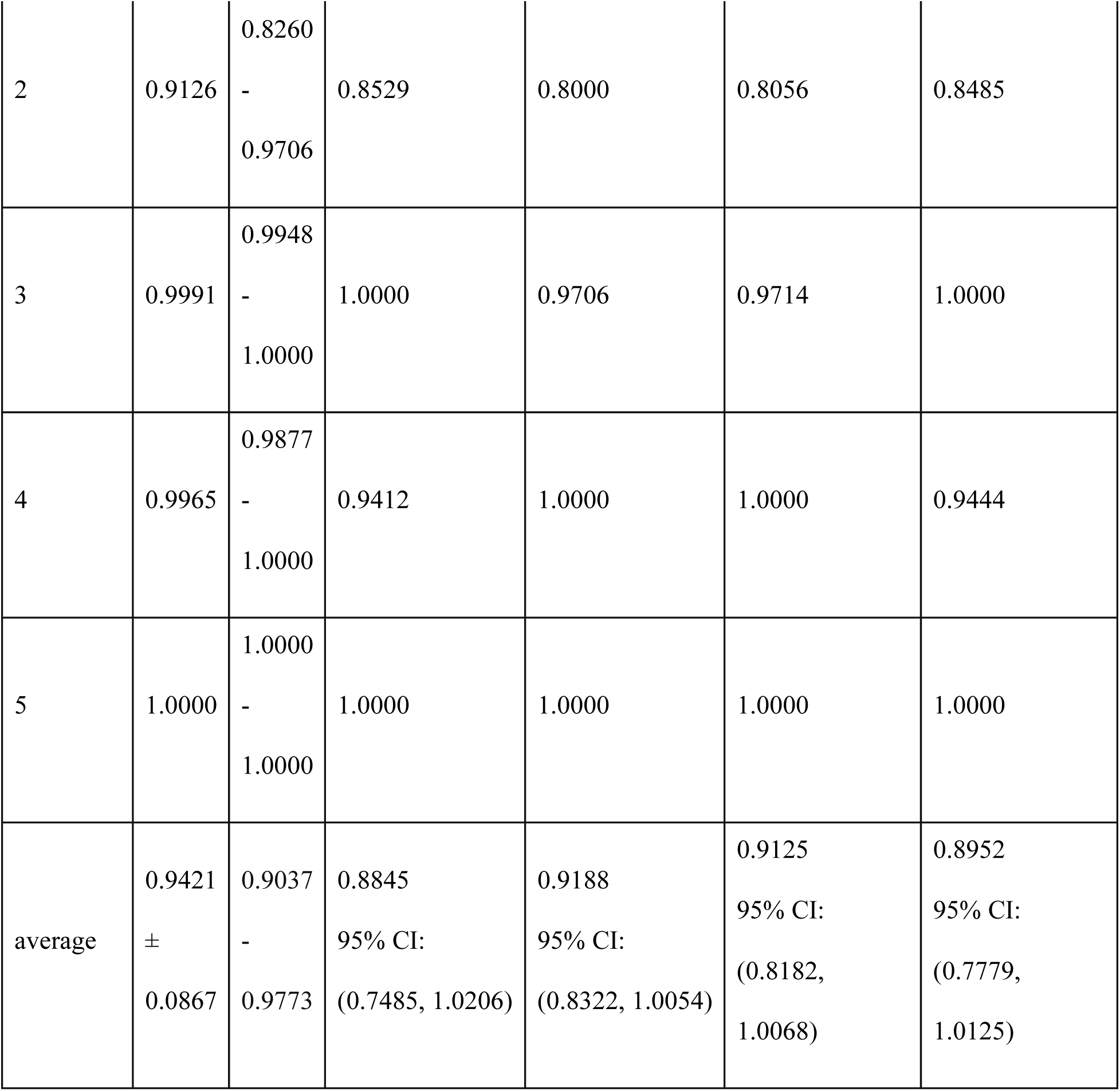

Results after removing column “Core Temp”:

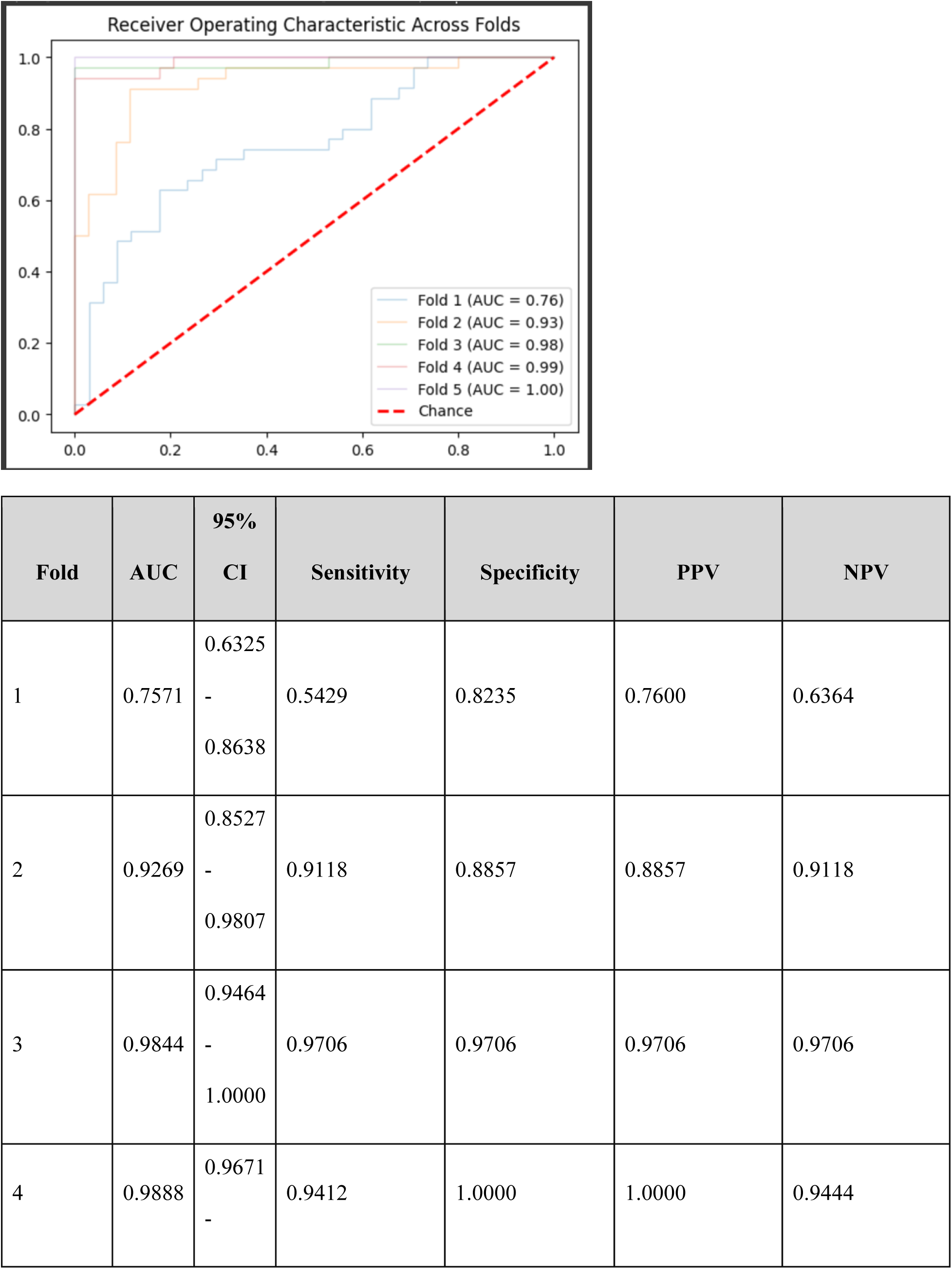
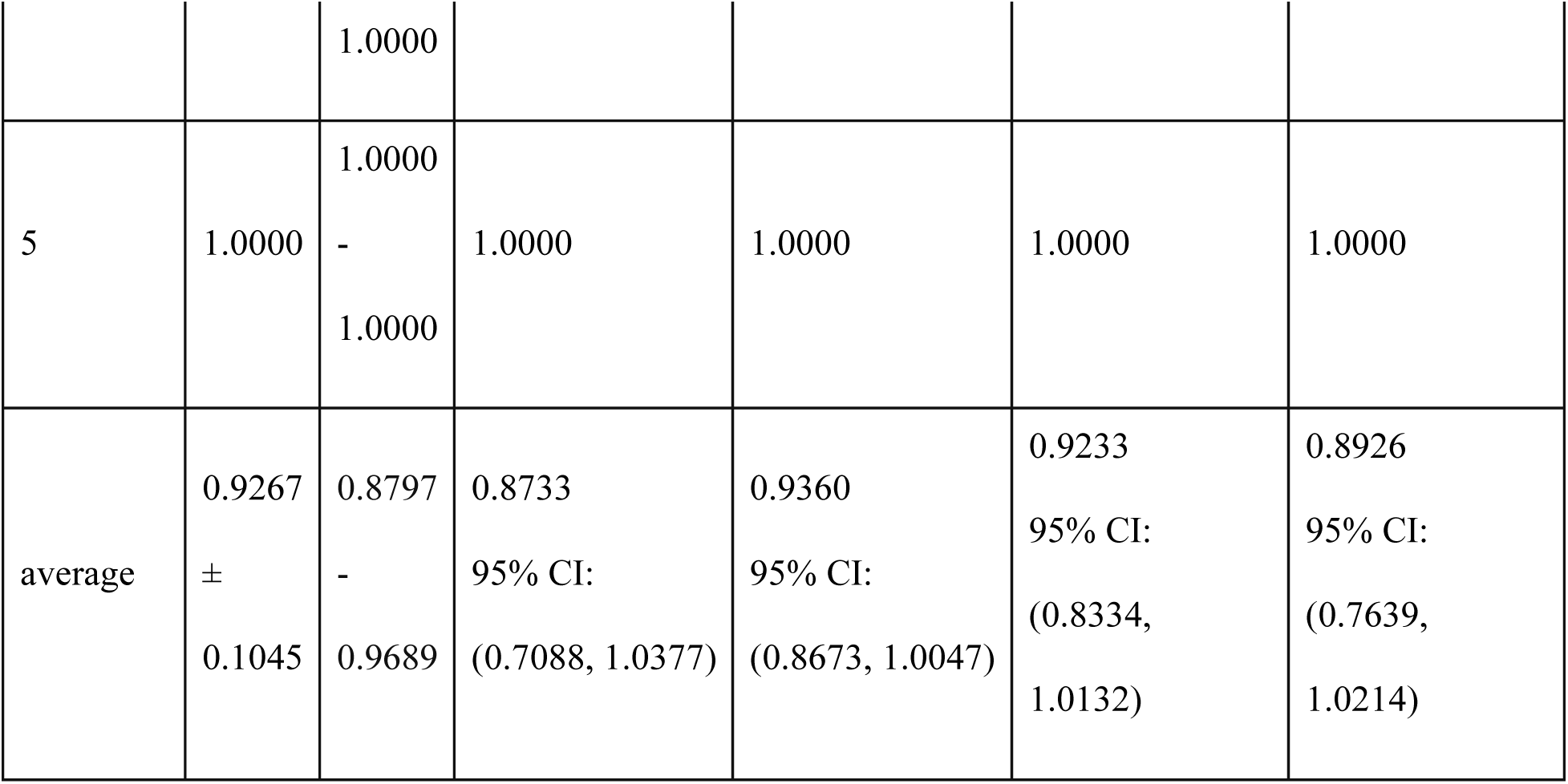

Results after removing column “ECG HR”:

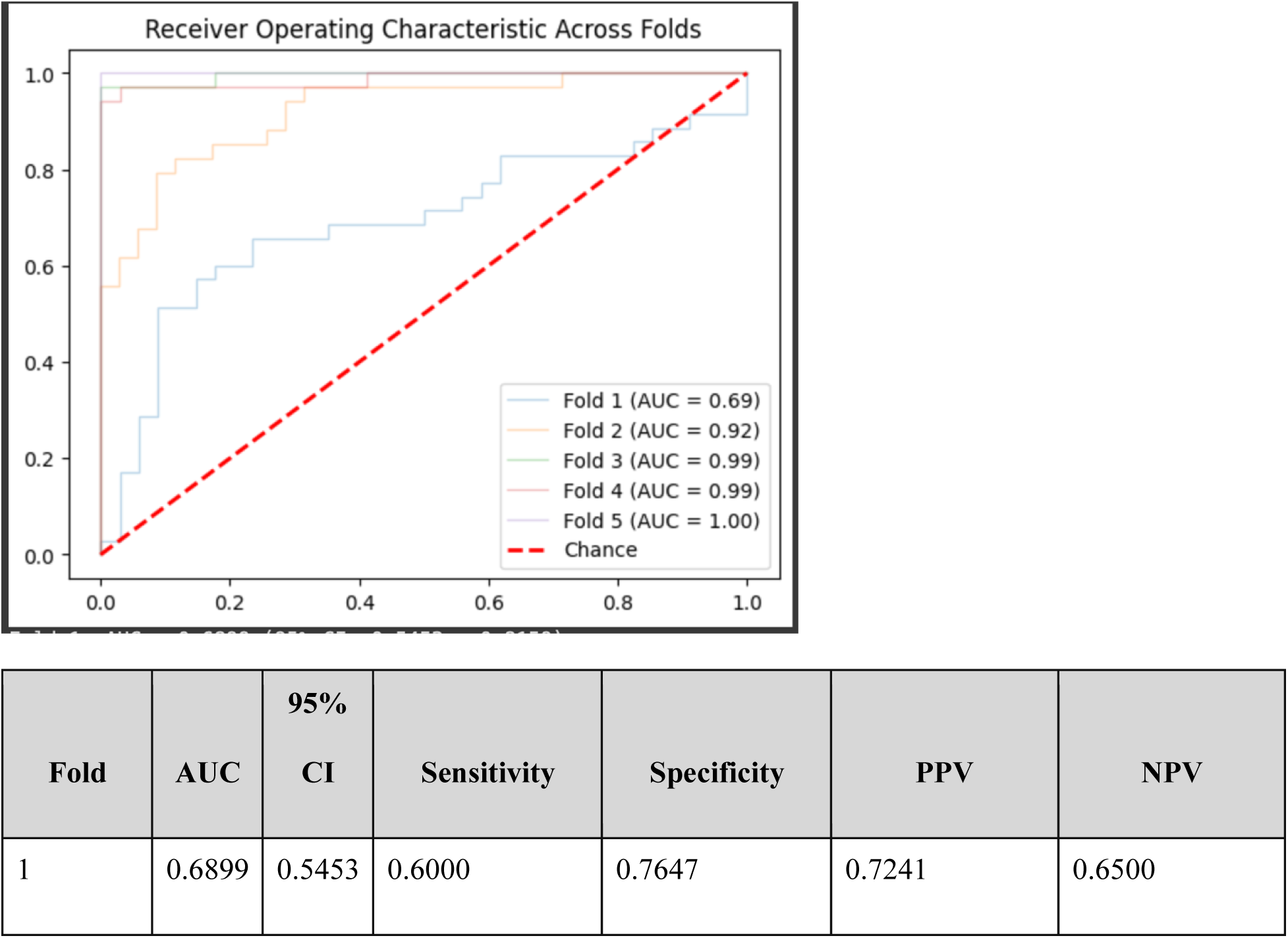
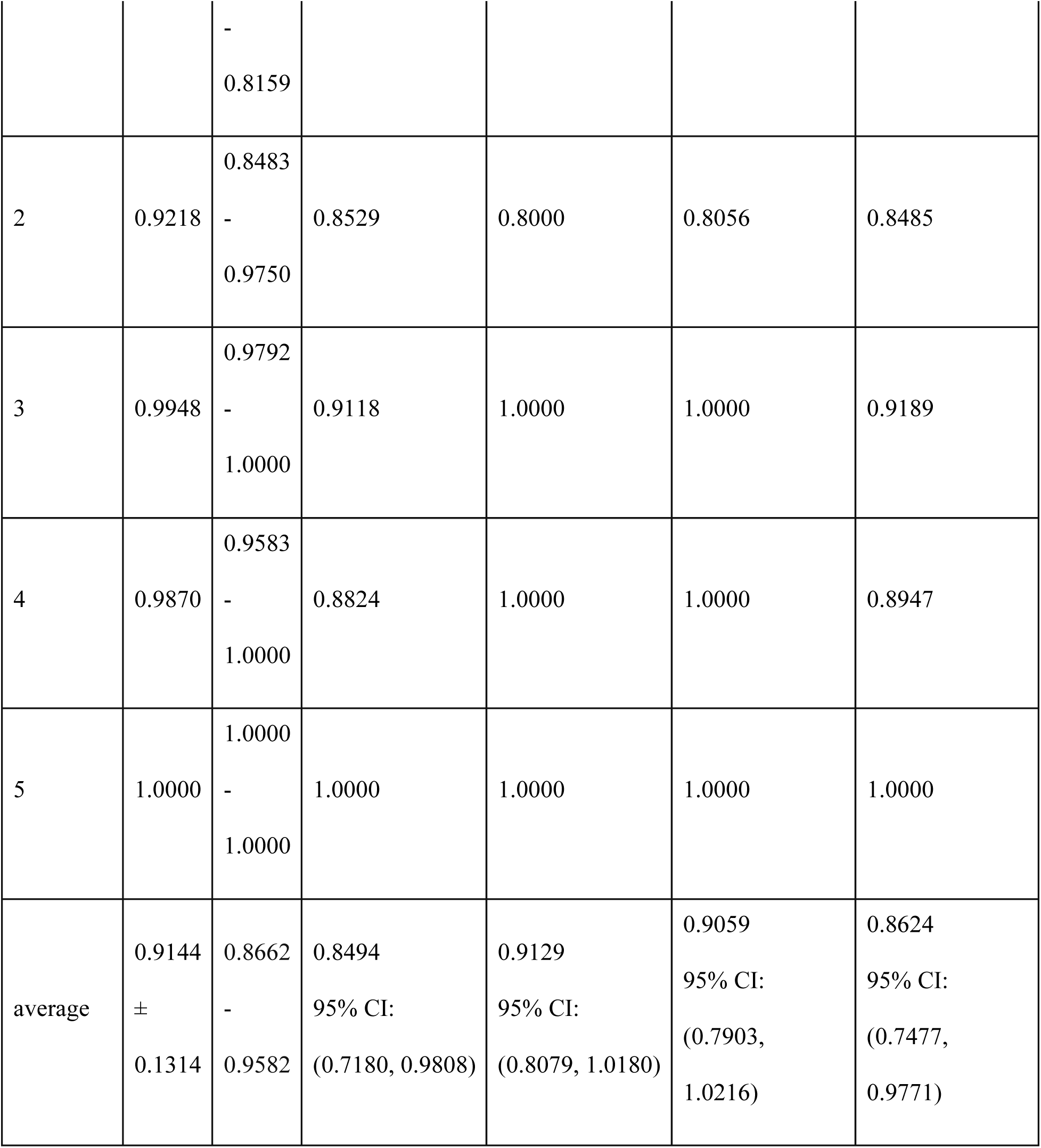

Results after removing column “Esoph Temp”:

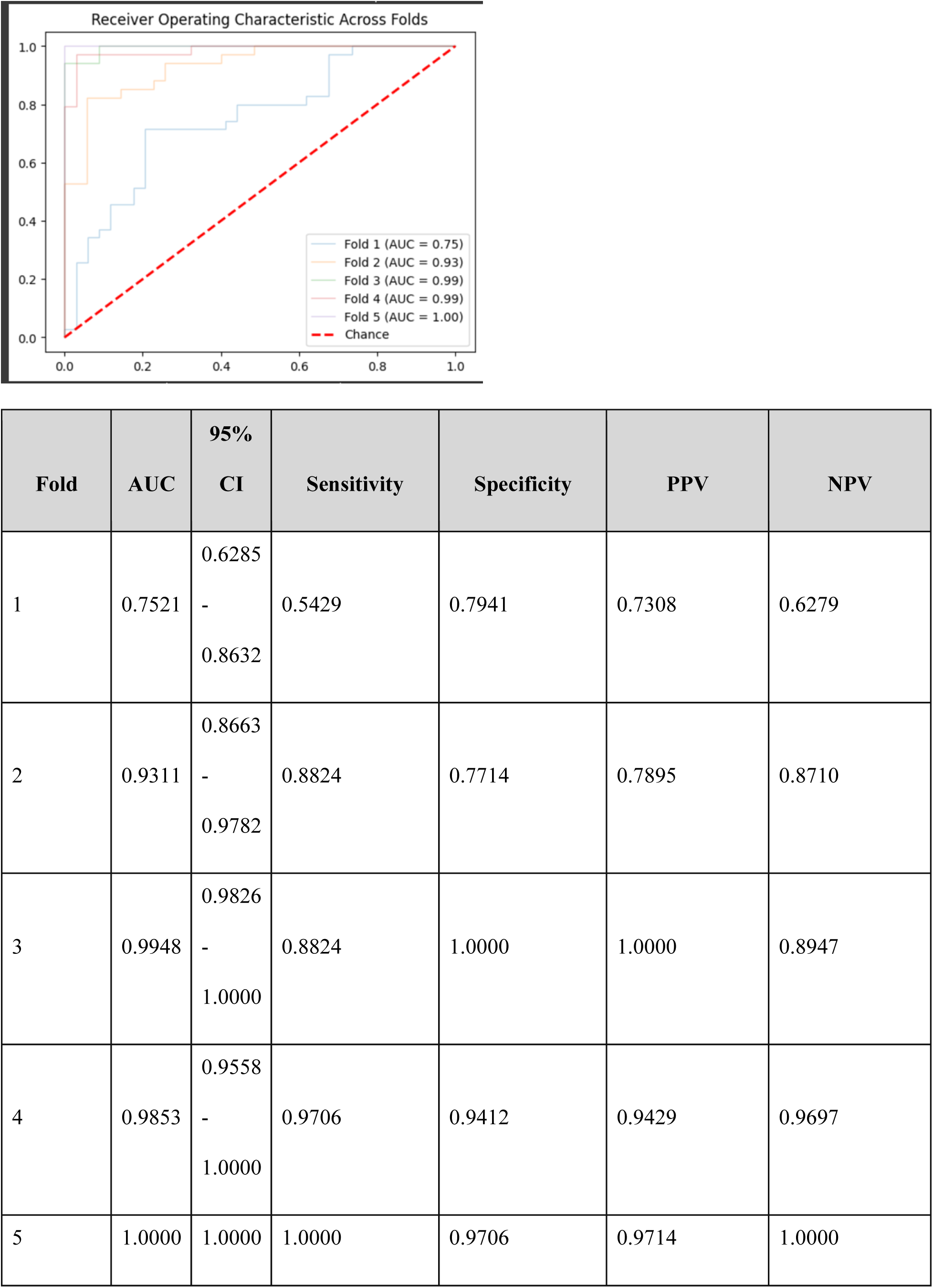
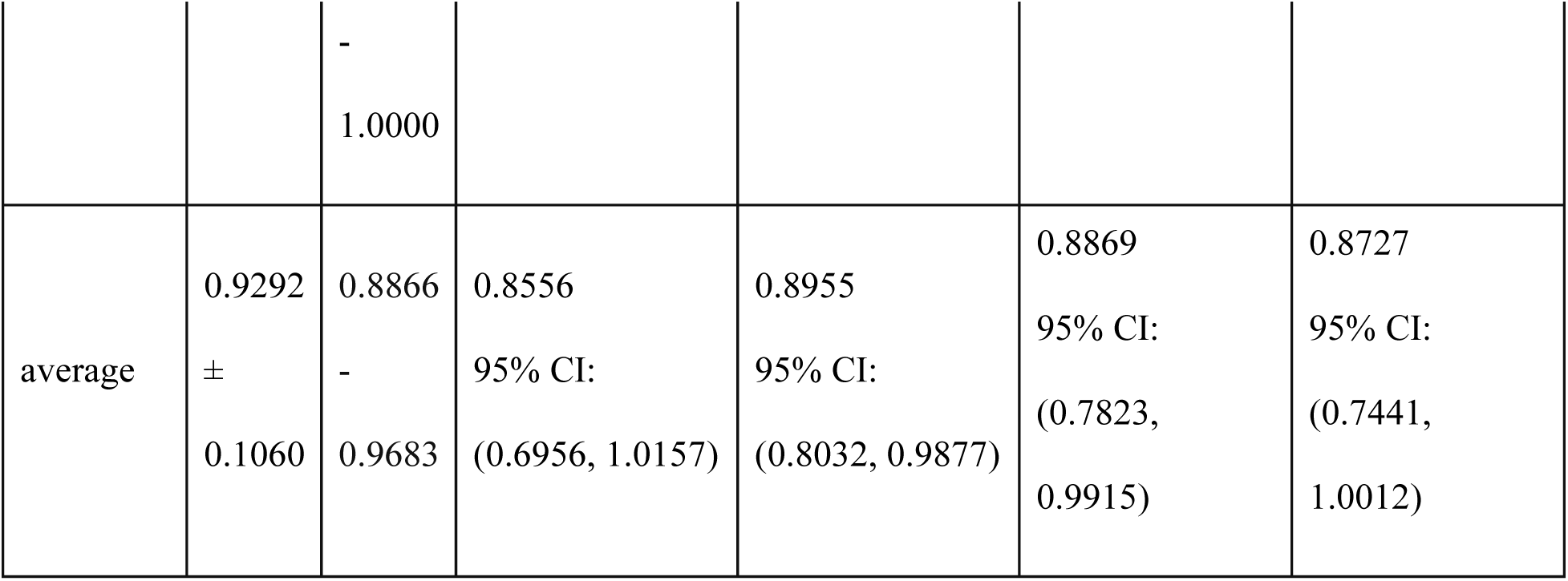

Results after removing column “Naso Temp”:

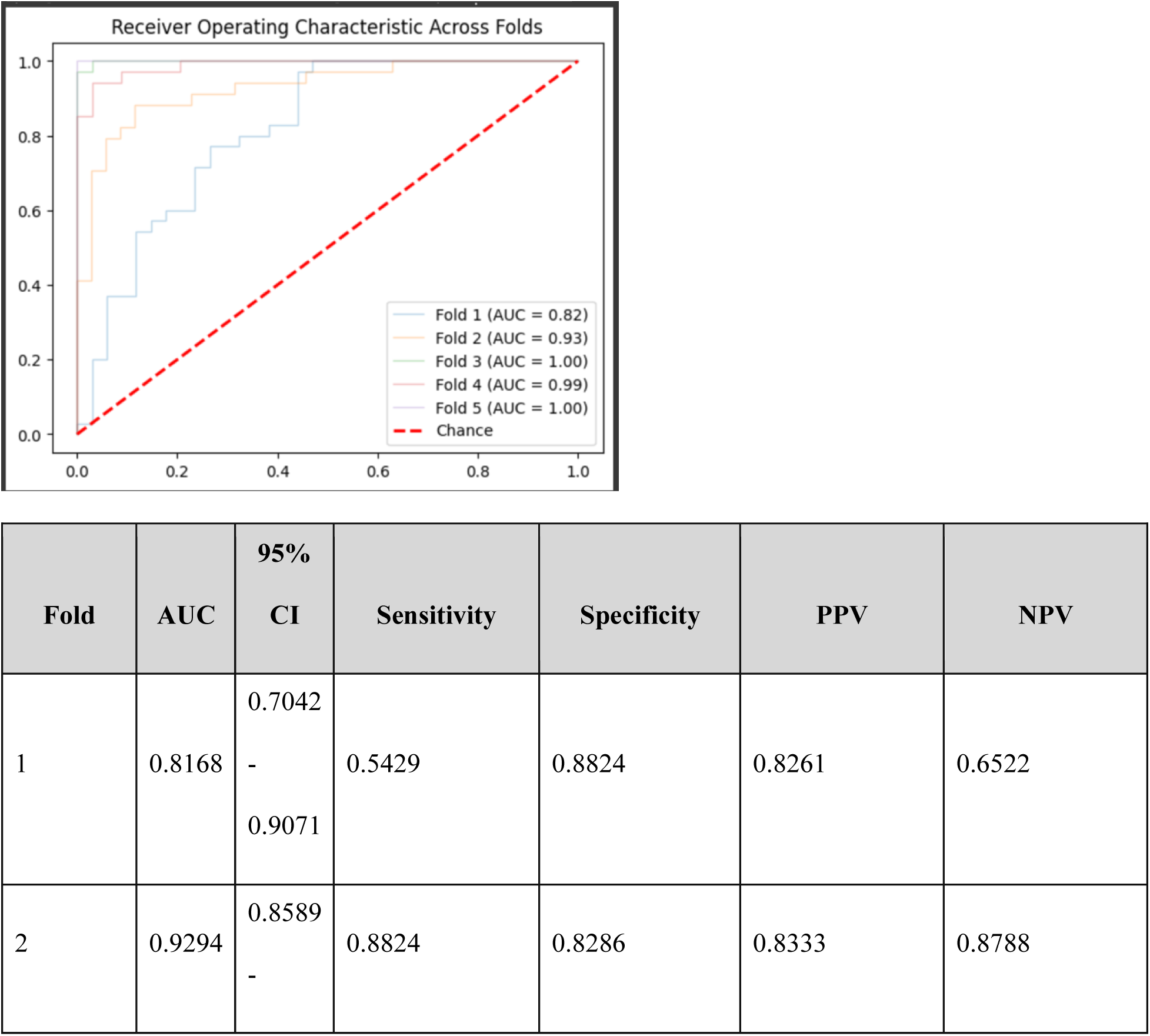
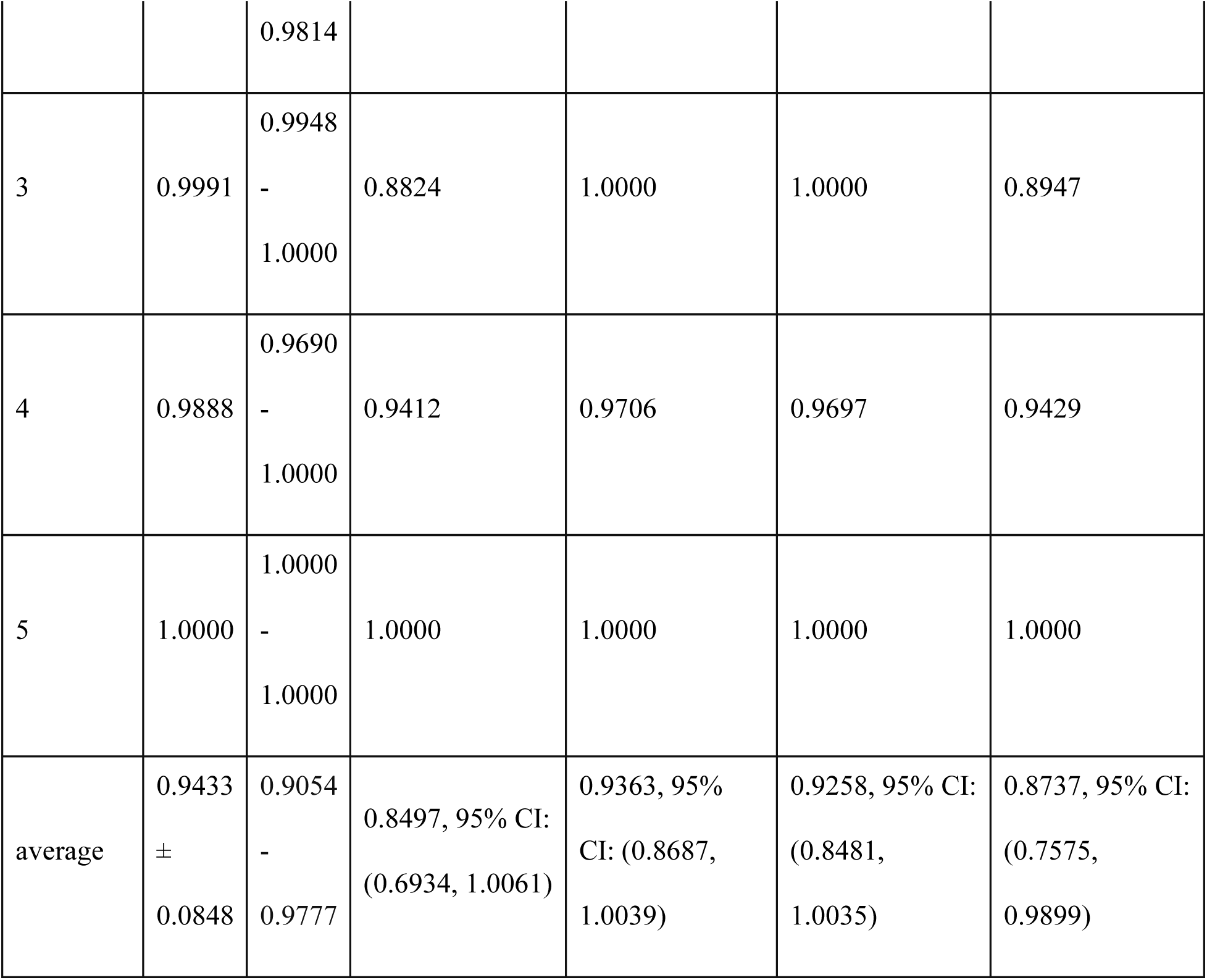

Results after removing column “Pulse Rate”:

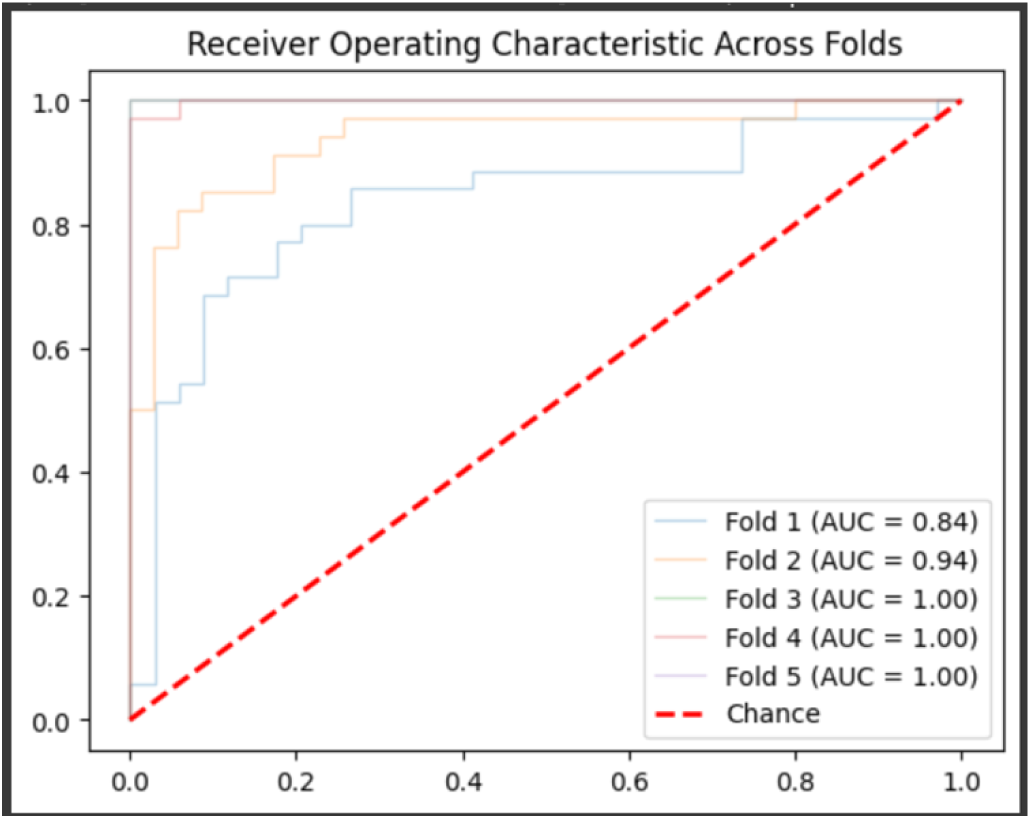
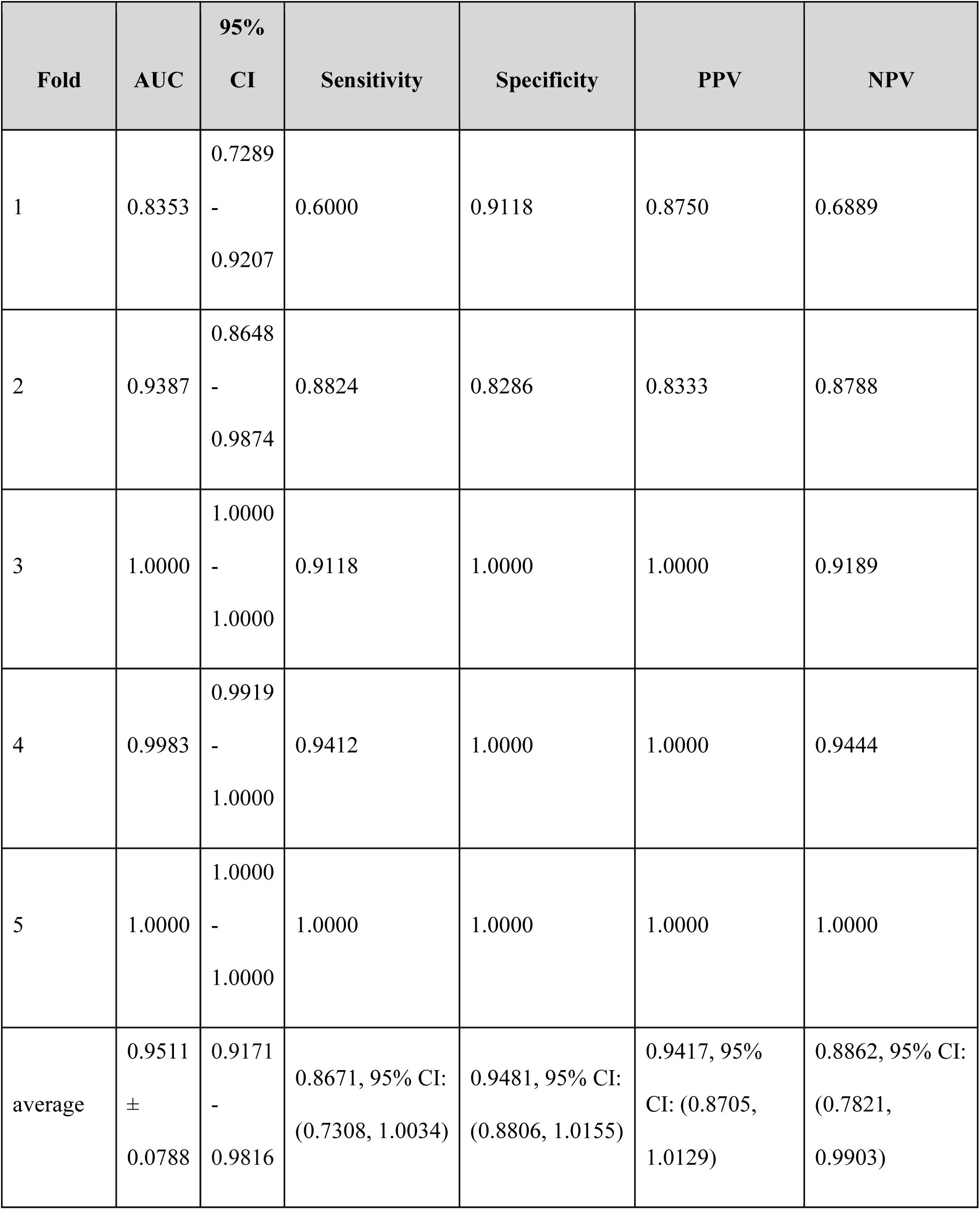

Results after removing column “ST generic V”:

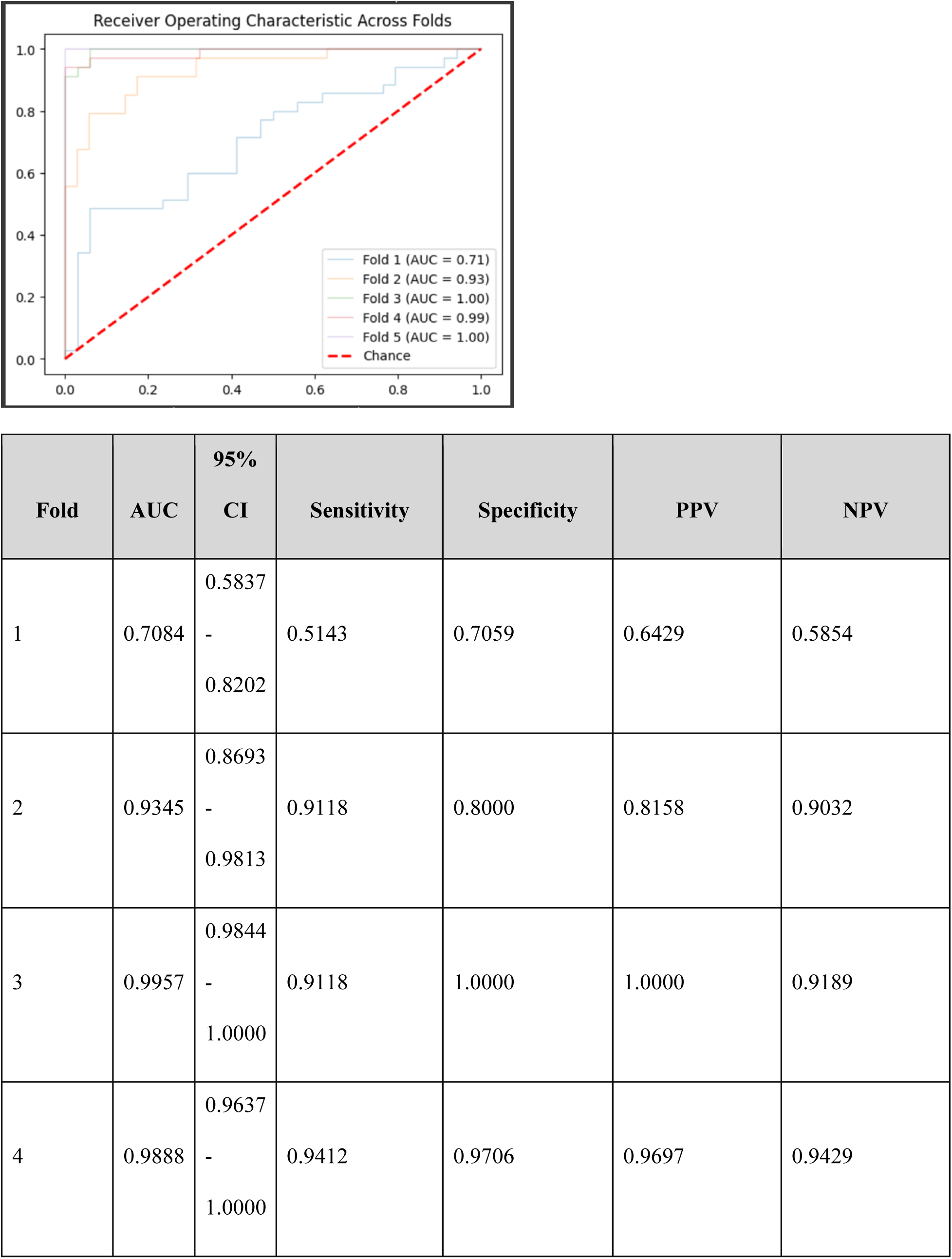
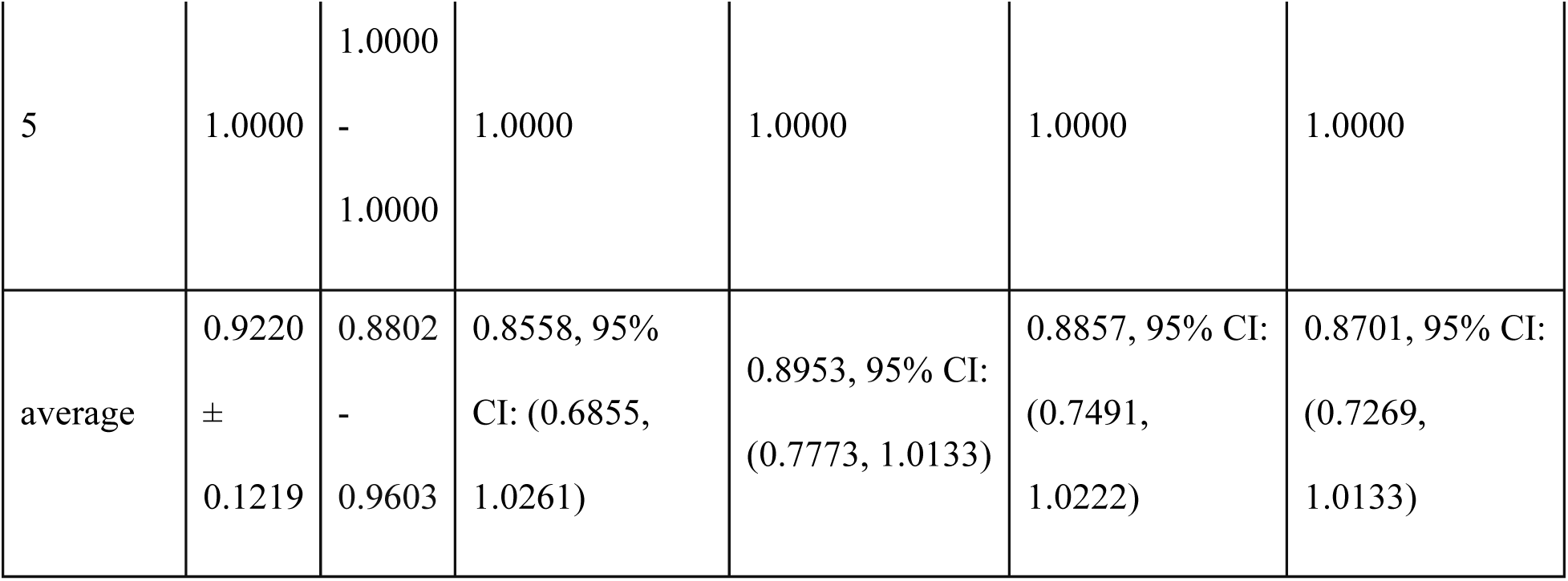

Results after removing column “bypass”:

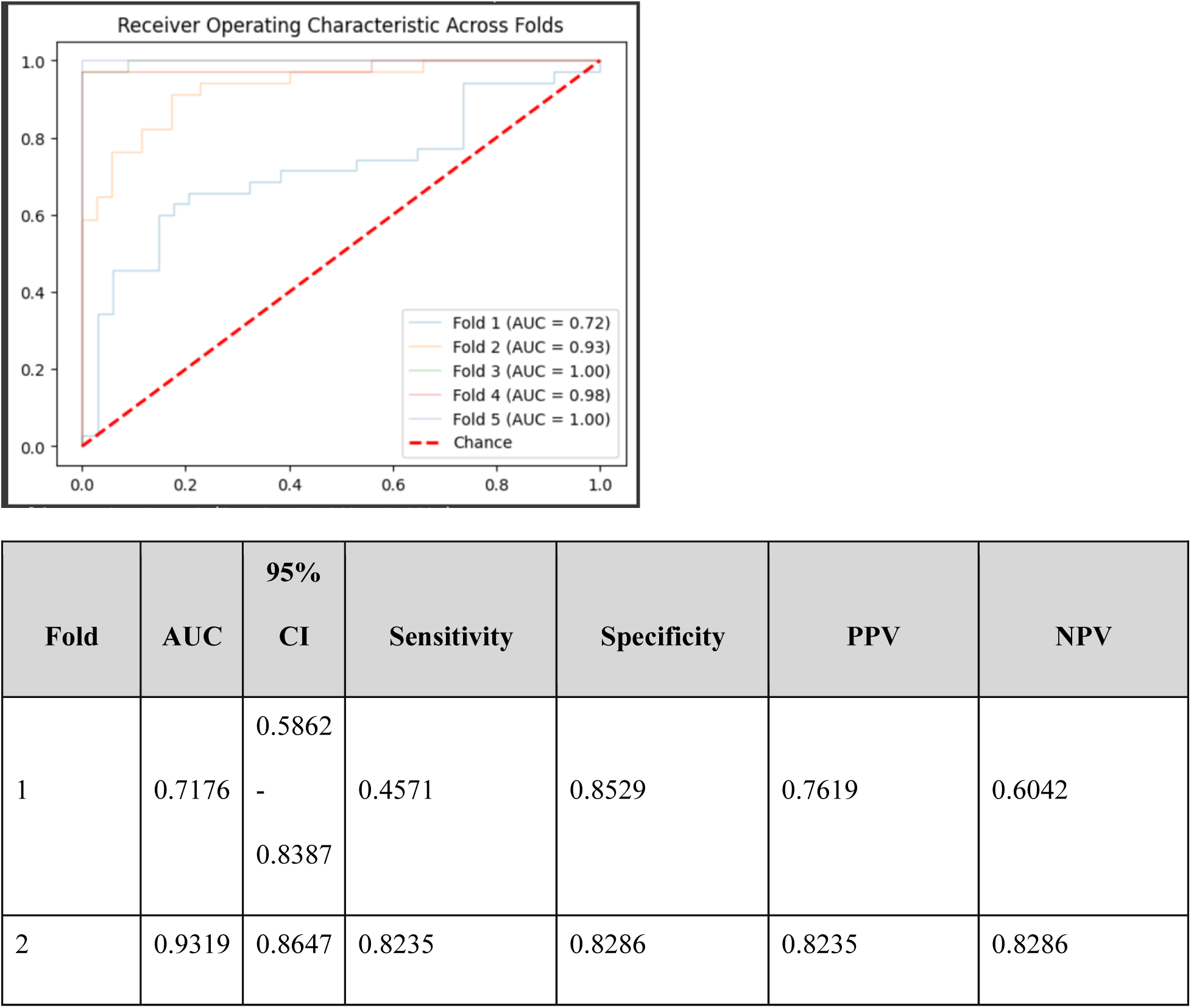
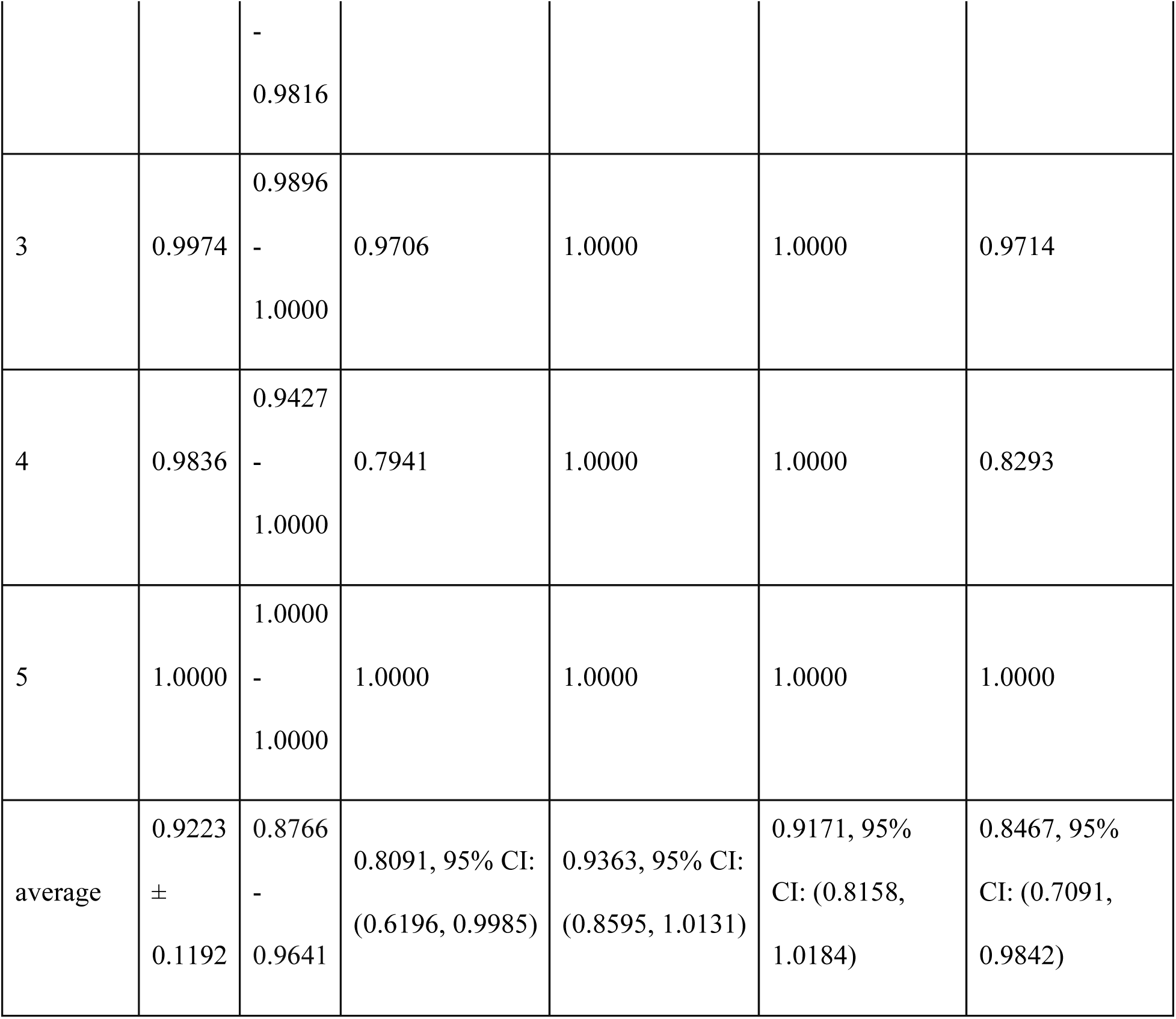

Results after removing column “xclamp”:

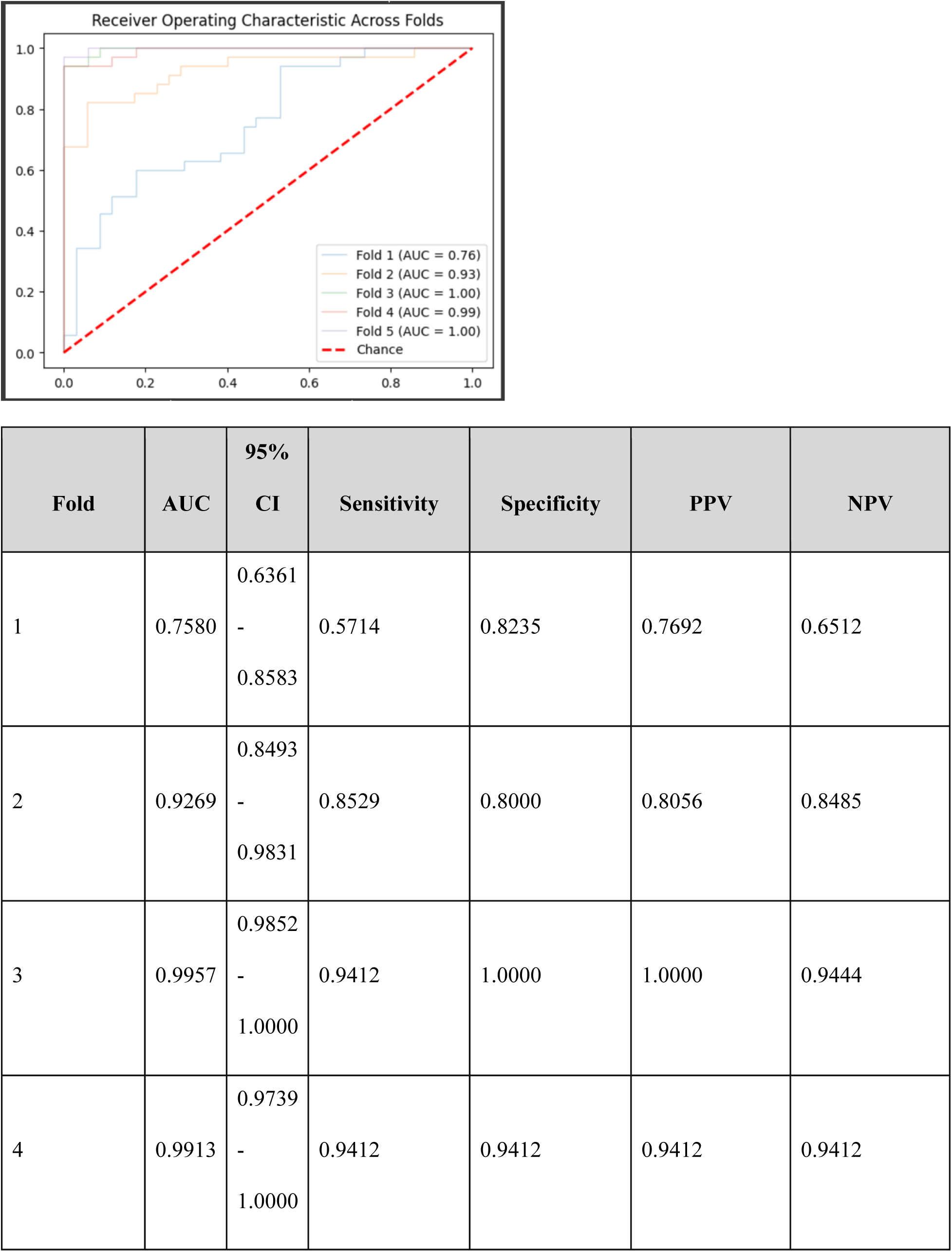
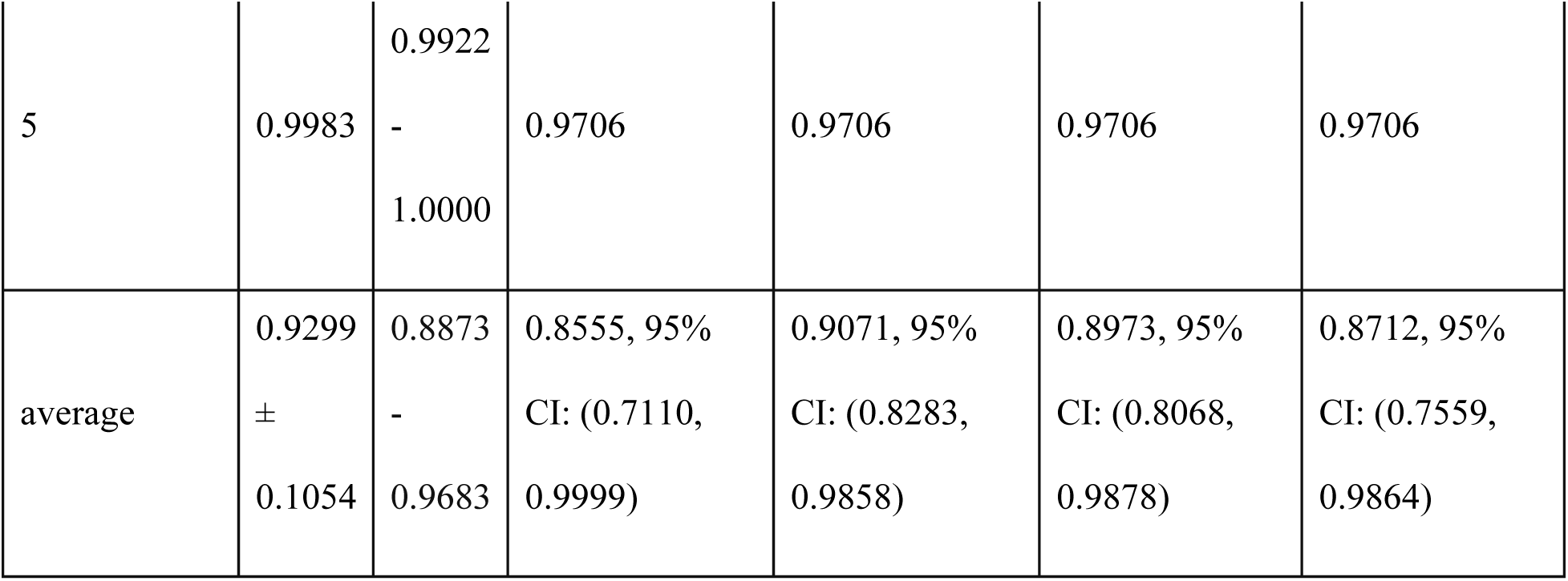

Results after removing column “Blood Temp”:

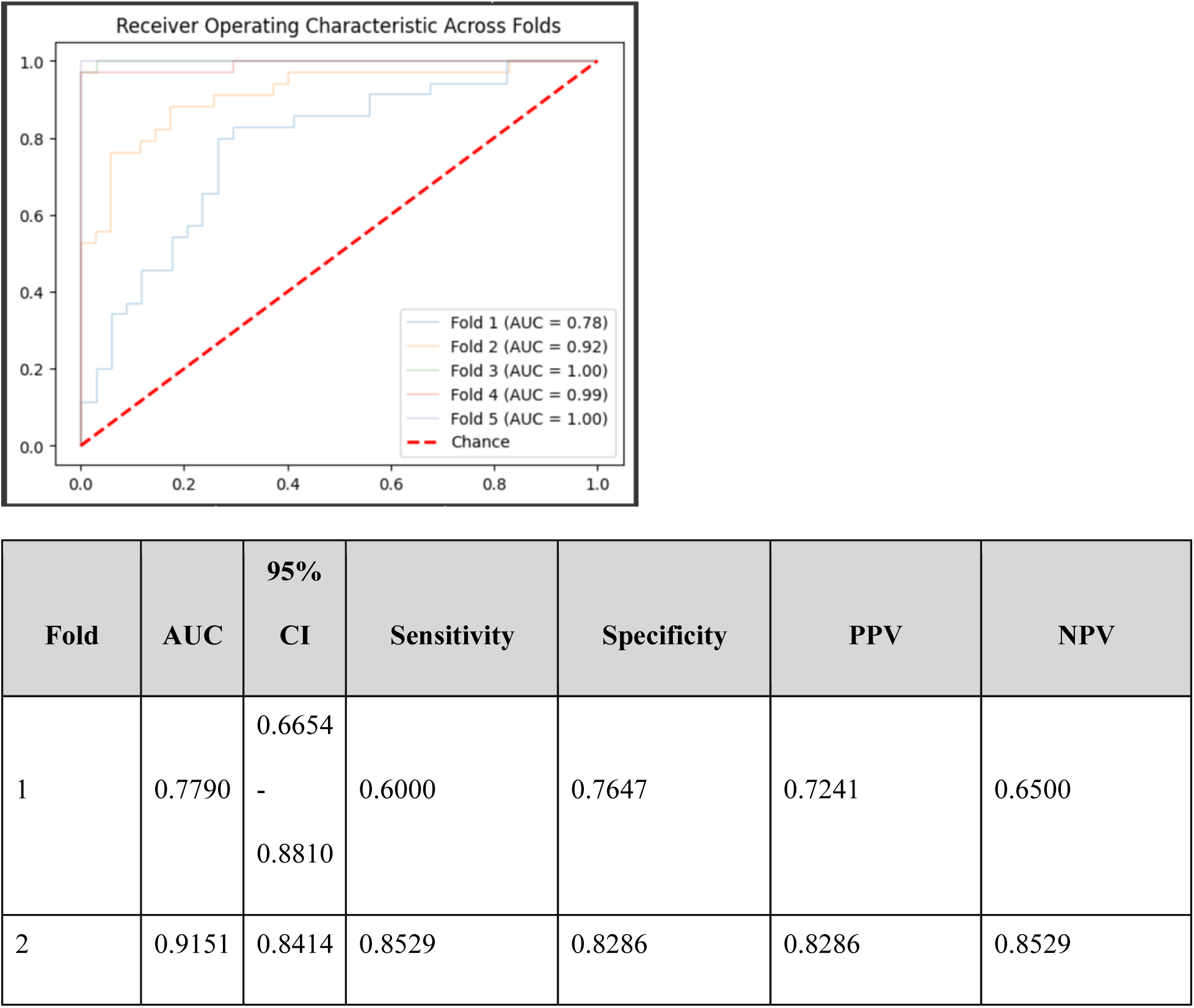
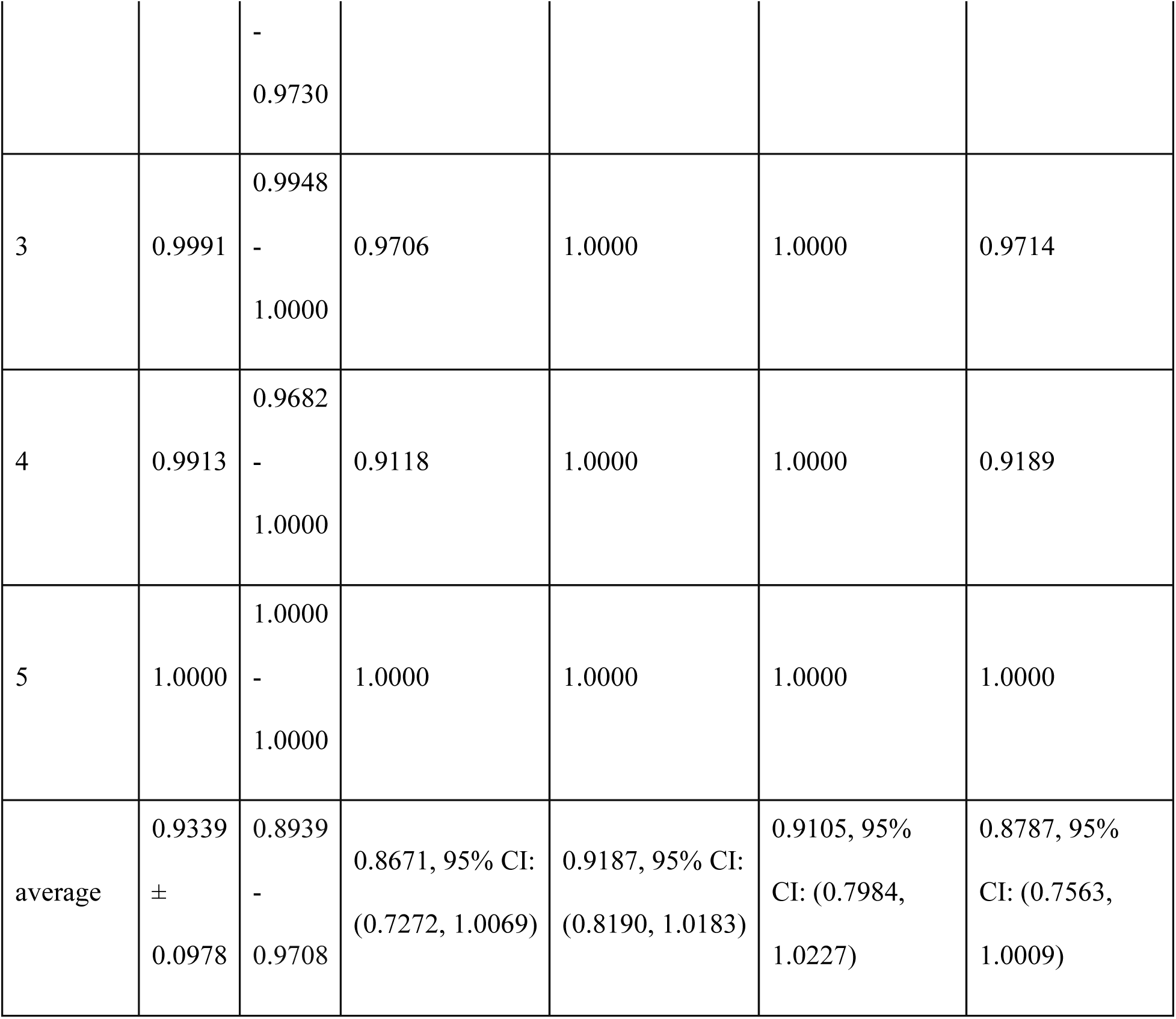

Results after removing column “Sp02”:

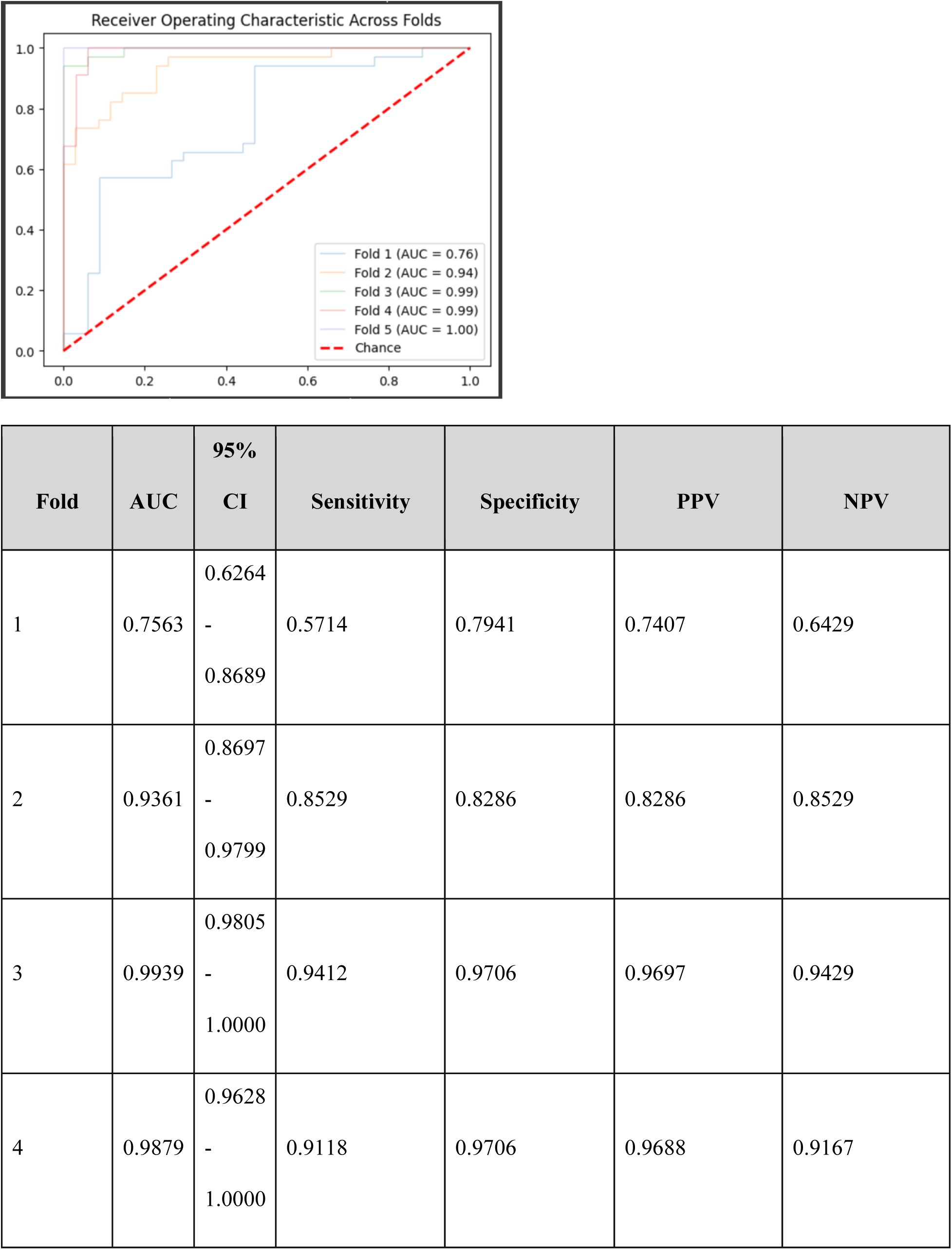
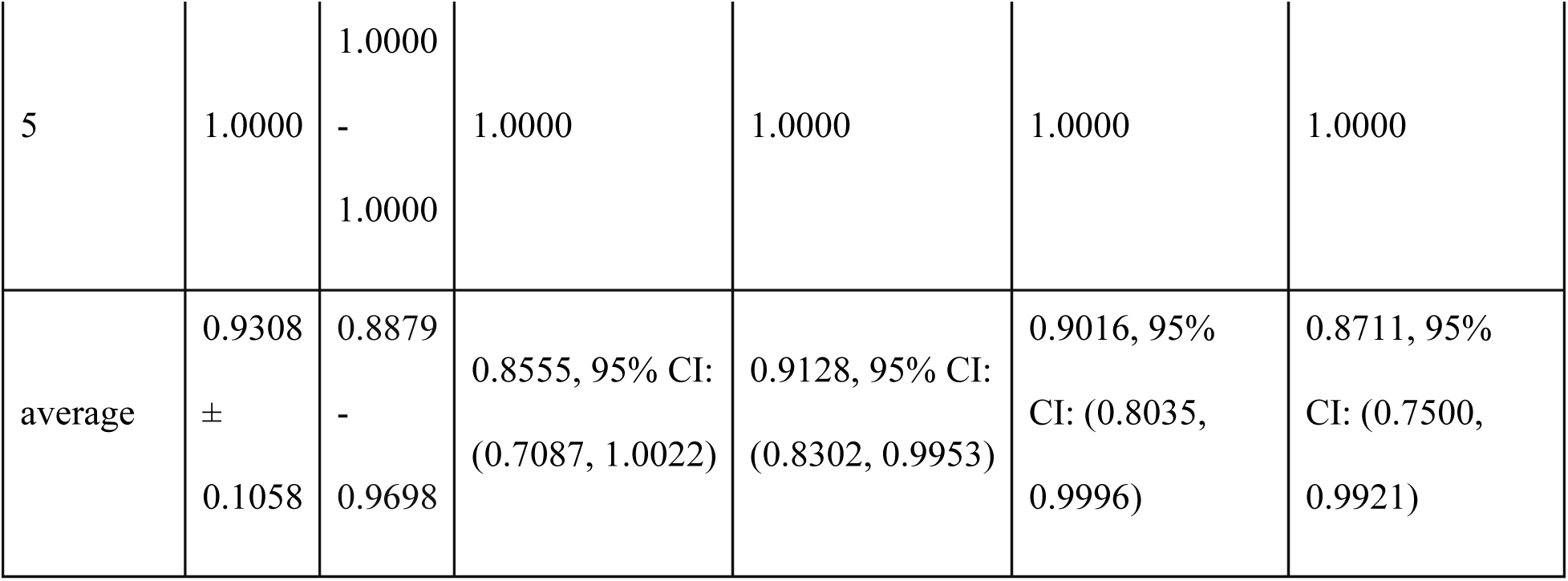

Results after removing column “ST generic II”:

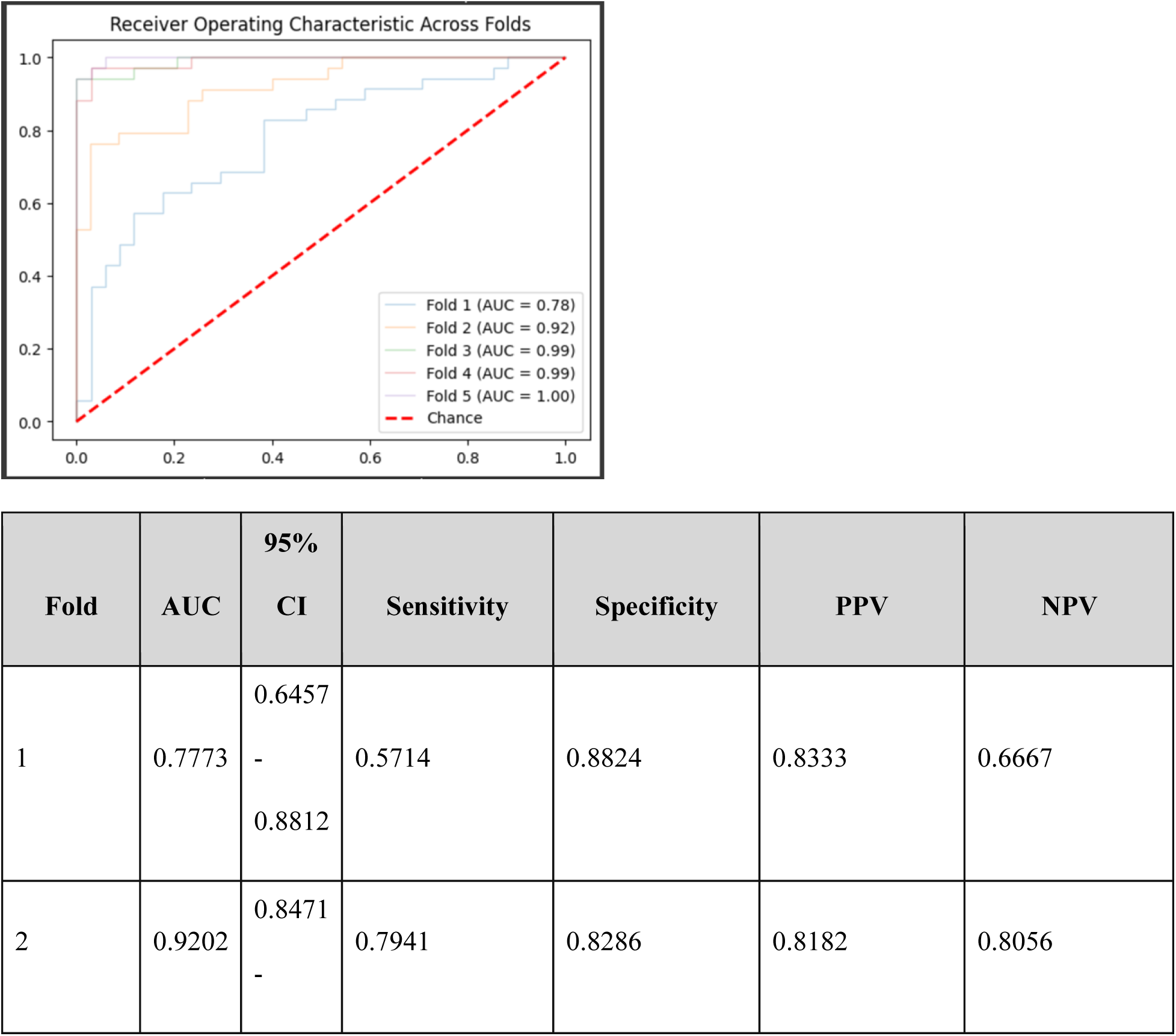
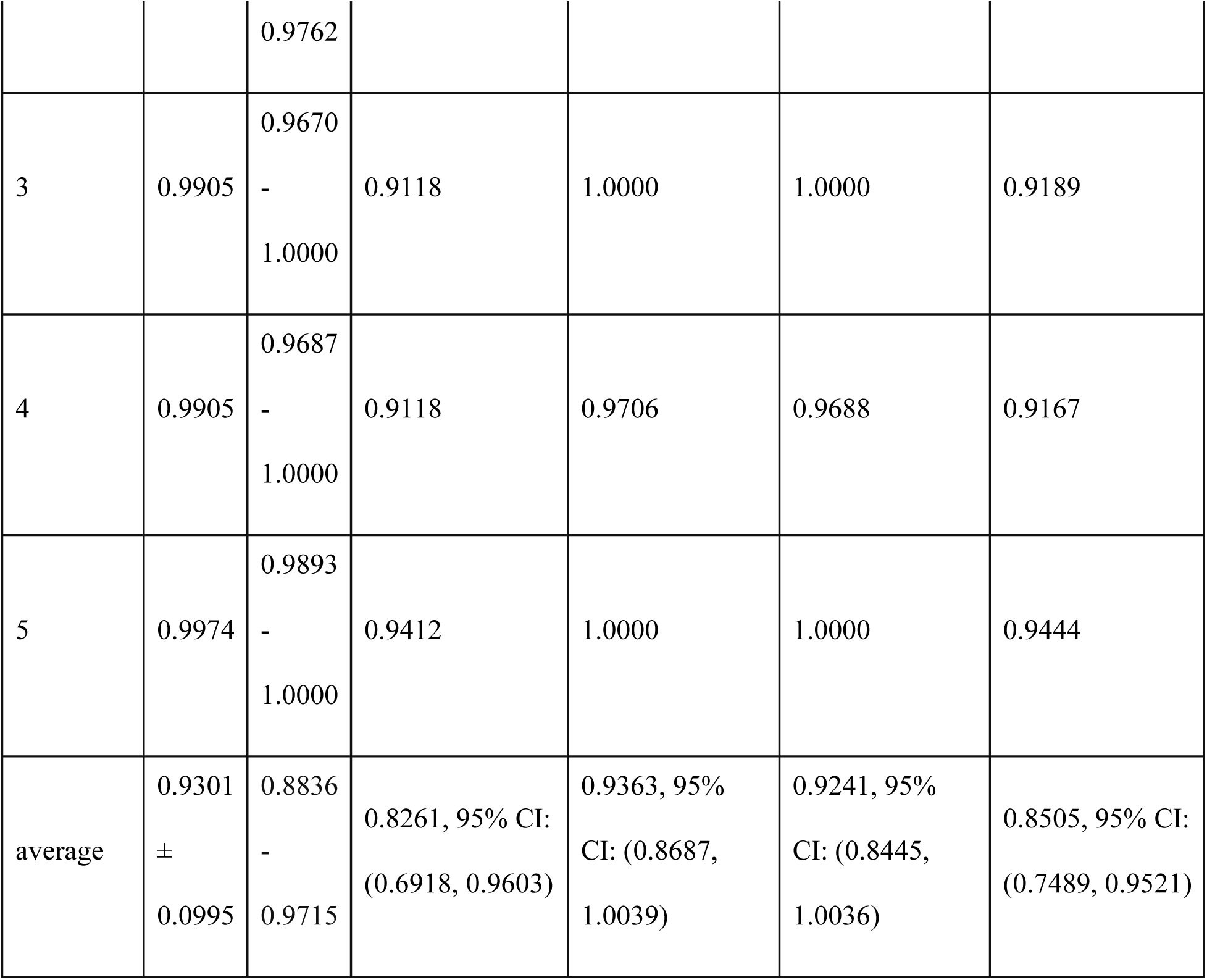

Results after removing column “PA Dia”:

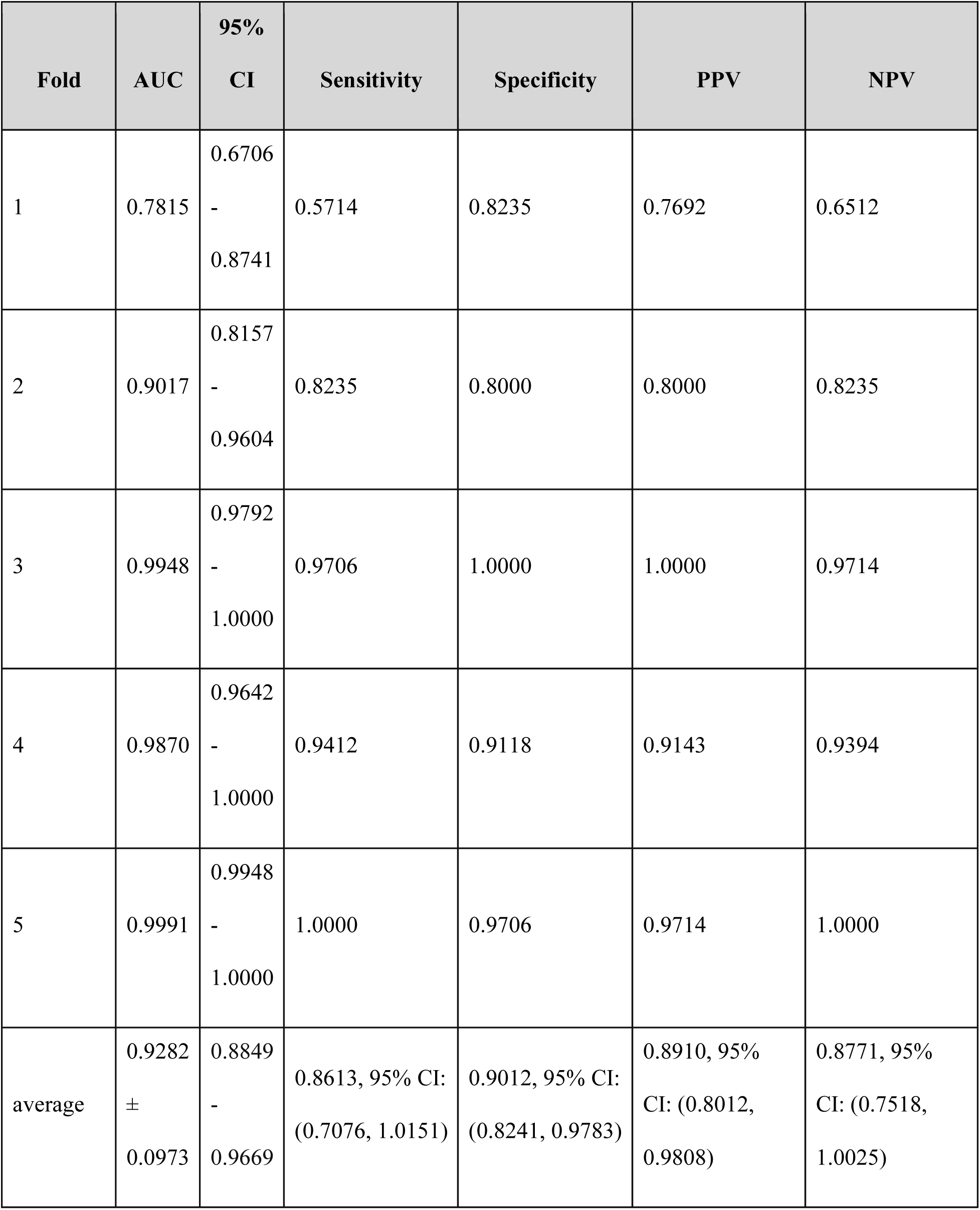

Results after removing column “PA Mean”:

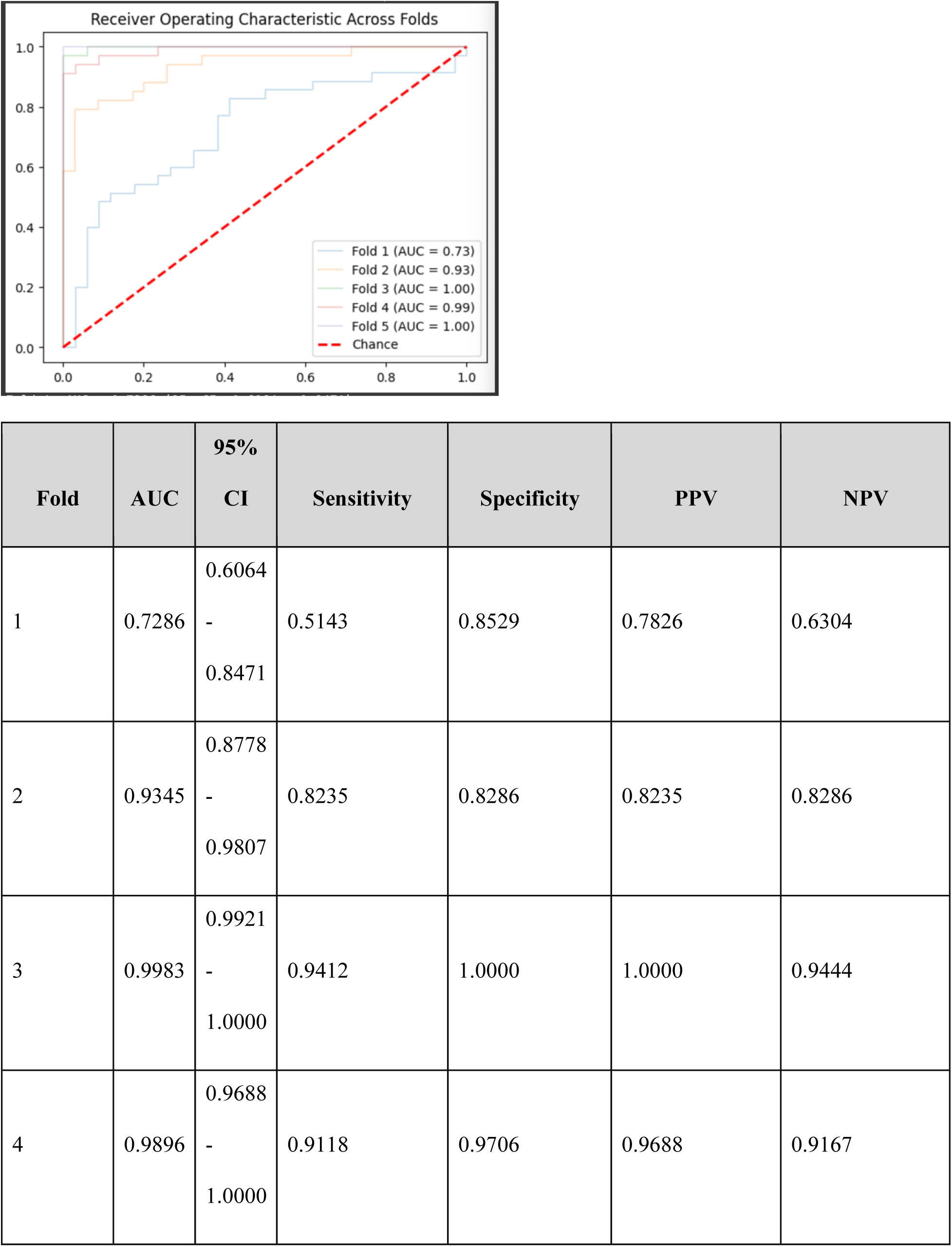
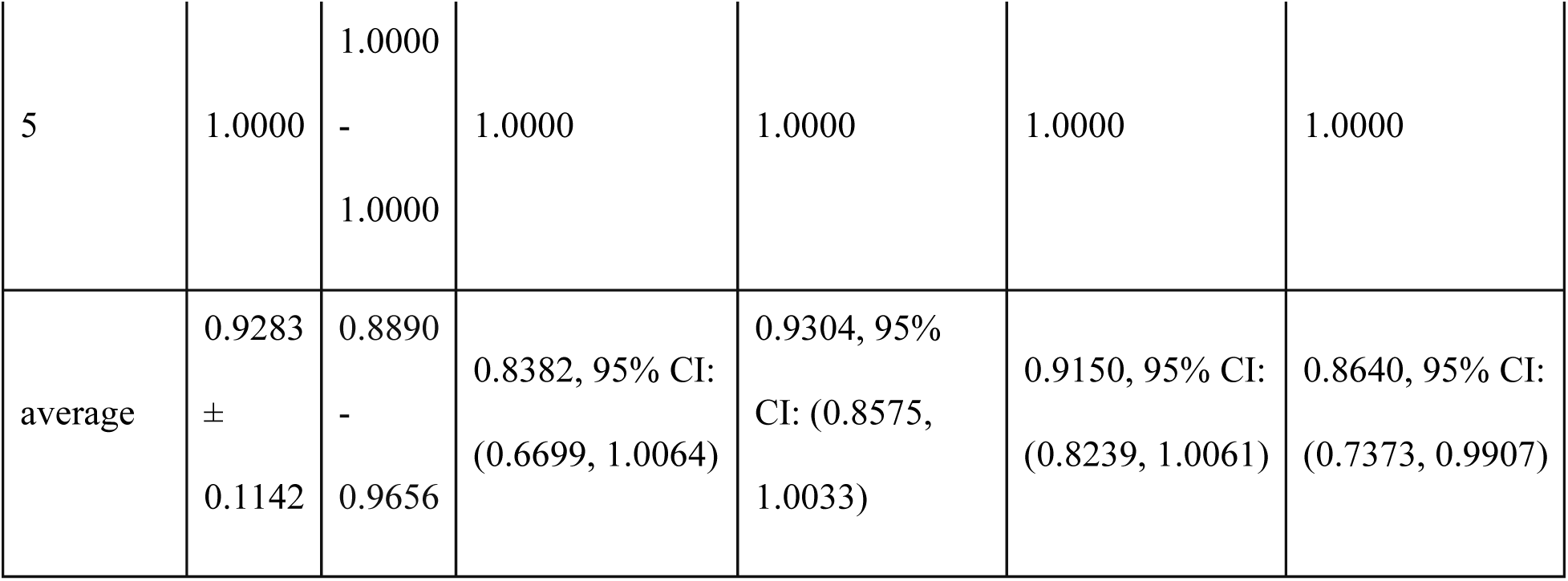

Results after removing column “PA Sys”:

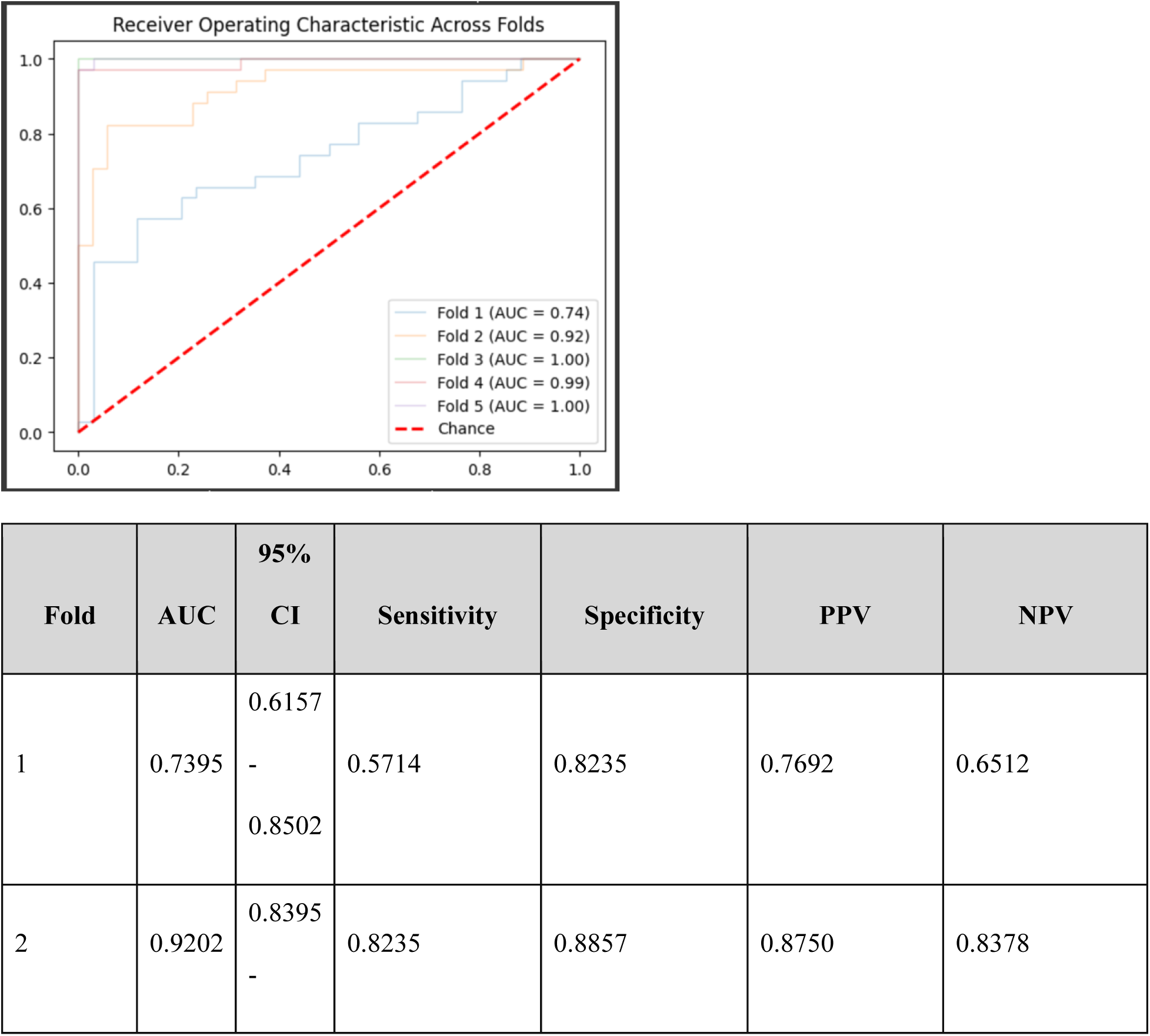
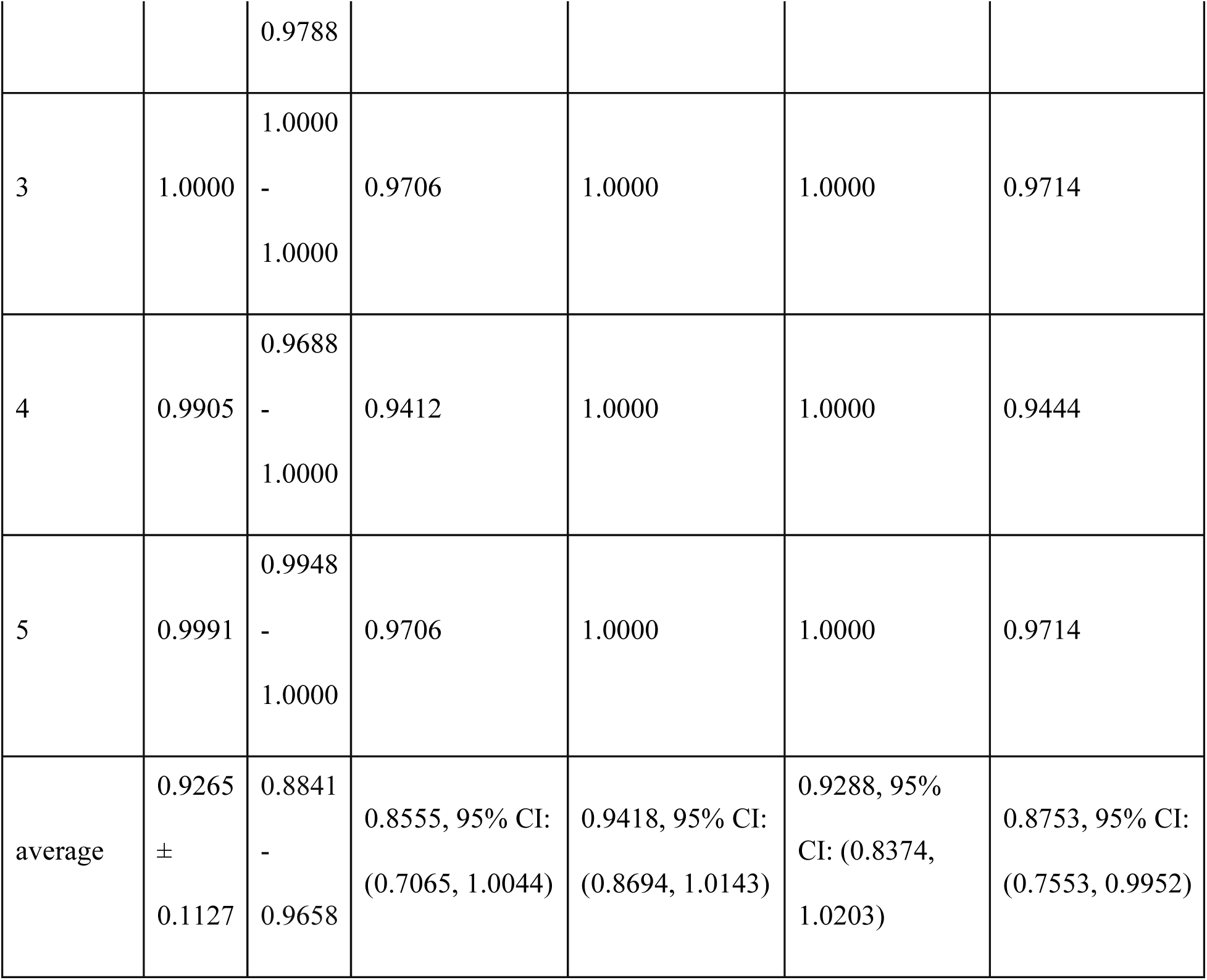

## Results from Unbalanced Data

We tried the same model with the unbalanced dataset with about 1299 patients in the control cohort with no adverse events and 171 patients in the cases cohort with at least one adverse event, for which Mean AUC of 0.93, Mean Sensitivity of 0.64, Mean Specificity of 0.99, Mean PPV of 0.86, Mean NPV of 0.95 was achieved.

**Figure 3a:**
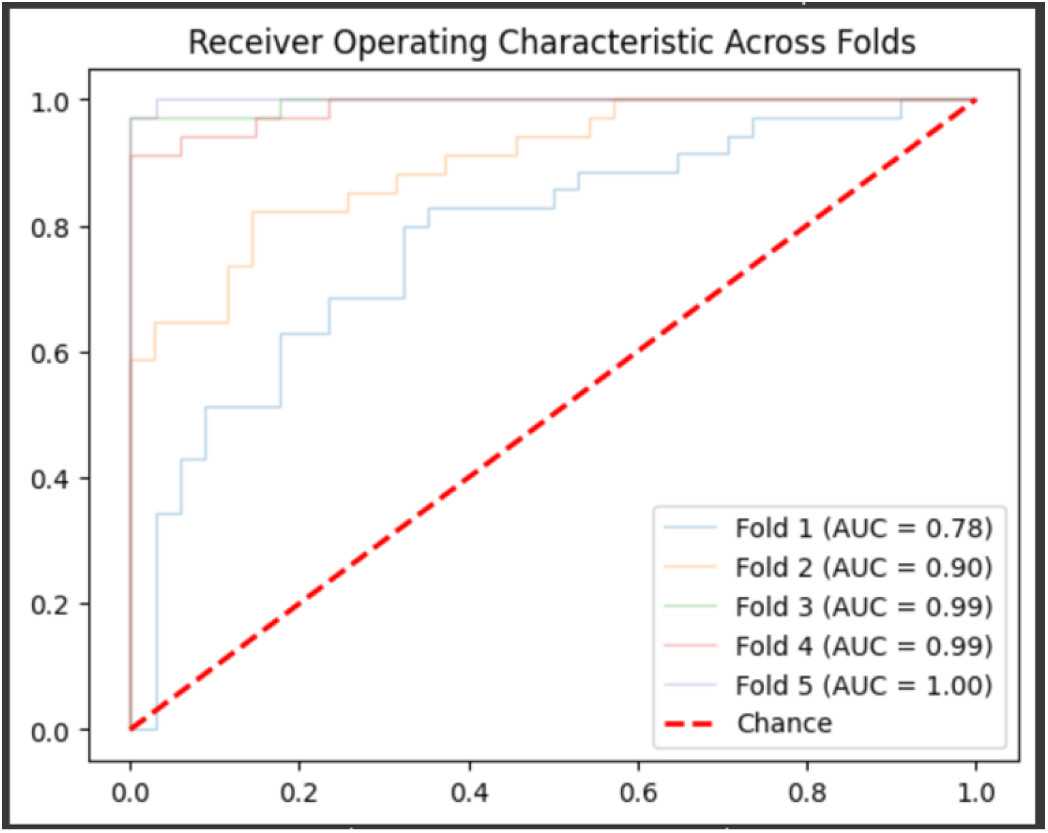
Area under the curve for the FC + LSTM model with unbalanced classes

**Table 4a:**
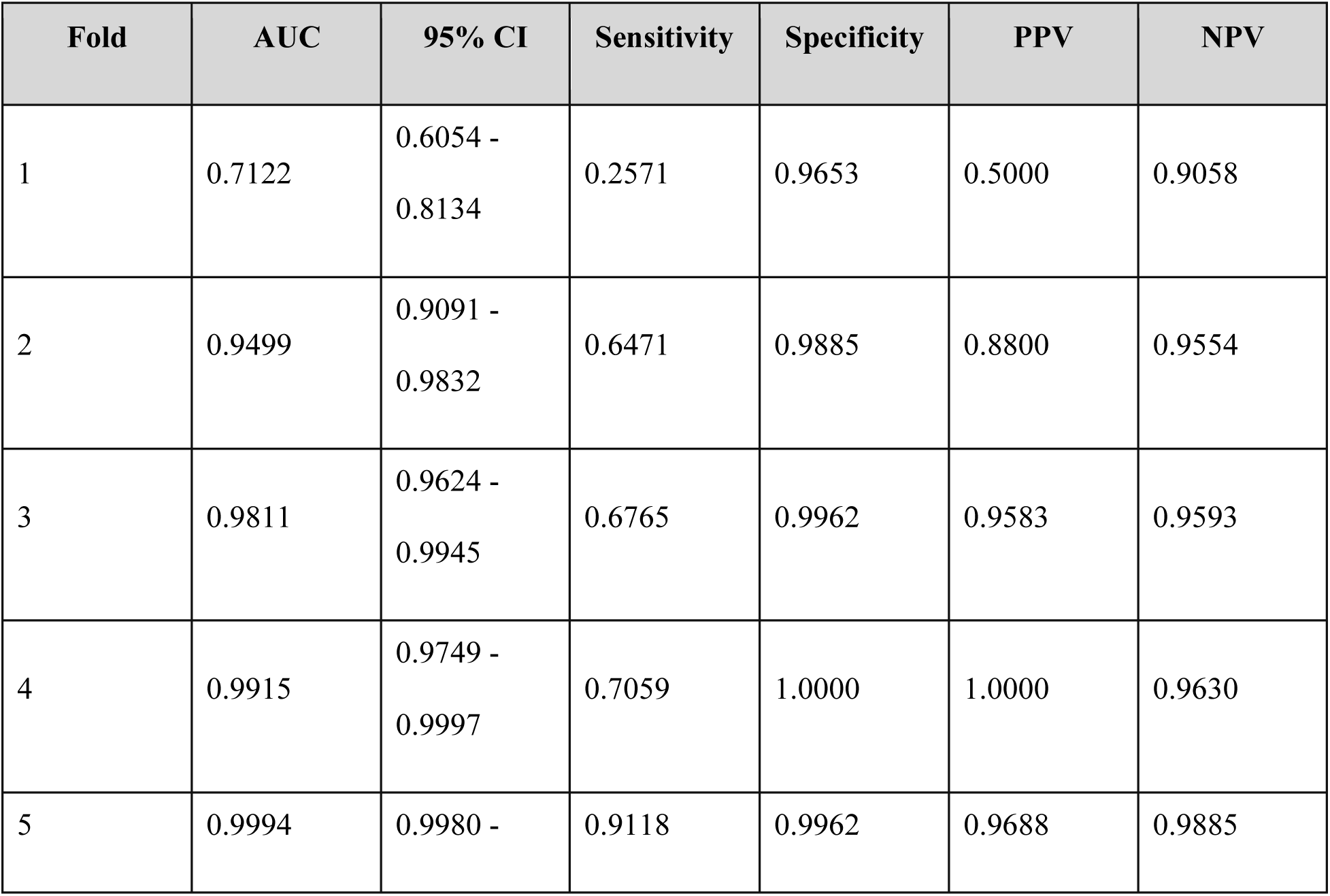
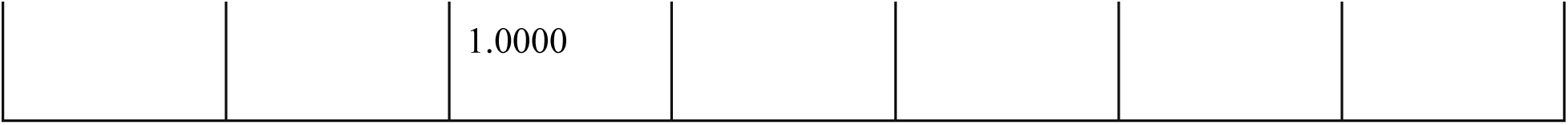
Performance of the model across all 5 folds in unbalanced dataset

## Ablation Study

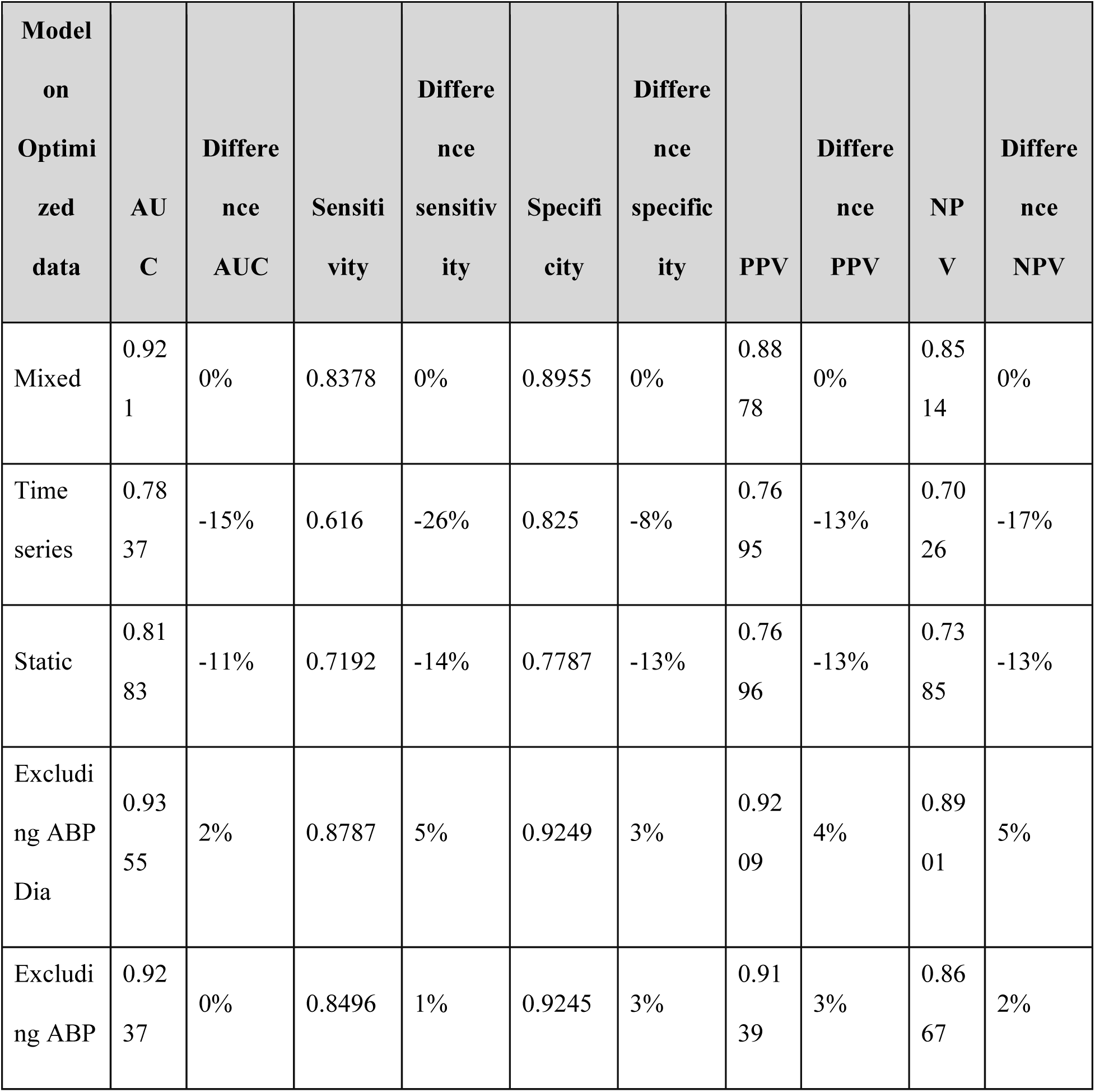
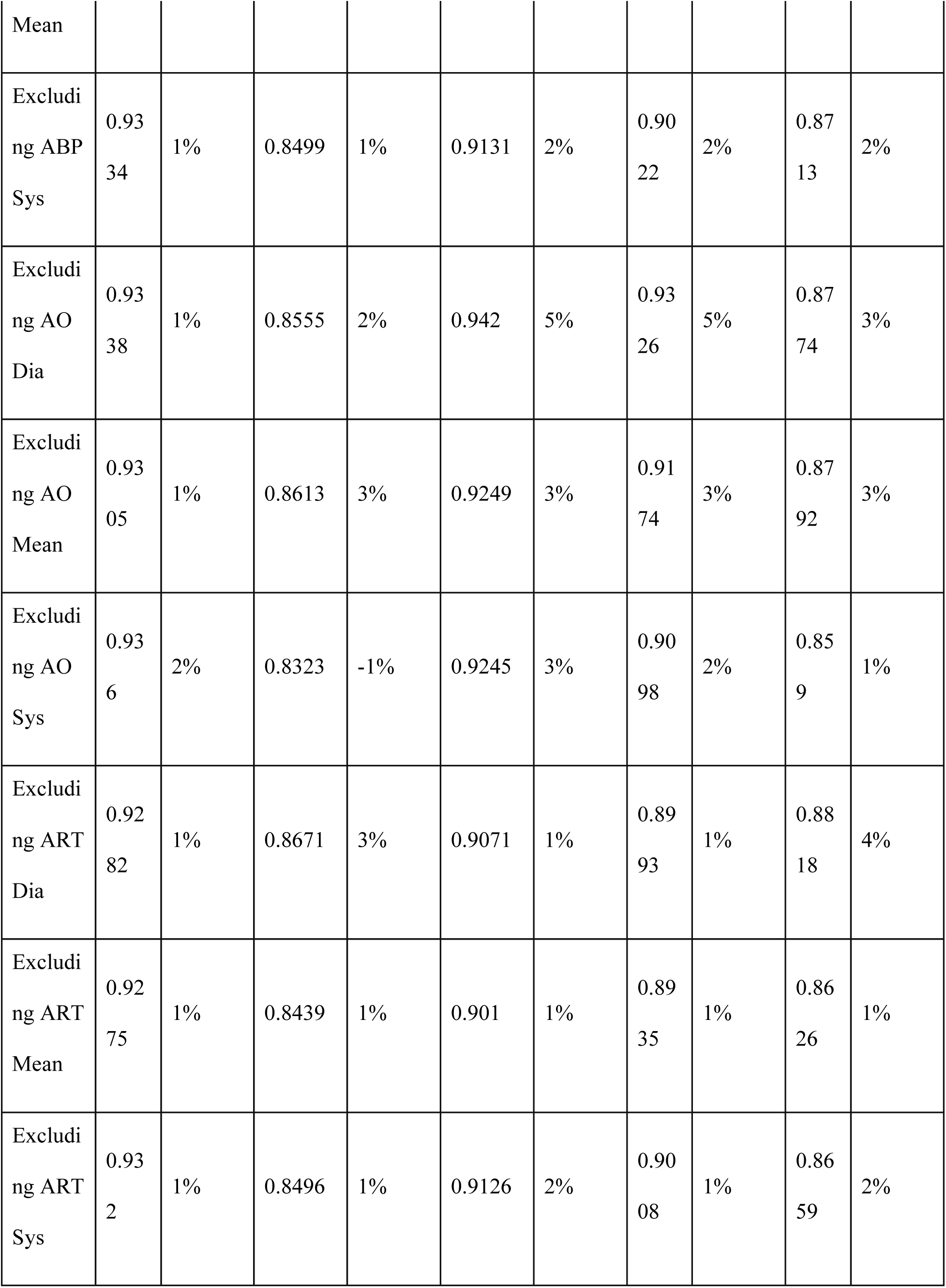
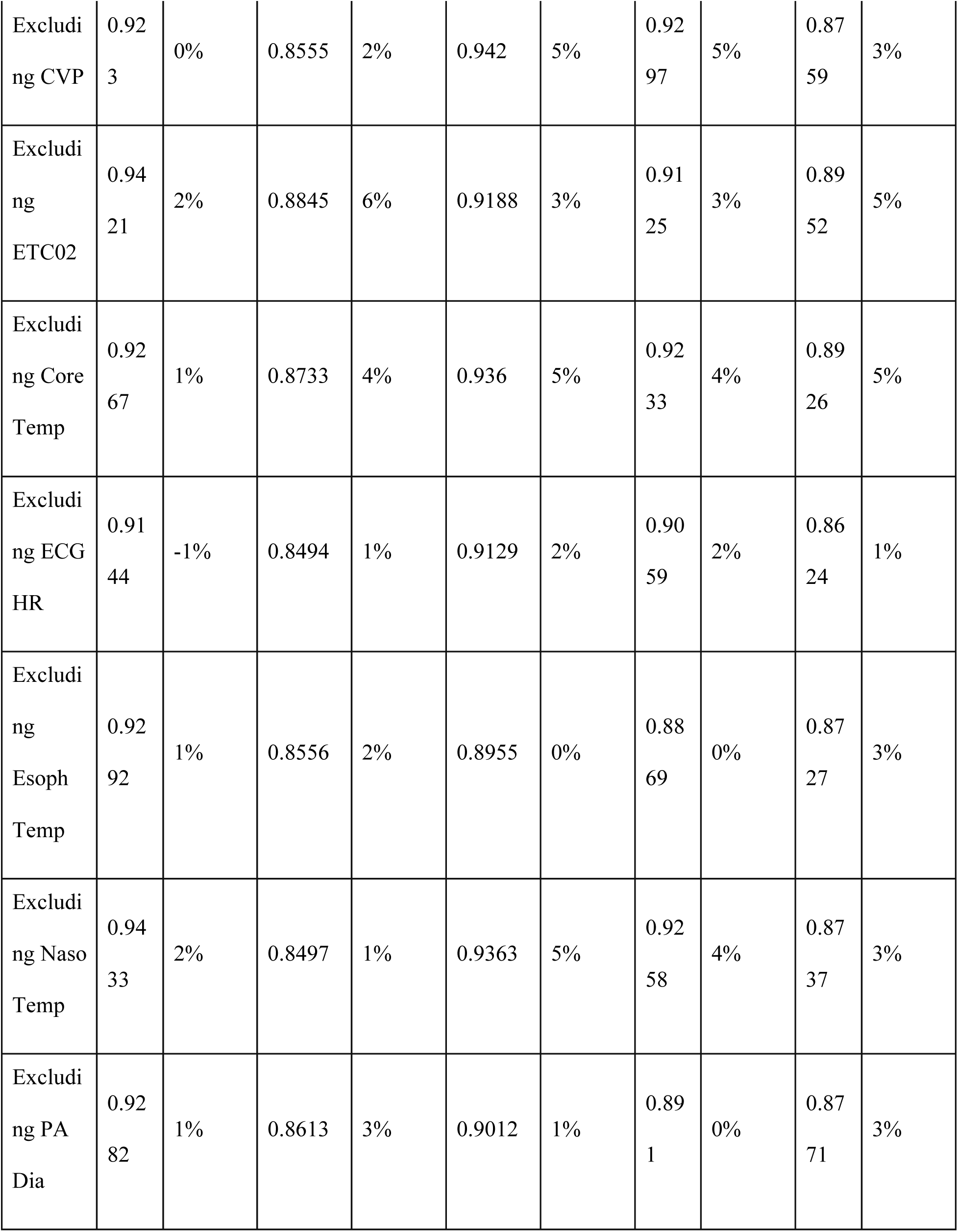
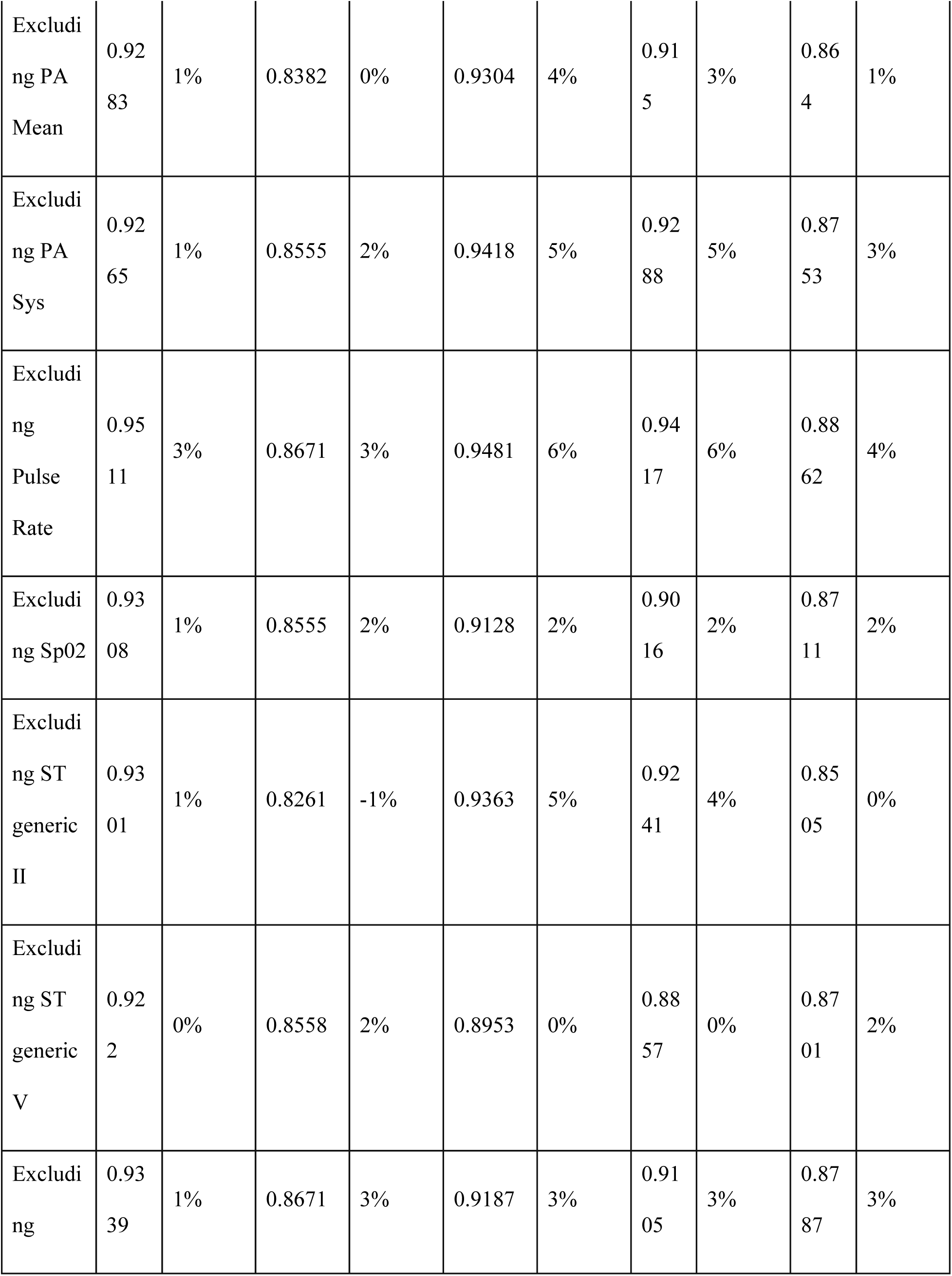
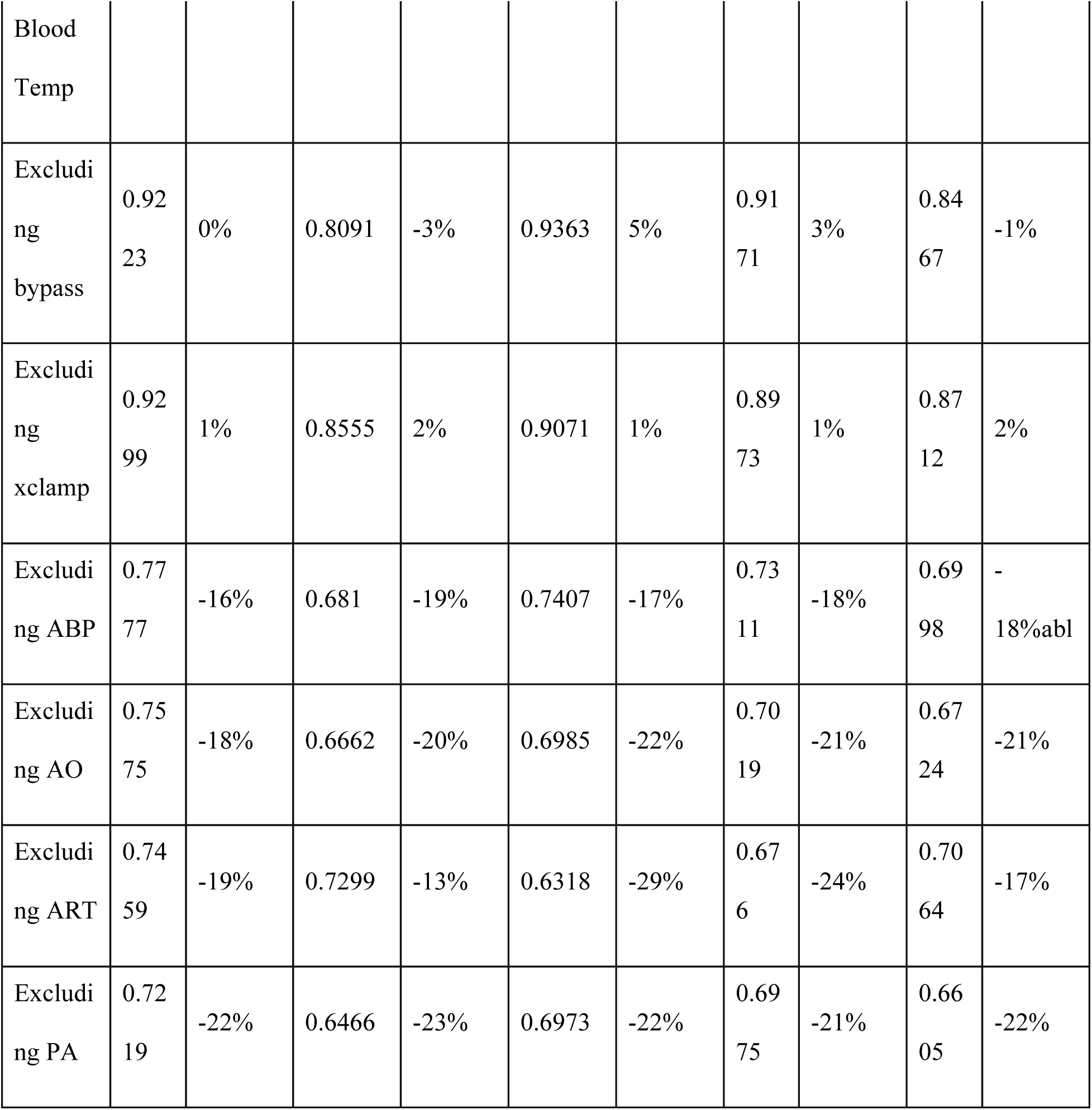

## Glossary

ACE: angiotensin-converting enzyme
ADP: adenosine diphosphate
ARB: angiotensin-receptor blocker
ASCD: Adult Cardiac Surgery Database
AUC: Area under the curve
AVR: aortic valve replacement
CABG: coronary artery bypass grafting surgery
CAD: coronary artery disease
CBA: Catheterization-based assist device
CI: Confidence interval
CVA: Cerebrovascular accident
CVD: Cardiovascular disease
CTICU: Cardiothoracic Intensive Care Unit
DSWI: Deep sternal wound infection
ECMO: Extracorporeal membrane oxygenation
FTR: Failure to rescue
IABP: Intra-aortic balloon pump
ICD: Implantable cardioverter-defibrillator
LAD: Left anterior descending artery
MMC: Maine Medical Center
NPV: Negative predictive value
NQF: National Quality Forum
PAD: Peripheral arterial disease
PCI: percutaneous coronary intervention
POLOS: Postoperative length of stay
PPV: Positive predictive value
ROC: Receiver operating curve
STS: Society of Thoracic Surgeons
TIA: Transient ischemic attack

## Notes

### Competing Interest Statement

The authors have declared no competing interest.

### Funding Statement

This study was funded by COBRE program at MaineHealth Institute for Research

